# Artificial Intelligence for Surgical Scene Understanding: A Systematic Review and Reporting Quality Meta-Analysis

**DOI:** 10.1101/2025.07.12.25330122

**Authors:** Matthias Carstens, Shubha Vasisht, Zheyuan Zhang, Iulia Barbur, Annika Reinke, Lena Maier-Hein, Daniel A. Hashimoto, Fiona R. Kolbinger

## Abstract

Surgical scene understanding (SSU) describes the use of Artificial Intelligence (AI) to provide an understanding of visual components of surgical imaging data, such as laparoscopic surgery videos. While hundreds of publications report AI capabilities to identify instruments, anatomical structures, and other contextual data and testify potential for real-time support in the operating room, the clinical implementation of SSU remains limited.

This systematic review and meta-analysis (registered in the PROSPERO database under CRD420251005301) assesses the current state and research gaps in computational SSU, focusing on data curation, model design, validation, uncertainty estimation, performance metrics, reporting quality, and clinical applicability. Studies were included if they analyzed intraoperative data from minimally invasive abdominal surgeries in humans, developed computational SSU methods, and reported trainable models with formal validation and performance metrics.

A total of 188 studies from six literature databases were included. Most relied on small, single-center datasets, often from laparoscopic cholecystectomies, with limited metadata and topical diversity. Research was largely descriptive, with limited reporting on clinical relevance, limitations, code availability, and model uncertainty. Validation was often inadequate, typically relying on simple hold-out strategies, with limited testing on external datasets and purely technical validation approaches without any clinical expert involvement. Clinical translation was addressed in only eleven works. Overall, studies showed minimal progress toward real-world application. Our findings highlight the need for diverse, multi-institutional datasets, robust validation practices, and clinically driven development to unlock the full potential of SSU in surgical practice.

## Introduction

Artificial Intelligence (AI) applications for clinical use and AI-enabled medical devices have increasingly been introduced since the late 2010s and are likely to fundamentally change medical practice across fields^1–3^. In surgery, potential clinical applications of AI include preoperative risk assessment and treatment stratification, intraoperative decision support, and education^4^.

Surgical scene understanding (SSU) refers to the analysis and interpretation of visual and contextual information from a surgical environment, typically using computer vision methods, to identify, for example, surgical tools, critical anatomical landmarks, bleeding, and actions^5^. Clinical SSU applications include real-time assistance in the operating room (OR), for instance, by highlighting hard-to-identify risk structures, or displaying safe dissection zones as part of surgical navigation^6–9^. Surgical procedure recordings underlie most SSU models and pose distinct challenges, such as the presence of involuntary (i.e., breathing, pulse) and voluntary (i.e., targeted instrument manipulation) movement, motion artifacts, reflection artifacts, suboptimal image quality due to blood or smoke in the camera field, anatomical variability, and variability in surgical approaches and techniques. Moreover, for real-time assistance in the OR, AI algorithms must process image data within milliseconds. SSU models need to handle these challenges to tap their full clinical potential. Currently, the use of neural networks in the OR is rare and assistive functions remain the subject of ongoing research.

This systematic review and meta-analysis characterizes the current state-of-the-art and key applications of SSU in minimally invasive abdominal surgery, with a specific focus on underlying data, model development and validation techniques, and model performance reporting. Additionally, we summarize clinical applications that SSU research targets and assess the quality of reporting according to reporting guidelines in medical AI. Overall, our findings highlight major gaps in current research, such as heavy reliance on a few small, single-center public datasets, poor reporting quality, limited model validation methods, or minimal focus on clinical translation, and indicate where future efforts in computational SSU should be directed to enable broader adoption of AI in the OR (**Fig. 1**).

**Fig. 1.**
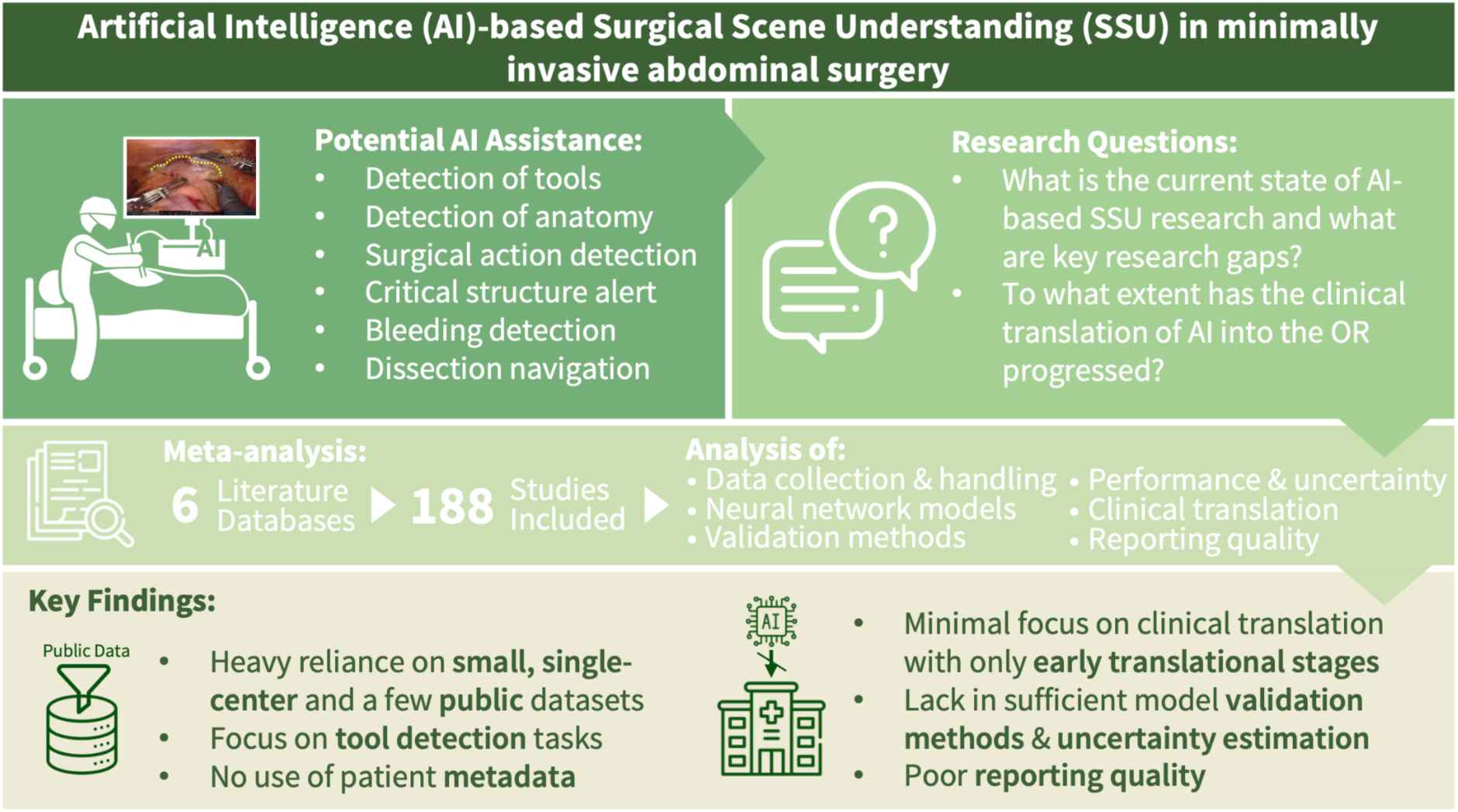
Conceptual overview of this systematic review and meta-analysis.

## Results

The initial literature search yielded 3,011 results. In addition, four publications that were incidentally identified during the literature review but not captured by the search strategy were manually added. Based on title and abstract screening, we selected 338 studies for full-text screening (**Fig. 2**). Of these, we selected 188 studies for data extraction. **Supplementary Tables 3-7** and **Supplementary Figure 1** provide study characteristics and extracted data for all included studies.

**Fig. 2.**
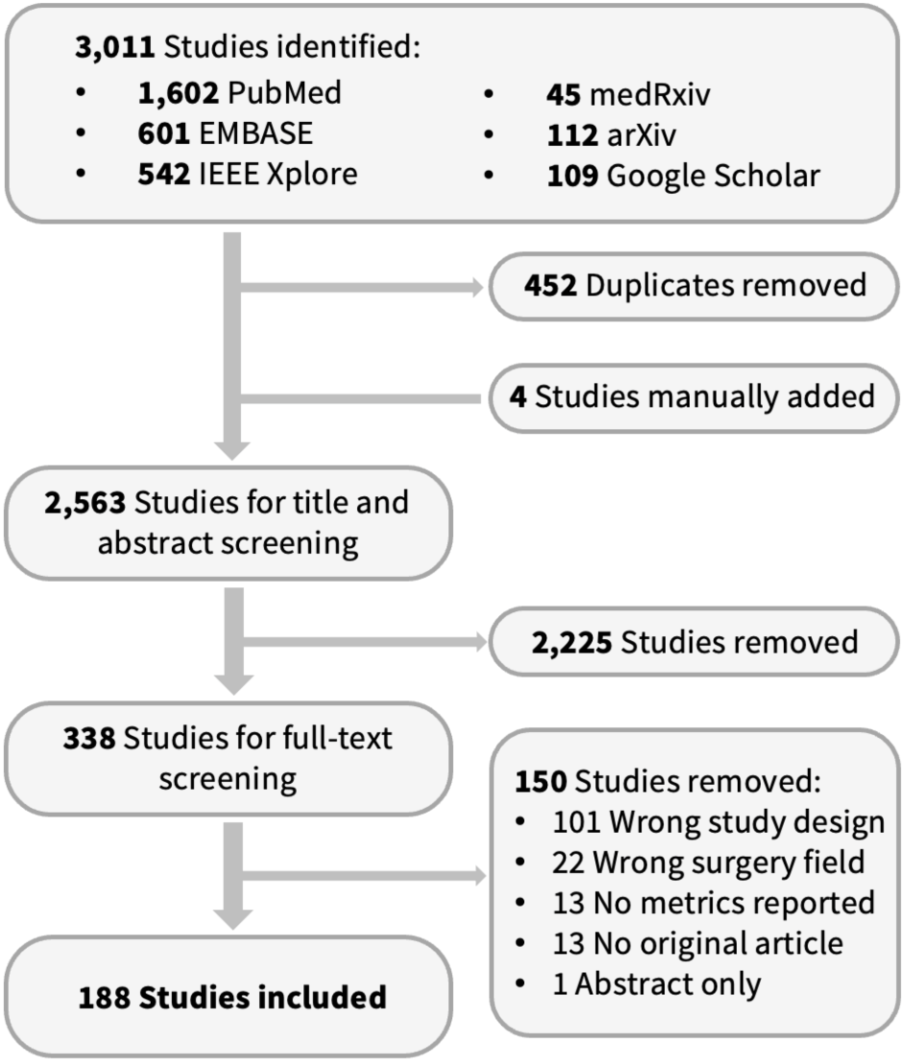
PRISMA flowchart. Flowchart summarizes the inclusion and exclusion of studies at each stage.

### Study characteristics

The annual number of publications on computational SSU has increased rapidly since 2020. Compared to 18 studies published between 2010 and 2019, the number increased to 170 from 2020 to March 2025.

About half of the studies (49.5%, n = 93) were authored exclusively by individuals identified as having non-clinical backgrounds, based on their departmental affiliations and/or academic degrees (i.e., PhD). In contrast, approximately 0.5% (n = 1) of the studies were authored exclusively by individuals with clinical backgrounds. Following journals categorization according to *Clarivate Journal Citation Reports* (*JCR*) Categories^10^, 20.7% (n = 39) of the publications appeared in journals that exclusively publish clinical content, whereas 42.9% (n = 79) were published in interdisciplinary venues and 37.2% (n = 70) were published in technical journals.

### Prediction targets

SSU applications in abdominal surgery are most commonly reported in the fields gastrointestinal surgery (86.2%, n = 162), urology (11.7%, n = 22), and gynecology (9.6%, n = 18). The most frequently analyzed procedures were laparoscopic cholecystectomy (59.0%, n = 111), robotic prostatectomy (9.6%, n = 18), laparoscopic and robotic rectal resection (each 6.4%, n = 12). All surgical procedures included in the analysis are listed in **Supplementary Table 3**. In 8.0% (n = 15) of included works, the specific abdominal surgical procedure was not identifiable. Most works focused exclusively on AI-based instrument detection (56.9%, n = 107) or organ and tissue detection (12.8%, n = 24). The remaining studies (30.3%) used AI to identify bleeding (0.5%, n = 1) or combined multiple detection targets (29.8%, n = 56).

### Data sources

Of all included studies, 83.0% (n = 156) used data from conventional laparoscopic surgeries, while the data underlying 23.4% of included studies (n = 44) originated from robotic surgeries, and for 2.1% (n = 4), the data source was unclear. For the training and validation of neural network models, real patient data were predominantly used (83.5%, n = 157). A smaller proportion of the studies further included non-human (15.4%, n = 29) or synthetically generated (3.7%, n = 7) data. Most of the studies used publicly available datasets (55.3%, n = 104) only, whereas 30.9% (n = 58) relied on privately generated data, and 4.8% (n = 9) of studies included data from unclear sources. A combination of public and private datasets was used in 9.0% (n = 17) of the studies.

Among the publicly available datasets, *Cholec80*^11^ and its derivatives (i.e. *CholecT40*^12^, *CholecT45* and *CholecT50*^13^, *CholecSeg8k*^14^) were most frequently used (56.2%, n = 68). Other commonly used datasets were *m2cai16*^15^ (partially based on *Cholec80*) and its derivatives (14.0%, n = 17), *Endoscapes*^16^ and its derivatives (7.4%, n = 9), *ROBUST-MIS*^17^ (subset of HeiCo^18^; 7.4%, n = 9), the *DSAD*^19^ (5.0%, n = 6), as well as *HeiCo*^18^ and *PSI-AVA*^5^ (each 4.1%, n = 5, **Fig. 3a**).

**Fig. 3.**
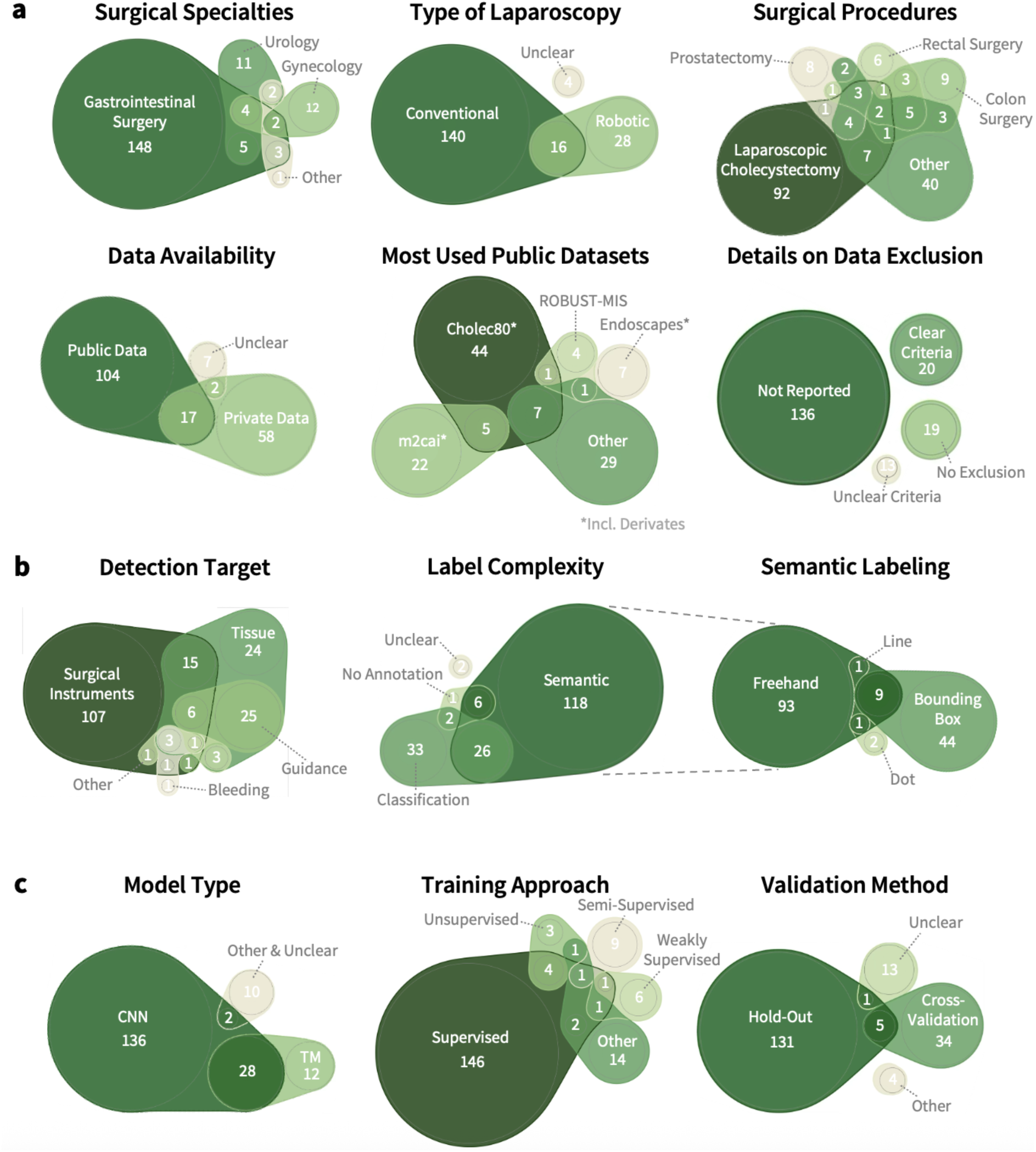
Distribution of data acquisition (a), data labeling (b) and modelling (c) characteristics of surgical scene understanding research. The circle area corresponds to the absolute frequency of respective characteristics, with overlapping regions indicating studies with several characteristics. Numbers within each section represent the exact count of studies. *Abbreviations: CNN = Convolutional Neural Network, TM = Transformer Model*.

### Data selection and labeling

The minority of SSU publications (10.6%, n = 20) provided detailed information on data exclusion, for example, due to poor visibility of the target structure, excessive smoke, or a soiled camera lens. Most studies (72.3%, n = 136) did not address data selection at all, 10.1% (n = 19) of studies explicitly mentioned that no data were excluded, and in 6.9% (n = 13) of included studies, data were reportedly excluded, yet based on unclear criteria.

Most included studies (79.8%, n = 150) used spatially annotated data. This includes freehand or polygon annotations (69.3%, n = 104), bounding box annotations (35.3%, n = 53), and labeling with lines or dots (1.3% or 2.0%, n = 2 or 3). Binary classifications of object presence were used in 32.4% (n = 61) of all studies. Smaller proportions of included studies employed unsupervised learning approaches on unlabeled data (4.8%, n = 9) or provided no information about data labeling (1.1%, n = 2, **Fig. 3b**).

### Model architectures and training approaches

Convolutional neural networks (CNNs) were the most commonly employed AI model architectures (88.3%, n = 166), including advanced versions, such as R-CNNs and Mask R-CNNs. Transformer Models (TM) were used in 21.3% (n = 40) of all works, either alone (6.4%, n = 12) or in combination or comparison with CNNs (14.9%, n = 28, **Fig. 3c**). Temporal aspects were considered in 27.1% (n = 51) of all studies, i.e. through methods like Optical Flow or (Spatial) Temporal Convolution Networks ((SP)-TCN).

SSU models were mostly trained with supervised learning methods (82.4%, n = 155). Other approaches, such as semi-supervised (5.3%, n = 10), weakly supervised (4.3%, n = 8), self-supervised (2.1%, n = 4), or unsupervised learning (4.3%, n = 8), were less common. In 7.4% (n = 14) of studies, the training method was unclear or other techniques were used. One publication incorporated an additional imaging modality (Indocyanine green fluorescence) in the AI application.

### Model validation strategies

The majority of studies (72.9%, n = 137) used hold-out strategies for model validation. Cross-validation approaches were used in 20.7% (n = 39) of SSU studies. Among these, five-fold cross-validation was the most common (44.7%, n = 17), followed by ten-fold (21.1%, n = 8) and four-fold (13.2%, n = 5) validation. In 7.4% (n = 14) of all studies, the validation method was not specified (**Fig. 3c**).

In 12.2% (n = 23) of the included studies, it was not clearly stated whether entirely new data, independent from the training set, were used for model validation or how data-splitting was conducted. In the remaining studies, the model was explicitly tested on previously unseen data, most commonly through data-splitting methods. In 58.5% (n = 109) of the studies, data splitting was performed at the patient level, ensuring that data from patients in the test set were not included during model training. In contrast, 29.8% (n = 56) applied splitting at the frame level, implying potential data leakage (i.e., data from one patient being included in both the training and validation set).

Additionally, we assessed the type of model validation performed, distinguishing between internal retrospective (training and validation using the same data source, 82.4%, n = 155), external retrospective (validation using data from other centers or data sources, 10.1%, n = 19), prospective observational (validation using newly collected data, i.e., from prospectively recorded surgeries, 5.9%, n = 11), and prospective interventional (validation during real-time interventions, 5.3%, n = 10).

### Dataset size and variability

The median number of frames per study used for the development of the AI application was 3,600 (minimum: 157, maximum: 901,000, **Fig. 4a**). When the number of videos per study was reported, the median number was 65 (minimum: 2, maximum: 1,955, **Fig. 4b**). In 13.3% (n = 25) of all studies, no clear statement regarding dataset size was provided. The median number of patients analyzed in the included studies was 40 (minimum: 1, maximum: 2,200, **Fig. 4c**). In 16.0% (n = 30) of included studies, no information on patient counts was provided.

**Fig. 4.**
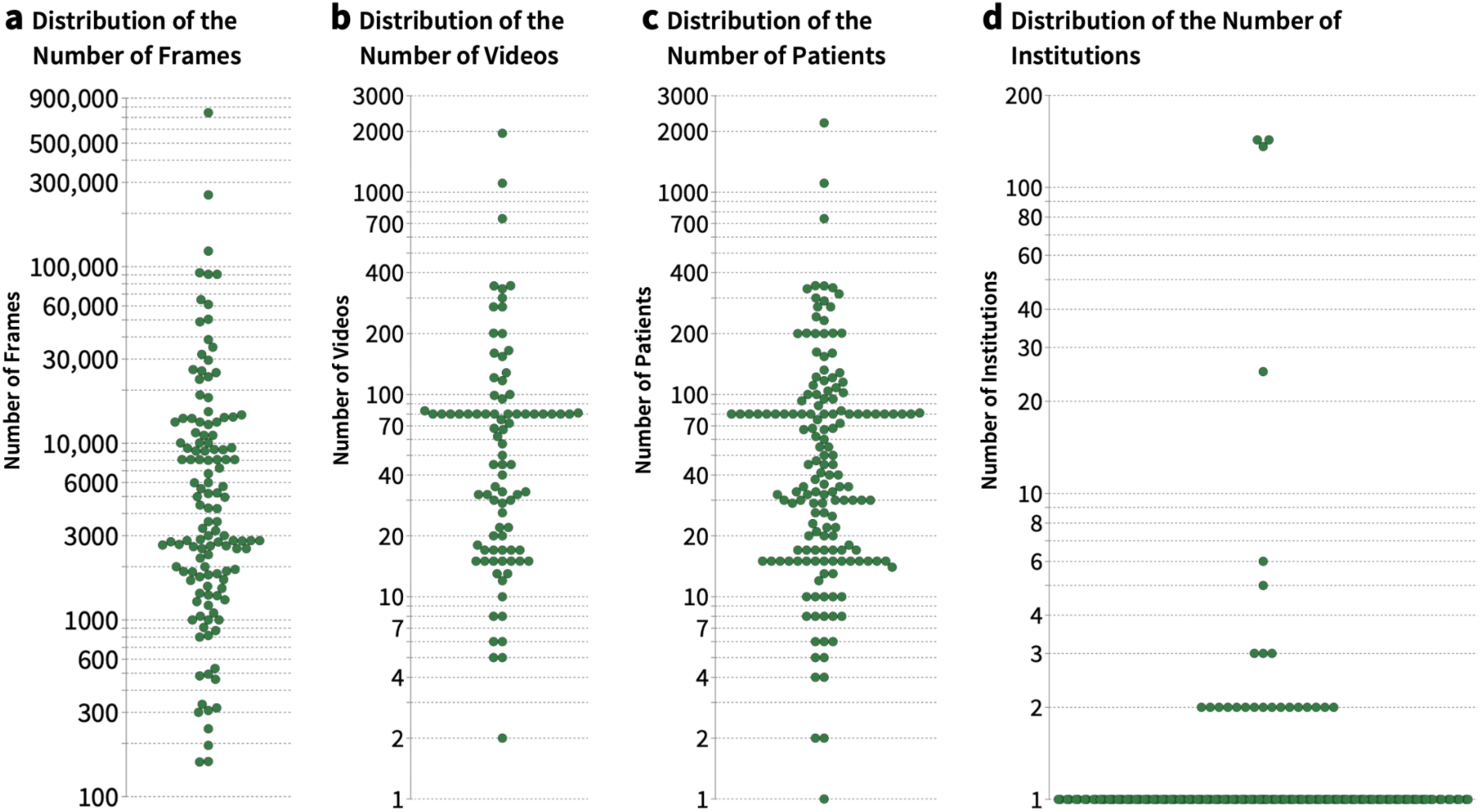
Distribution of dataset sizes, the number of included patients and institutions across surgical scene understanding studies. **a** The dataset size based on the number of frames, **b** the dataset size based on the number of videos, **c** the number of patients included in the studies and **d** number of contributing institutions. Publications were plotted when dataset characteristics were explicitly reported.

Patient metadata, such as age, sex, or surgery indication, were reported in 17.3% (n = 14) of included studies, typically when in-house datasets were generated. The vast majority of studies relied on data from a single institution (70.7%, n = 133, **Fig. 4d**). In 16.0% (n = 30) of included studies, the number of institutions was unclear.

### Reporting of model performance and model uncertainty

When model performance was reported for semantic segmentation tasks, the most commonly used object detection metrics were Intersection over Union (IoU, 43.3%, n = 65), Dice Similarity Coefficient (DSC, 37.3%, n = 56), Precision (27.3%, n = 41), and Recall (27.3%, n = 41). For classification tasks, mean Average Precision (mAP, 45.9%, n = 28), Accuracy (29.5%, n = 18), Recall (29.5%, n = 18), and Precision (27.9%, n = 17) were most commonly applied. Reported results for the most commonly used object detection metrics were extracted and are presented in **Supplementary Table 5**.

In 34.6% (n = 65) of the studies, metrics were aggregated per frame across the entire test set without accounting for individual patients, whereas in 59.6% (n = 112), data splitting was also performed at the patient level. In 4.8% (n = 9), aggregation was performed within individual samples (i.e., all frames of one patient or video) before summarizing across all patients in the test set. For cross-validation, metrics were either aggregated within samples of each fold or calculated per frame and aggregated per fold (each 2.7%, n = 5). In most cases (57.4%, n = 108), the aggregation method was not clearly specified. Temporal metrics, such as the Temporal Consistency Score (TCS), were reported in three publications (1.6%).

Qualitative visualizations of results were presented in 75.5% (n = 142) of studies, most often as an overlay of segmentation on the raw image. 15.4% (n = 29) of the publications did not report qualitative results. Other common visualizations were heat maps, probability maps or class activation maps. Exploratory evaluations of real-time operation capabilities were carried out in 48.9% (n = 92) of SSU studies (i.e., analysis of inference times). When reported (n = 73), the average model inference time was 69.9 ± 110.5 ms. A total of 25 (34.2%) publications with reported inference time reported inference times below 25 frames per second, which would facilitate real-time operation.

Model uncertainty and variability were reported in 38.3% (n = 72) of included studies. Among these, 29.8% (n = 56) provided standard deviations for the metrics, 2.1% (n = 4) reported confidence intervals, 1.6% (n = 3) included interquartile ranges, 1.6% (n = 3) presented the standard error, and 5.3% (n = 10) other methods. In 4.8% (n = 9), variability was only visualized in graphs (i.e., error bars). In 8.5% (n = 16) of SSU studies, uncertainties was calculated per frame and aggregated across all frames, ignoring patient-level grouping. In 2.1% (n = 4), aggregation of model uncertainty measures was performed within individual samples (i.e., all frames of one video or patient). For cross-validation, 1.6% (n = 3) aggregated metrics either per sample or per frame within each fold before summarizing across folds. In 22.9% (n = 43), the aggregation method of prediction variability estimates was not specified. Other statistical tests, such as comparisons of model performance or with human expert annotations using frequentist parametric (i.e., t-test), non-parametric (i.e., Mann-Whitney), or other methods, were reported in 12.8% (n = 24) of the included studies.

### Clinical translation

In 5.9% (n = 11) of all studies, the AI application was tested in real-life scenarios during surgeries (**Supplementary Table 6**). Five studies tested the application during laparoscopic cholecystectomies. The remaining studies implemented SSU tools in robot-assisted esophagectomies (n = 1), robotic kidney surgeries (n = 1), laparoscopic gastrectomies (n = 1), partial nephrectomies (n = 1), colorectal surgeries (n = 1), and in one study, the surgery type was not specified. The number of patients on whom the AI models were tested ranged from one to 40. All eleven studies were technical feasibility studies, with endpoints including AI-based landmark detection, recognition of key surgical features related to bleeding, surgical instrument detection, and trocar insertion assistance.

All included works fall into the preclinical (T1) translational stage according to the *Translational Vision Science and Technology* framework^20^.

### Reporting Quality

Two reviewers independently assessed the reporting quality of the included works using key reporting items for studies on AI in medicine^21^. The interrater agreement between both reviewers was substantial to almost perfect^22^ with Cohen’s Kappa values ranging from 0.73 to 1 (**Supplementary Table 7**).

The reporting quality varied substantially across the evaluated categories. While some criteria were consistently and comprehensively reported, such as information about the study design (97.9%), the model type (94.7%), and the prediction problem (62.8%), others demonstrated notable gaps in reporting. For instance, information on the clinical problem (46.8%), clinical goal (38.8%), existing AI applications (83.5%), data preprocessing (76.1%), model validation (81.9%), and model performance (78.7%) was frequently incompletely provided. Particularly poor reporting was observed for information about data selection (not reported in 74.5% of studies), performance errors (not reported in 68.6% of studies), clinical implications (not reported in 53.4% of studies), study limitations (not reported in 55.9% of studies), and code publication or availability (not reported in 70.2% of studies) (**Fig. 5a**, **Supplementary Table 7**).

**Fig. 5.**
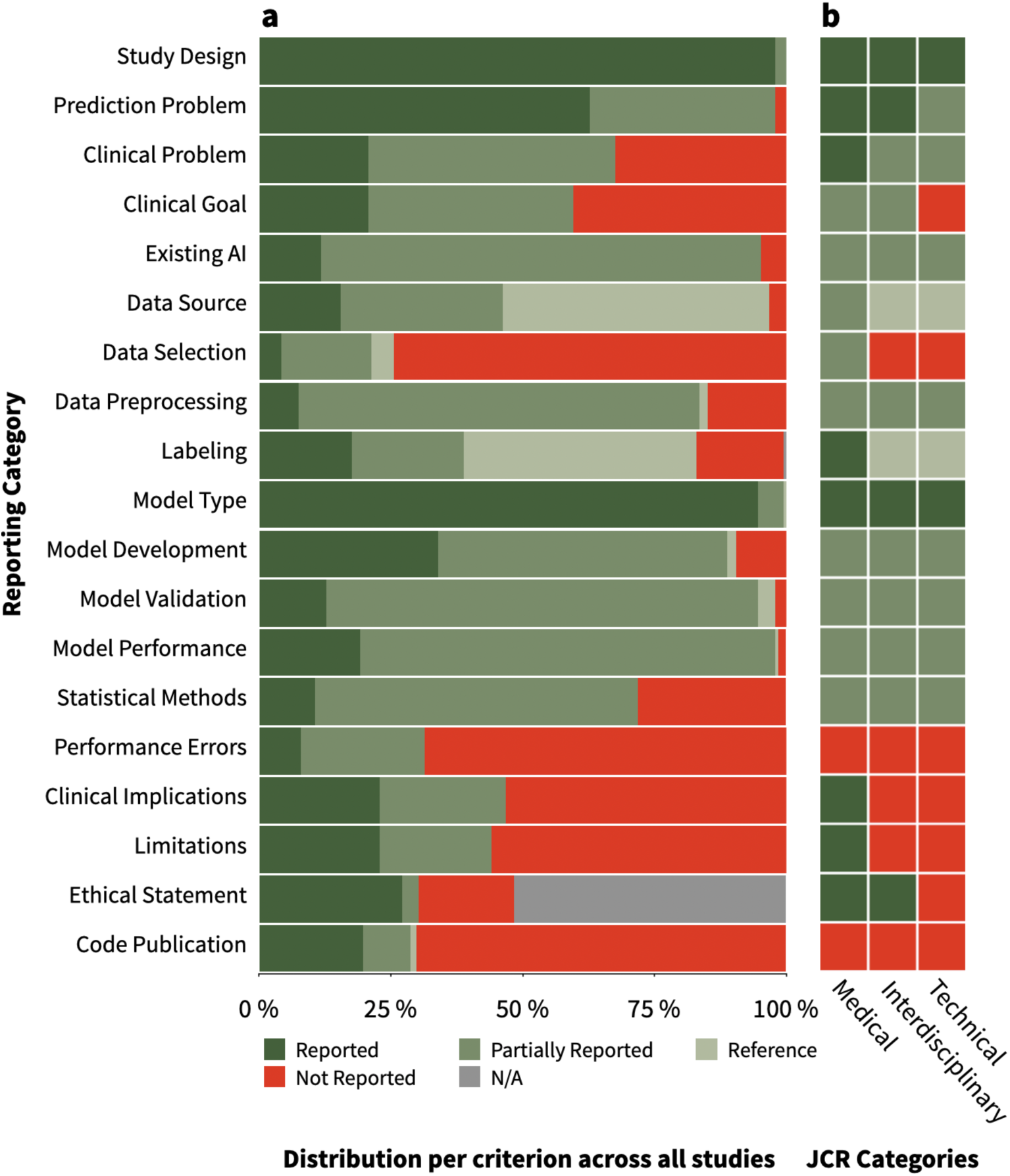
Reporting quality of publications on computational surgical scene understanding. **a** Bars indicate the reporting quality distribution for each reporting criterion, evaluated across all included studies. **b** Heatmap illustrating the median rating per quality criterion across journals, grouped by their *Journal Citation Reports* (*JCR*) groups: (1) medical, (2) interdisciplinary (medical & technical), and (3) technical journals. The criterion “Ethical Statement” was included in the heatmap based on all applicable studies where reporting of an ethics statement would be appropriate. (N/A = Not Applicable)

We compared reporting quality across *JCR* journal groups^10^. Of the included studies, 20.7% were published in medical journals (n = 39), 42.0% in interdisciplinary journals (n = 79), and 37.2% in technical journals (n = 70). We observed significant differences in the reporting quality across these journal categories, particularly between medical journals and the other two *JCR* groups. Specifically, medical journals had substantially higher reporting quality for clinical reporting characteristics, such as the description of the clinical problem and clinical goal, with highly significant differences in pairwise comparisons (p < 0.001). Similar patterns were found for data origin and selection, data labeling, clinical implications, and discussion of study limitations, where reporting in medical journals was consistently more complete (p < 0.001, **Fig. 5b**, **Supplementary Table 8**, **Supplementary Figure 2**).

## Discussion

AI-based SSU is an active field of research that holds potential to support surgeons’ intraoperative decision-making and improve patient outcomes. Despite this promise, the clinical translation of such algorithms remains rare and the clinical relevance of SSU remains to be evaluated.

Our systematic review and reporting quality meta-analysis of 188 studies identified consequential methodological and conceptual limitations in the current SSU research landscape. Key shortcomings are related to the underlying data, the limited clinical translation and relevance of prediction tasks, and the use of suboptimal validation strategies and metrics that poorly reflect an AI model’s clinical potential.

Most works are based on single-center datasets of fewer than 100 patients for which no patient metadata, such as demographic or disease-related parameters, are typically available. Our results indicate that one publicly available single-center dataset of laparoscopic cholecystectomy videos, *Cholec80*^11^, underlies the majority of published SSU research. Other frequently used datasets like *m2cai*^15^ or *Endoscapes*^16^ also originate from laparoscopic cholecystectomies. While many works rely on custom-curated datasets that facilitate new prediction tasks, these datasets are often not published alongside research results. This narrow representation of surgical procedures in publicly available data strongly shapes the research landscape, and most published works in the field of SSU focus on laparoscopic cholecystectomies and evaluate instrument recognition tasks of limited clinical relevance. Of the six most common abdominal surgical procedures in the US in 2018/19 (cholecystectomy, colectomy, gastrointestinal adhesiolysis, fallopian tube ligation and excision, and appendectomy^23^), only two - cholecystectomy and colectomy - are part of the current SSU research landscape. This reflects the limited diversity of SSU research and suggests that the field has been driven more by data availability than by clinical needs.

The need for multicenter data for the development of robust clinical AI tools has been widely acknowledged by the research community across clinical fields^24–27^. Single-center datasets without patient metadata, as are commonly used in SSU research, not only limit the analysis of model generalizability across different disease contexts and institutions, but also hinder the identification of biases, i.e., related to demographic factors, medical history, different medical standards across institutions with different surgical and recording equipment, and variability in surgical techniques. In addition, datasets without patient metadata do not facilitate connecting SSU with patient-level outcomes, such as establishing a relationship between instrument-tissue interactions and surgical complications. Overall, these common data-related practices in the SSU community lead to limited model reproducibility, an extremely limited breadth of use cases, and an overall indeterminate pathway to clinical relevance for computational SSU and intraoperative computer vision applications.

Current SSU research is characterized by a disconnect from clinical practice in that it largely focuses on prediction tasks of limited or indirect clinical relevance and makes marginal progress toward clinical translation. We observed a trend towards studies of limited clinical relevance being published in technical journals, which tend to differ from works published in medical journals in reporting quality, particularly regarding clinical aspects, data labeling, and discussion of study limitations. This trend likely stems from a predominant focus within the technical research community on methodological innovation, such as developing novel model architectures or improving performance on established benchmark datasets^28–30^. Consequently, the primary output of many studies is the demonstration of technical feasibility, often on solely descriptive tasks like instrument detection or anatomical landmark identification, rather than the exploration of clinical utility or a clear pathway to clinical translation. At present, all studies on AI-based SSU fall into the earliest, preclinical stages of translation (T1). The path to clinical translation is further obstructed by barriers, including unresolved ethical and liability questions, strong data protection requirements, and the technical challenges of real-time processing, hardware constraints and seamless integration into complex OR workflows and existing devices^21^. To bridge this gap, a shift in research priorities is necessary. This requires not only the more stringent adoption of AI-specific reporting guidelines to improve transparency and reproducibility, but also the cultivation of sustainable interdisciplinary collaborations between technical and clinical researchers.

The evaluation of SSU models is often undermined by suboptimal validation strategies, the use of performance metrics lacking clinical context, and the absence of model uncertainty estimation. Commonly, evaluations rely on simple hold-out validation without any testing on external datasets or any relation with clinical endpoints or human performance in the validation process. Validation of computational models on a hold-out test set can introduce bias, as performance heavily depends on the chosen data split. Cross-validation, although more computationally intensive, reduces this bias and improves generalizability, which is particularly important for the small datasets that are common in SSU research^31,32^. With regard to the selection of performance metrics, AI-based SSU poses specific challenges, as standard performance metrics often fail to capture clinical relevance. While each metric has distinct strengths, clinical applications require evaluation methods that go beyond standard tasks like classification or spatial localization. Models must demonstrate whether they can reliably highlight delicate structures, such as critical blood vessels or fine nerves, and whether their predictions correlate with critical surgical decisions, for example, identifying safe dissection planes or predicting outcomes based on certain incisions^33^. Moreover, the estimation of model uncertainty, which is critical for establishing trust and safety in high-risk clinical applications, is neglected in most SSU publications. Without transparent evaluation of a model’s confidence, it is difficult to assess its suitability for real-world deployment where errors can directly impact patient safety^34,35^.

Our systematic review has limitations. First, although we applied a broad and systematic search strategy, it is possible that some relevant studies were not identified, for example due to alternative terminology for surgical procedures or AI applications, or a focus on tasks not explicitly captured by our keywords. Second, we excluded studies focusing solely on surgical skill classification and phase recognition that did not include detection or segmentation tasks of objects from the surgical scene. As the boundaries between these two fields and SSU are not clearly defined, this may have led to the omission of relevant studies. Despite these limitations, the present literature review and reporting quality meta-analysis represents the most comprehensive evaluation of SSU publications in the scientific literature to date. Our detailed analysis of reporting quality documents dramatic quality limitations that likely hamper the accessibility of AI methods for surgical patient care.

To address research challenges highlighted by our results, the surgical data science community will need to fundamentally change practices related to data, model reporting, and clinical translation. While data recording has become more standardized at medical institutions^36^, links to patient outcomes and metadata are necessary to increase the clinical relevance of computational SSU. Similarly, dataset publication, along with more transparent reporting of model development methodologies and validation procedures, is needed to increase the reproducibility of published research. While these aspects per se will increase the translational potential of SSU, real-world evaluation in patient care and surgical education settings are key steps towards quantifying their clinical impact on patient- and caregiver-related outcomes.

The past decade of SSU research has focused on developing technical solutions for challenging temporo-spatial prediction tasks that surgical video data pose. In terms of clinical applications, research has largely followed data availability rather than addressing clinically pressing needs. To enable meaningful progress, the focus must now shift toward the identification of clinically relevant tasks and proactively gathering diverse, high-quality, and representative data and metadata to address them. Additionally, suboptimal and non-standardized validation practices, a persistent disconnect between technical performance and clinical utility, and the absence of clear ethical, regulatory, and clinical guidelines are key gaps that must be addressed to bring SSU research closer to clinical impact.

## Methods

This systematic review and meta-analysis adheres to the Preferred Reporting Items for Systematic Reviews and Meta-Analyses (PRISMA) reporting guidelines (**Supplementary Table 1**)^37^. The study was prospectively registered in the International Prospective Register of Systematic Reviews (PROSPERO) database on March 7, 2025 (CRD420251005301).

### Ethics statement

This systematic review exclusively analyzed publicly available publications. No studies involving humans or animals were conducted by the authors for this article.

### Search strategy

Six literature databases (*PubMed*, *EMBASE*, *IEEE Xplore*, *Google Scholar*, *medRxiv* and *arXiv*) covering the interdisciplinary literature on AI-based SSU were searched on October 1, 2024, for publications on AI-based SSU in the field of abdominal surgery published on or after January 1, 2010. **Supplementary Table 2** presents the complete search strategy and search terms.

The screening process was conducted using the *Covidence* platform (*Covidence systematic review software*, *Veritas Health Innovation*, Melbourne, Australia, https://www.covidence.org/). Duplicate studies were removed both automatically and manually. A team of five independent raters carried out study selection: three physician-scientists with at least five years of experience in computational SSU (M.C., D.A.H., F.R.K.), an engineering researcher with three years of experience in surgical and endoscopic video analysis (Z.Z.), and a medical student with two years of experience in computational SSU (S.V.). The study selection process comprised two stages: A first selection stage based on title and abstract, and a second selection stage based on the full text. During each selection stage, each study was screened by two raters, with arbitration of a third rater in case of disagreement.

### Inclusion and exclusion criteria

For inclusion, studies had to meet all of the following criteria: (1) Studies analyzing intraoperative data from abdominal minimally invasive surgeries (including gynecological and urological surgeries) in living humans, (2) development of computational models aimed at SSU (identification and/or localization of tissues/objects visible in surgical images and/or tracking their movement over time, i.e., keypoint tracking or action recognition), (3) reporting of trainable models with a formal validation procedure (i.e., training and test set, cross-validation), reporting of model performance metrics, (4) original articles, (5) published on or after January 1, 2010 and (6) full-text available in English. Publications that combined human and non-human data were included.

Publications were excluded if any of the following criteria applied: (1) Studies analyzing data from open surgeries, radiological surgeries, and extra-abdominal surgeries, (2) surgeries performed only in animals and cadaver studies, (3) studies that do not report computational models with trainable methodologies (i.e., those without any described training or validation methodology) or that do not report any model performance metrics, (4) studies that solely focus on skill classification or phase recognition without spatial scene analysis and (5) reviews, commentaries, editorials, conference abstracts or PhD theses.

### Data extraction

Data extraction was conducted by one physician-scientist with five years of experience in computational SSU (M.C.) using a predefined data extraction table, summarizing the year of publication, authors’ field of expertise (based on their affiliations), *Journal Citation Reports* (*JCR*) Categories of journals^10^, characteristics of the underlying datasets, neural network model architectures, as well as details on model training, validation, and performance reporting. For the underlying datasets, the extracted items included surgical field and surgeries used, type of minimally invasive surgery (robotic vs. conventional laparoscopic surgery), data source and availability, data exclusion criteria, data labeling methodology, dataset size, number of patients, and number of institutions involved. Regarding the neural network model architectures, extracted information comprised model architecture, training approach, validation methodologies, and distribution (technical and clinical), temporal aspects of data usage, temporal performance metrics, additional imaging modalities, performance metrics, uncertainty estimation techniques, visual performance reporting, and assessment of real-time feasibility.

Additionally, we examined the translational stage, i.e., whether the AI application had been evaluated in a clinical patient care setting. This was categorized based on the T stage of *Translational Vision Science and Technology* as first described by Zarbin^20^. T1 research spans from the development of concepts from basic research to early clinical trials. T2 research establishes safety and efficacy in humans. T3 research centers on dissemination and implementation research. T4 research assesses the effectiveness of interventions at the population level. All extracted data are reported in **Supplementary Tables 3-7**.

### Reporting quality assessment

Since no suitable reporting guidelines for AI research in SSU exist, we used key reporting criteria identified in a systematic review of clinical AI reporting guidelines to evaluate the reporting quality for each included publication (**Tab. 1**)^21^. Each criterion was independently rated for each publication by two experts (M.C. and S.V.) using the categories: "Reported", "Partially Reported", "Not Reported", "Not Applicable", or "Reference to Another Publication". In cases of disagreement, a third expert (F.R.K.) reviewed the criterion and made the final decision. For statistical analysis, inter-rater reliability was assessed using Cohen’s Kappa to measure the agreement between the two independent raters.

**Table 1.** Criteria for evaluating the reporting quality.

| Topic | Category | Reporting question |
| --- | --- | --- |
| **Clinical Rationale** | Study Design | Is the study design reported? |
|  | Prediction Problem | Is the prediction problem/target reported? |
|  | Clinical Problem | Are details about the clinical problem and the intended use of the AI application reported? |
|  | Clinical Goal | Is the relation between a prediction problem and the clinical goal reported? |
|  | Existing AI | Are already existing related AI applications mentioned including performance metrics, level of translation or clinical applications? |
| **Data** | Data Sources | Is the data source, the data type and the data structure reported? |
|  | Data Selection | Are inclusion and exclusion criteria for the data/patient collection reported? |
|  | Data Preprocessing | Is it reported, how the data was preprocessed (data transformation, handling of missing data/outliers) |
|  | Labeling | Is it reported how the labeling of input data took place? (number and expertise of labelers) |
| **Model Training and Validation** | Model Type | Is the type of algorithm reported? |
|  | Model Development | Are details about the model development reported? (identification and removal of redundant independent variables, model training, selection strategy) |
|  | Model Validation | Is it reported how the model validation took place? (internal vs external vs cross & validation metrics) |
|  | Model Performance | Is the model performance reported and interpreted? (metrics, confidence intervals) |
|  | Statistical Methods | Are appropriate methods and significance levels for performance comparison of baseline and proposed model reported? |
|  | Performance Errors | Are performance errors reported and analyzed? |
| **Critical Appraisal** | Clinical Implications | Are potential applications of the proposed approach in a real-life clinical setting reported? |
|  | Limitations | Are limitations (bias, generalizability, interpretation pitfalls) of the AI application mentioned? |
| **Ethics and Reproducibility** | Ethical Statement | If necessary: are details on the IRB approval and informed consent procedure reported? |
|  | Code Publication | Is there a statement on public availability of the code reported? |

For downstream analysis, journals were categorized into three groups based on *JCR*^10^ categories: medical journals, interdisciplinary journals (medical and technical), and technical journals (primarily engineering, biomedical engineering, robotics, or computer science). The distribution of reporting quality ratings was compared between journal groups using the Kruskal-Wallis test, followed by Dunn’s post hoc tests with Bonferroni correction for pairwise comparisons.

## Reporting summary

Further information on research design is available in the Nature Research Reporting Summary linked to this article.

## Data availability

All included publications are publicly available. The data that was extracted and analyzed in this systematic review is published along with this work (**Supplementary Tables 3-7**). No custom code was developed for this study.

## Acknowledgements

The authors gratefully acknowledge advisory support regarding model uncertainty estimation from Olivier Colliot (Centre national de la recherche scientifique (CNRS), Paris, France) and Evangelia Christodoulou (German Cancer Research Center (DKFZ), Heidelberg, Germany). D.A.H. received support from the American Surgical Association Foundation and the University of Pennsylvania Linda Pechenik Montague Award for this work. F.R.K. receives support from the German Cancer Research Center (CoBot 2.0), the Joachim Herz Foundation (Add-On Fellowship for Interdisciplinary Life Science), the Central Indiana Corporate Partnership AnalytiXIN Initiative, the Evan and Sue Ann Werling Pancreatic Cancer Research Fund, and the Indiana Clinical and Translational Sciences Institute (EPAR4157) funded, in part, by Grant Number UM1TR004402 from the National Institutes of Health, National Center for Advancing Translational Sciences, Clinical and Translational Sciences Award. The content is solely the responsibility of the authors and does not necessarily represent the official views of the National Institutes of Health. Part of this work was funded by Helmholtz Imaging (HI), a platform of the Helmholtz Incubator on Information and Data Science.

## Author contributions

F.R.K., A.R., and M.C. conceptualized the study. F.R.K., Z.Z., and D.A.H. developed the search strategy. M.C., S.V., Z.Z., I.B., D.A.H., and F.R.K. conducted the review and curated the data. M.C., S.V., and F.R.K. analyzed and interpreted the data. M.C. and F.R.K. prepared visualizations. F.R.K. provided oversight, mentorship, and funding. M.C. and F.R.K. wrote the original draft of the manuscript. All authors critically reviewed, revised, and approved the final version of the manuscript.

## Competing interests

D.A.H. is an independent consultant for Medtronic. F.R.K. declares advisory roles for Radical Healthcare, USA and the Surgical Data Science Collective, USA. The other authors declare no conflicts of interest.

