## Supplementary Material for "Artificial Intelligence for Surgical Scene Understanding: A Systematic Review and Reporting Quality Meta-Analysis"

#### **Supplementary Table 1**: PRISMA Checklist

| Section and Topic | Item # | Checklist item | Location where item is reported |
|---|---|---|---|
| **TITLE** | | | |
| Title | 1 | Identify the report as a systematic review. | Page 1 |
| **ABSTRACT** | | | |
| Abstract | 2 | See the PRISMA 2020 for Abstracts checklist. | Page 2 |
| **INTRODUCTION** | | | |
| Rationale | 3 | Describe the rationale for the review in the context of existing knowledge. | Pages 2-3 |
| Objectives | 4 | Provide an explicit statement of the objective(s) or question(s) the review addresses. | Pages 2-3 |
| **METHODS** | | | |
| Eligibility criteria | 5 | Specify the inclusion and exclusion criteria for the review and how studies were grouped for the syntheses. | Page 15-16 |
| Information sources | 6 | Specify all databases, registers, websites, organisations, reference lists and other sources searched or consulted to identify studies. Specify the date when each source was last searched or consulted. | Page 15 |
| Search strategy | 7 | Present the full search strategies for all databases, registers and websites, including any filters and limits used. | Suppl. Tab. 2 |
| Selection process | 8 | Specify the methods used to decide whether a study met the inclusion criteria of the review, including how many reviewers screened each record and each report retrieved, whether they worked independently, and if applicable, details of automation tools used in the process. | Page 15 |
| Data collection process | 9 | Specify the methods used to collect data from reports, including how many reviewers collected data from each report, whether they worked independently, any processes for obtaining or confirming data from study investigators, and if applicable, details of automation tools used in the process. | Page 15 |
| Data items | 10a | List and define all outcomes for which data were sought. Specify whether all results that were compatible with each outcome domain in each study were sought (e.g. for all measures, time points, analyses), and if not, the methods used to decide which results to collect. | Pages 16-17 |
| | 10b | List and define all other variables for which data were sought (e.g. participant and intervention characteristics, funding sources). Describe any assumptions made about any missing or unclear information. | Pages 16-17 |
| Study risk of bias assessment | 11 | Specify the methods used to assess risk of bias in the included studies, including details of the tool(s) used, how many reviewers assessed each study and whether they worked independently, and if applicable, details of automation tools used in the process. | Pages 15, 17 |
| Effect measures | 12 | Specify for each outcome the effect measure(s) (e.g. risk ratio, mean difference) used in the synthesis or presentation of results. | Page 17 |
| Synthesis methods | 13a | Describe the processes used to decide which studies were eligible for each synthesis (e.g. tabulating the study intervention characteristics and comparing against the planned groups for each synthesis (item #5)). | Pages 15-17 |
| | 13b | Describe any methods required to prepare the data for presentation or synthesis, such as handling of missing summary statistics, or data conversions. | Page 16 |
| | 13c | Describe any methods used to tabulate or visually display results of individual studies and syntheses. | Page 16 |
| | 13d | Describe any methods used to synthesize results and provide a rationale for the choice(s). If meta-analysis was performed, describe the model(s), method(s) to identify the presence and extent of statistical heterogeneity, and software package(s) used. | Pages 16-17 |
| | 13e | Describe any methods used to explore possible causes of heterogeneity among study results (e.g. subgroup analysis, meta-regression). | Pages 16-17 |

| Section and Topic | Item # | Checklist item | Location where item is reported |
|---|---|---|---|
| | 13f | Describe any sensitivity analyses conducted to assess robustness of the synthesized results. | Page 16-17 |
| Reporting bias assessment | 14 | Describe any methods used to assess risk of bias due to missing results in a synthesis (arising from reporting biases). | Page 16-17 |
| Certainty assessment | 15 | Describe any methods used to assess certainty (or confidence) in the body of evidence for an outcome. | Page 16-17 |
| **RESULTS** | | | |
| Study selection | 16a | Describe the results of the search and selection process, from the number of records identified in the search to the number of studies included in the review, ideally using a flow diagram. | Page 4, Fig. 1 |
| | 16b | Cite studies that might appear to meet the inclusion criteria, but which were excluded, and explain why they were excluded. | N/A |
| Study characteristics | 17 | Cite each included study and present its characteristics. | Suppl. Tab. 3-7 |
| Risk of bias in studies | 18 | Present assessments of risk of bias for each included study. | Page 10, Suppl. Tab. 8 |
| Results of individual studies | 19 | For all outcomes, present, for each study: (a) summary statistics for each group (where appropriate) and (b) an effect estimate and its precision (e.g. confidence/credible interval), ideally using structured tables or plots. | Fig 3, 4, 5, Suppl. Tab. 3-7 |
| Results of syntheses | 20a | For each synthesis, briefly summarise the characteristics and risk of bias among contributing studies. | Fig. 3-5, Supp. Fig. 1-2, Pages 4-10 |
| | 20b | Present results of all statistical syntheses conducted. If meta-analysis was done, present for each the summary estimate and its precision (e.g. confidence/credible interval) and measures of statistical heterogeneity. If comparing groups, describe the direction of the effect. | Fig. 3-5, Supp. Fig. 1-2, Pages 9-10 |
| | 20c | Present results of all investigations of possible causes of heterogeneity among study results. | N/A |
| | 20d | Present results of all sensitivity analyses conducted to assess the robustness of the synthesized results. | N/A |
| Reporting biases | 21 | Present assessments of risk of bias due to missing results (arising from reporting biases) for each synthesis assessed. | N/A |
| Certainty of evidence | 22 | Present assessments of certainty (or confidence) in the body of evidence for each outcome assessed. | Suppl. Tab. 8 & Fig. 2 |
| **DISCUSSION** | | | |
| Discussion | 23a | Provide a general interpretation of the results in the context of other evidence. | Pages 12-14 |
| | 23b | Discuss any limitations of the evidence included in the review. | Page 13 |
| | 23c | Discuss any limitations of the review processes used. | Page 13 |
| | 23d | Discuss implications of the results for practice, policy, and future research. | Pages 12-14 |
| **OTHER INFORMATION** | | | |
| Registration and protocol | 24a | Provide registration information for the review, including register name and registration number, or state that the review was not registered. | Page 15 |
| | 24b | Indicate where the review protocol can be accessed, or state that a protocol was not prepared. | Page 15 |
| | 24c | Describe and explain any amendments to information provided at registration or in the protocol. | Page 15 |

| Section and Topic | Item # | Checklist item | Location where item is reported |
|---|---|---|---|
| Support | 25 | Describe sources of financial or non-financial support for the review, and the role of the funders or sponsors in the review. | Page 20 |
| Competing interests | 26 | Declare any competing interests of review authors. | Page 20 |
| Availability of data, code and other materials | 27 | Report which of the following are publicly available and where they can be found: template data collection forms; data extracted from included studies; data used for all analyses; analytic code; any other materials used in the review. | Page 17 |

*From:* Page MJ, McKenzie JE, Bossuyt PM, Boutron I, Hoffmann TC, Mulrow CD, et al. The PRISMA 2020 statement: an updated guideline for reporting systematic reviews. BMJ 2021;372:n71. doi: 10.1136/bmj.n71

**Supplementary Table 2**: Search Strategy

Below is the search strategy for our review. Terms were combined with the Boolean operators AND (to link major concepts) and OR (to capture synonyms/variants). Searches were limited to articles published in English between 1 January 2010 and 1 October 2024.

| Literature Database | Search Terms |
|---|---|
| PubMed | 1. “surg* instrument*”[Title/Abstract] OR “surgical instruments”[MeSH] OR “surg* scene*”[Title/Abstract] OR “surg* tool*”[Title/Abstract] OR “anatom*”[Title/Abstract] OR “critical view of safety”[Title/Abstract]<br>2. “laparoscopy”[MeSH] OR “video-assisted surgery”[MeSH] OR “surg* video*”[Title/Abstract] OR “surg* imag*”[Title/Abstract] OR “laparoscop*”[Title/Abstract] OR “robot* surg*”[Title/Abstract] OR “minimally invasive”[Title/Abstract]<br>3. “Artificial Intelligence”[MeSH] OR “artificial intelligence”[Title/Abstract] OR “deep learning”[Title/Abstract] OR “machine learning”[Title/Abstract] OR “neural network*”[Title/Abstract] OR “segmentation”[Title/Abstract] OR “classification”[Title/Abstract] OR “computer vision”[Title/Abstract]<br>4. Final PubMed set: 1 AND 2 AND 3. |
| EMBASE | 1. “surg* instrument*”:ti,ab OR “surgical equipment”:de OR “surg* scene*”:ti,ab OR “surg* tool*”:ti,ab OR “anatom*”:ti,ab<br>2. “laparoscopy”:de OR “laparoscopy”:ti,ab OR “video-assisted surgery”:de OR “surg* video*”:ti,ab OR “surg* imag*”:ti,ab OR “laparoscop*”:ti,ab OR “robot* surg*”:ti,ab OR “minimally invasive”:ti,ab<br>3. “artificial intelligence”:de OR “artificial intelligence”:ti,ab OR “deep learning”:ti,ab OR “machine learning”:ti,ab OR “neural network*”:ti,ab OR “computer vision”:ti,ab OR “segmentation”:ti,ab<br>4. Final EMBASE set: 1 AND 2 AND 3. |
| IEEE Xplore | 1. “surg* instrument*” OR “surg* scene*” OR “surg* tool*” OR “anatom*” OR “critical view of safety”<br>2. “laparoscopy” OR “video-assisted surgery” OR “surg* video*” OR “surg* imag*” OR “laparoscop*” OR “robot* surg*” OR “minimally invasive”<br>3. “artificial intelligence” OR “deep learning” OR “machine learning” OR “neural network*” OR “segmentation” OR “classification” OR “computer vision”<br>4. Final IEEE Xplore: 1 AND 2 AND 3. |
| Google Scholar | “scene understanding” AND “surgical video” AND “artificial intelligence” |
| arXiv | 1. “surgical instruments” OR “surgical scene” OR “surgical tool” OR anatomy OR “critical view of safety”<br>2. “laparoscopy” OR “video-assisted surgery” OR “surgical video” OR “surgical image” OR “robot surgery” OR “minimally invasive”<br>3. “artificial intelligence” OR “deep learning” OR “machine learning” OR “neural network” OR “segmentation” OR “classification” OR “computer vision”<br>4. Final Arxiv: 1 AND 2 AND 3. |
| medRxiv / bioRxiv | “surgical video” AND (“scene” OR “anatomy”) AND “artificial intelligence” |

#### **Supplementary Table 3**: Studies and study characteristics included in the systematic review and meta-analysis (1)

The following table presents an overview of all included studies, detailing key characteristics across four main categories: **Publication Characteristics, Medical Field, Data, and Model Characteristics**.

| Publication Characteristics | | | Medical Field | | Data | | | | | | | | Model Characteristics | | |
|---|---|---|---|---|---|---|---|---|---|---|---|---|---|---|---|
| **First author, year [reference]** | **Clarivate Journal Citation Reports Categories[a]** | **Authors background** | **Application field** | **Surgeries used** | **Proposed detection** | **Human (real) only or additional data** | **Data Origin** | **Dataset name(s)[b]** | **Annotation type** | **Annotation tool** | **Exclution of images containing artifacts** | **What was excluded** | **Model type** | **Temporal aspects** | **Training approach** |
| Acharya, 2022 [1] | Engineering Conference Paper | Computer Science | Gynecological Surgery | Laparoscopic Endometriosis | Organs/Tissue | Real | Public dataset(s) | GLENDA | Classification | unclear | Unclear | n/a | CNN | not reported | Supervised |
| Al Hajj, 2018 [2] | Radiology, Nuclear Medicine & Medical Imaging; Engineering, Biomedical; Computer Science, Interdisciplinary Applications; Computer Science, Artificial Intelligence | Interdisciplinary | Visceral / General Surgery | Laparoscopic cholecystectomy | Surgical Instruments | Real | Public dataset(s) | Cholec80 | Classification | unclear | No | n/a | CNN | "(CNNs) whose outputs are fed to recurrent neural networks (RNNs) in order to take temporal relationships between events into account" | Supervised |
| Alapatt, 2021 [3] | Preprint | Interdisciplinary | Visceral / General Surgery | Laparoscopic cholecystectomy | Organs/Tissue, Surgical Instruments, Technical guidance | Real | Private dataset(s), Public dataset(s) | Endoscapes | Semantic | unclear | Unclear | n/a | CNN | Temporally Constrained Neural Network (TCNN) | Semi-supervised |
| Ali, 2022 [4] | Preprint | Computer Science | Visceral / General Surgery | Laparoscopic cholecystectomy | Surgical Instruments | Real | Public dataset(s) | m2cai-tl | Bounding box | unclear | Unclear | n/a | CNN | not reported | Semi-supervised |
| Alkhamaiseh, 2023 [5] | Surgery | Interdisciplinary | Visceral / General Surgery | Laparoscopic cholecystectomy | Organs/Tissue, Technical guidance | Real | Private dataset(s), Public dataset(s) | open source videos | Semantic | Fiji plugin (i.e., annotator) | Yes (clear criteria) | "14 videos were excluded from initial modelling because of poor image quality due to either bleeding or less visible or obscured landmarks, anatomical differences or misses, or procedures that do not follow the general standard of cholecystectomy." | CNN | not reported | Supervised |
| Alshirbaji, 2021 [6] | Engineering, Biomedical | Computer Science | Visceral / General Surgery | Laparoscopic cholecystectomy | Surgical Instruments | Real | Public dataset(s) | Cholec80 | Classification | unclear | Unclear | n/a | CNN | STM-clip (sequence-to-one configuration) | Supervised |
| Alshirbaji, 2021 [7] | Engineering, Biomedical | Computer Science | Visceral / General Surgery | Laparoscopic cholecystectomy | Surgical Instruments | Real | Public dataset(s) | Cholec80, Cholec20 | Classification | unclear | Unclear | n/a | CNN | not reported | Supervised |
| Alshirbaji, 2018 [8] | Engineering, Biomedical | Computer Science | Visceral / General Surgery | Laparoscopic cholecystectomy | Surgical Instruments | Real | Public dataset(s) | Cholec80 | Classification | unclear | No | n/a | CNN | not reported | Supervised |
| Ángeles-Cerón, 2021 [9] | Engineering Conference Paper | Computer Science | Visceral / General Surgery | Laparoscopic Colectomy, Laparoscopic Rectal Resection, Laparoscopic Sigmoid Resection | Surgical Instruments | Real | Public dataset(s) | HeiCo | Semantic | unclear | Yes (clear criteria) | "996 frames with no visible instruments" | CNN | not reported | Supervised |
| Ángeles-Cerón, 2022 [10] | Radiology, Nuclear Medicine & Medical Imaging; Engineering, Biomedical; Computer Science, Interdisciplinary Applications; Computer Science, Artificial Intelligence | Computer Science | Visceral / General Surgery | Laparoscopic Colectomy, Laparoscopic Rectal Resection, Laparoscopic Sigmoid Resection | Surgical Instruments | Real | Public dataset(s) | ROBUST-MIS | Semantic | Understand.ai | Yes (clear criteria) | "17% empty frames(ef) on its trainingset. These frames do not have any visible instruments in them, and although we could have left them as negative examples for training,we remove them from the trainingset." | CNN | not reported | Supervised |
| Aoyama, 2024 [11] | Surgery | Interdisciplinary | Visceral / General Surgery | Laparoscopic Gastrectomy | Organs/Tissue, Technical guidance | Real | Private dataset(s) | n/a | Semantic | unclear | Unclear | n/a | CNN | not reported | Supervised |
| Arabian, 2022 [12] | Engineering, Biomedical | Computer Science | Visceral / General Surgery | Laparoscopic cholecystectomy | Surgical Instruments | Real | Public dataset(s) | Cholec80 | Classification | unclear | Unclear | n/a | CNN | not reported | Supervised |
| Arabian, 2023 [13] | Chemistry, Analytical; Instruments & Instrumentation; Engineering, Electrical & Electronic | Computer Science | Visceral / General Surgery | Laparoscopic cholecystectomy | Surgical Instruments | Real, Also non-human data included | Public dataset(s) | Cholec80 | Classification, Bounding box | unclear | Unclear | n/a | CNN | not reported | Supervised |
| Aspart, 2022 [14] | Surgery; Engineering, Biomedical; Radiology, Nuclear Medicine & Medical Imaging | Interdisciplinary | Visceral / General Surgery | Laparoscopic cholecystectomy | Surgical Instruments | Real | Private dataset(s) | n/a | Classification | unclear | Unclear | n/a | CNN | not reported | Supervised |
| Attia, 2017 [15] | Engineering Conference Paper | Computer Science | Visceral / General Surgery | Robotic Colorectal Surgery | Surgical Instruments | Real, Also non-human data included | Public dataset(s) | EndoVis15 | Semantic | unclear | Unclear | n/a | CNN | RNN | Supervised |
| Ayobi, 2024 [16] | Preprint | Interdisciplinary | Urological Surgery | Robotic Prostatectomy | Surgical Instruments | Real | Private dataset(s) | GraSP | Semantic, Classification | Label-Studio, Toronto Annotation Suite | Unclear | n/a | CNN, TM | "we employ MViT as our video feature extractor to capture intricate details across various space-time scales by leveraging transformers." | Supervised |
| Bai, 2023 [17] | Preprint | Computer Science | Visceral / General Surgery | Laparoscopic cholecystectomy | Surgical Instruments | Real, Also non-human data included | Public dataset(s) | m2cai-t | Bounding box | unclear | Unclear | n/a | CNN | not reported | Supervised |
| Bakker, 2024 [18] | Urology & Nephrology | Interdisciplinary | Urological Surgery, Visceral / General Surgery | Robotic Prostatectomy, Robotic Rectal Resection | Organs/Tissue, Others | Real | Private dataset(s), Public dataset(s) | DSAD | Semantic | LabelMe, 3DSlicer | Yes (unclear criteria) | "if their intraoperative videos did not include the urethral dissection phase or if the image quality was too poor. And clearly visible and unobstructed parts of structures were annotated" | CNN | not reported | Supervised |

| | | | | | | | | | | | | | | | |
|---|---|---|---|---|---|---|---|---|---|---|---|---|---|---|---|
| Bamba, 2021 [19] | Surgery; Engineering, Biomedical; Radiology, Nuclear Medicine & Medical Imaging | Medicine | Visceral / General Surgery | Laparoscopic Colectomy, Laparoscopic Rectal Resection, Laparoscopic Hernia Repair, Laparoscopic Sigmoid Resection | Organs/Tissue, Surgical Instruments, Bleeding | Real | Private dataset(s) | n/a | Semantic | unclear | Unclear | n/a | CNN | not reported | Supervised |
| Bamba, 2021 [20] | Multidisciplinary Sciences | Interdisciplinary | Visceral / General Surgery | Laparoscopic Colectomy, Laparoscopic Rectal Resection, Laparoscopic Hernia Repair, Laparoscopic Sigmoid Resection | Surgical Instruments | Real | Private dataset(s) | n/a | Semantic | unclear | Unclear | n/a | CNN | not reported | Supervised |
| Ban, 2023 [21] | Preprint | Interdisciplinary | Visceral / General Surgery | Laparoscopic cholecystectomy | Organs/Tissue, Technical guidance | Real | Public dataset(s) | CholecT45 | Classification | unclear | Unclear | n/a | CNN, TM | not reported | Supervised |
| Batić, 2024 [22] | Surgery; Engineering, Biomedical; Radiology, Nuclear Medicine & Medical Imaging | Computer Science | Visceral / General Surgery, Gynecological Surgery, Urological Surgery | Robotic Prostatectomy, Rectal Resection, Robotic Gastrectomy, Robotic Rectal Resection, Laparoscopic Gynecological Surgery, Laparoscopic cholecystectomy | Organs/Tissue, Surgical Instruments | Real | Public dataset(s) | ESAD, LapGyn4, Surgical Actions160, GLENDA, hsDB-instrument, HeiCo, PSI-AVA, DSAD, Cholec80, CholecT45, CholecSeg8k | Semantic | different | Yes (clear criteria) | "all synthetic images are excluded" | CNN, TM | not reported | Unsupervised, Supervised |
| Batić, 2023 [23] | Preprint | Computer Science | Visceral / General Surgery, Urological Surgery, Gynecological Surgery | Robotic Prostatectomy, Rectal Resection, Robotic Gastrectomy, Robotic Rectal Resection, Laparoscopic Gynecological Surgery, Laparoscopic cholecystectomy | Organs/Tissue, Surgical Instruments | Real | Public dataset(s) | ESAD, LapGyn4, Surgical Actions160, GLENDA, hsDB-instrument, HeiCo, PSI-AVA, DSAD, Cholec80, CholecT45, CholecSeg8k | Classification, No annotation | unclear | Unclear | n/a | CNN, TM | Multi-Stage Temporal Convolutional Network (MS-TCN) (for phase recognition) | Unsupervised, Supervised |
| Boonkong, 2022 [24] | Engineering Conference Paper | Interdisciplinary | Gynecological Surgery | Laparoscopic Gynecological Surgery | Surgical Instruments | Real | Private dataset(s) | n/a | Unclear | unclear | Unclear | n/a | CNN | not reported | Others/unclear |
| Brandenburg, 2023 [25] | Surgery | Interdisciplinary | Visceral / General Surgery | Robotic Esophagectomy | Organs/Tissue, Surgical Instruments, Bleeding, Others | Real | Private dataset(s) | n/a | Classification | CVAT | No | n/a | CNN | not reported | Active |
| Casella, 2021 [26] | Engineering Conference Paper | Computer Science | Urological Surgery | Robotic Partial Nephrectomy | Organs/Tissue | Real | Public dataset(s) | Nephrec9 | Semantic | unclear | Yes (clear criteria) | "Frames with heavy motion blur (due to quick changes in camera position) and with largely occluded vessels were removed" | CNN | 3D Fully-Convolutional Neural Network | Supervised |
| Chen, 2024 [27] | Engineering, Biomedical; Radiology, Nuclear Medicine & Medical Imaging; Engineering, Electrical & Electronic; Imaging Science & Photographic Technology; Computer Science, Interdisciplinary Applications | Interdisciplinary | Visceral / General Surgery | Laparoscopic cholecystectomy, Robotic Hepato-Pancreatic-Biliary Surgery | Organs/Tissue, Surgical Instruments | Real, Also non-human data included | Private dataset(s), Public dataset(s) | CholecSeg8k | No annotation, Semantic | unclear | Unclear | n/a | TM | not reported | Self-supervised |
| Chen, 2013 [28] | Engineering Conference Paper | Interdisciplinary | Others | unclear | Surgical Instruments | Real | Unclear | unclear | No annotation | n/a | Unclear | n/a | Others/Unclear (Spiking Neural Network (SNN)) | not reported | Others/unclear |
| Chen, 2017 [29] | Engineering Conference Paper | Interdisciplinary | Visceral / General Surgery | unclear | Surgical Instruments | Real | Unclear | unclear | Bounding box | unclear | Unclear | n/a | CNN | "spatio-temporal context (STC) learning algorithm for tracking between video frames." | Supervised |
| Choi, 2017 [30] | Engineering Conference Paper | Computer Science | Visceral / General Surgery | Laparoscopic cholecystectomy | Surgical Instruments | Real | Public dataset(s) | m2cai-t | Bounding box, Classification | unclear | Unclear | n/a | CNN | not reported | Supervised |
| Ciaparrone, 2020 [31] | Engineering Conference Paper | Computer Science | Gynecological Surgery | unclear | Surgical Instruments | Real | Unclear | n/a | Semantic | VGG Image Annotator | No | "We chose to keep those noisy frames, as they can be useful to analyze if the models are able to generalize on unseen conditions and unfiltered noisy frames." | CNN | not reported | Supervised |
| Colleoni, 2024 [32] | Radiology, Nuclear Medicine & Medical Imaging; Engineering, Biomedical; Computer Science, Interdisciplinary Applications; Computer Science, Artificial Intelligence | Computer Science | Visceral / General Surgery, Urological Surgery | Laparoscopic cholecystectomy, Robotic Partial Nephrectomy, Robotic Prostatectomy | Organs/Tissue | Real, Synthetic | Public dataset(s), Private dataset(s) | CholecSeg8k | Semantic | unclear | Unclear | n/a | CNN, TM | not reported | Others/unclear |
| Colleoni, 2022 [33] | Engineering, Biomedical; Radiology, Nuclear Medicine & Medical Imaging; Engineering, | Computer Science | Urological Surgery | Robotic Prostatectomy | Surgical Instruments | Real, Synthetic, Also non-human data included | Public dataset(s), | RARP45 | No annotation, Semantic | unclear | Unclear | n/a | CNN | not reported | Others/unclear |

| | | | | | | | | | | | | | | | |
|---|---|---|---|---|---|---|---|---|---|---|---|---|---|---|---|
| | Electrical & Electronic; Imaging Science & Photographic Technology; Computer Science, Interdisciplinary Applications | | | | | | Private dataset(s) | | | | | | | | |
| Daneshgar Rahbar, 2023 [34] | Imaging Science & Photographic Technology | Computer Science | Visceral / General Surgery, Others | Robotic Lobectomy, Robotic Prostatectomy | Surgical Instruments | Real, Also non-human data included | Public dataset(s) | US NLM | Semantic | unclear | Unclear | n/a | CNN | not reported | Supervised |
| Davila, 2023 [35] | Engineering Conference Paper | Computer Science | Visceral / General Surgery | Laparoscopic cholecystectomy | Surgical Instruments | Real, Also non-human data included | Public dataset(s) | Cholec80 | Classification | unclear | Unclear | n/a | CNN | not reported | Supervised |
| De Backer, 2023 [36] | Urology & Nephrology | Interdisciplinary | Urological Surgery | Robotic Partial Nephrectomy | Surgical Instruments | Real | Private dataset(s) | n/a | Semantic | SuperAnnotate | Unclear | n/a | CNN | not reported | Supervised |
| den Boer, 2023 [37] | Surgery | Interdisciplinary | Visceral / General Surgery | Robotic Esophagectomy | Organs/Tissue | Real | Private dataset(s) | n/a | Semantic | LabelMe | Yes (clear criteria) | "Lymphatic or fatty tissue was excluded in the annotation of the anatomy." | CNN | not reported | Supervised |
| Derathé, 2020 [38] | Surgery; Engineering, Biomedical; Radiology, Nuclear Medicine & Medical Imaging | Interdisciplinary | Visceral / General Surgery | Laparoscopic Sleeve Gastrectomy | Surgical Instruments, Organs/Tissue, Technical guidance | Real | Private dataset(s) | n/a | Classification, Semantic | Surgery Workflow Toolbox, CamiTK | Unclear | n/a | Others/Unclear (Linear Discriminant Analysis (LDA); Support Vector Machine (SVM)) | not reported | Supervised |
| Du, 2019 [39] | Radiology, Nuclear Medicine & Medical Imaging; Engineering, Biomedical; Computer Science, Interdisciplinary Applications; Computer Science, Artificial Intelligence | Computer Science | Visceral / General Surgery | Robotic Colorectal Surgery | Surgical Instruments | Real, Also non-human data included | Public dataset(s) | EndoVis Conventional Laparoscopic Instrument Dataset | Bounding box | unclear | No | n/a | Others/Unclear | not reported | Others/unclear |
| Endo, 2023 [40] | Surgery | Interdisciplinary | Visceral / General Surgery | Laparoscopic cholecystectomy | Organs/Tissue, Technical guidance | Real | Private dataset(s) | n/a | Semantic | unclear | Yes (clear criteria) | "We excluded LC cases with severe inflammation and abnormal biliary anatomy because it was difficult to annotate anatomical landmarks" | CNN | not reported | Supervised |
| Fernandez-Rodríguez, 2024 [41] | Preprint | Computer Science | Visceral / General Surgery | Laparoscopic cholecystectomy | Surgical Instruments | Real | Public dataset(s) | Cholec80, CholecSeg8k | Classification, Semantic | PixelAnnotaion Tool | Unclear | n/a | CNN | OpticalFlow | Supervised |
| Fuentes-Hurtado, 2019 [42] | Surgery; Engineering, Biomedical; Radiology, Nuclear Medicine & Medical Imaging | Computer Science | Visceral / General Surgery | Robotic Colorectal Surgery, Laparoscopic Sleeve Gastrectomy, Laparoscopic Gastric Bypass | Surgical Instruments | Real | Public dataset(s), Private dataset(s) | EndoVis15 | Line, Semantic | unclear | Unclear | n/a | CNN | not reported | Supervised, Weakly-supervised |
| Fujinaga, 2023 [43] | Surgery | Interdisciplinary | Visceral / General Surgery | Laparoscopic cholecystectomy | Organs/Tissue, Technical guidance | Real | Private dataset(s) | n/a | Bounding box, Semantic | unclear | Yes (unclear criteria) | "with bad anatomical visibility" | CNN | not reported | Others/unclear |
| Ghamsarian, 2024 [44] | Surgery; Engineering, Biomedical; Radiology, Nuclear Medicine & Medical Imaging | Interdisciplinary | Gynecological Surgery | Laparoscopic Colorectal Surgery, Laparoscopic Endometriosis Surgery | Organs/Tissue | Real | Public dataset(s) | ENID | Semantic | Endoscopic Concept Annotation Tool | Yes (unclear criteria) | | CNN | not reported | Supervised |
| Gitau, 2024 [45] | Engineering Conference Paper | Computer Science | Visceral / General Surgery | Laparoscopic cholecystectomy | Surgical Instruments, Others | Also non-human data included, Real | Public dataset(s) | Cholec80 | Classification | unclear | Unclear | n/a | CNN | not reported | Supervised |
| Grammatikopoulou, 2023 [46] | Surgery; Engineering, Biomedical; Radiology, Nuclear Medicine & Medical Imaging | Computer Science | Urological Surgery, Visceral / General Surgery | Laparoscopic cholecystectomy, Robotic Partial Nephrectomy | Organs/Tissue, Surgical Instruments | Real | Private dataset(s), Public dataset(s) | CholecSeg8k | Semantic | PixelAnnotaion Tool | Unclear | n/a | CNN, TM | "spatio-temporal decoder based on the TCN model to augment any semantic segmentation backbone for Spatial temporal convolutional network (SP-TCN)" | Supervised |
| Guédon, 2021 [47] | Surgery | Interdisciplinary | Visceral / General Surgery, Gynecological Surgery | Laparoscopic cholecystectomy, Laparoscopic Hysterectomy | Surgical Instruments | Real | Private dataset(s) | n/a | Classification | NOUS application | Unclear | n/a | CNN | "temporal smoothing on the automatic recognition of the phases in order to avoid shifting between phases for only a few frames. For instance, as the network only used visual information, a few frames without instruments present in the frames may cause a wrong phase recognition. Therefore, a window of 15 to 30 frames was used for the smoothing on the automatic recognition of the phases." | Supervised |
| Hasan, 2021 [48] | Engineering Conference Paper | Computer Science | Visceral / General Surgery | Robotic Colorectal Surgery | Surgical Instruments | Real, Also non-human data included | Public dataset(s) | EndoVis15 | Semantic | unclear | Unclear | n/a | CNN | not reported | Supervised |
| Huang, 2022 [49] | Robotics; Engineering, Biomedical | Interdisciplinary | Urological Surgery | Robotic Partial Nephrectomy, unclear | Surgical Instruments | Real | Public dataset(s) | n/a | No annotation, Semantic | unclear | Unclear | n/a | CNN | not reported | Unsupervised |

| | | | | | | | | | | | | | | | |
|---|---|---|---|---|---|---|---|---|---|---|---|---|---|---|---|
| Jalal, 2022 [50] | Engineering, Biomedical | Computer Science | Visceral / General Surgery | Laparoscopic cholecystectomy | Surgical Instruments | Real | Public dataset(s) | Cholec80 | Bounding box, Classification | unclear | Yes (clear criteria) | "Images that have no tool or multiple instances of the same tool were excluded" | CNN | not reported | Weakly-supervised |
| Jalal, 2023 [51] | Chemistry, Analytical; Instruments & Instrumentation; Engineering, Electrical & Electronic | Computer Science | Visceral / General Surgery | Laparoscopic cholecystectomy | Surgical Instruments | Real | Public dataset(s) | Cholec80, Cholec80-Boxes | Classification, Bounding box | MATLAB Video Labeler toolbox | No | n/a | CNN | "The CNN-SE-MSF was combined with an LSTM network to model temporal dependencies along the video sequence" | Supervised |
| Jamal, 2024 [52] | Preprint | Computer Science | Urological Surgery, Gynecological Surgery, Visceral / General Surgery | Robotic Prostatectomy, Laparoscopic Hysterectomy, Laparoscopic cholecystectomy | Surgical Instruments | Real, Also non-human data included | Public dataset(s) | SAR-RARP50, AutoLaparo, LapI2I, CholecSeg8k | Semantic, Bounding box | unclear | Unclear | n/a | TM, CNN | not reported | Supervised |
| Jang, 2023 [53] | Engineering Conference Paper | Computer Science | Visceral / General Surgery | unclear | Organs/Tissue | Real | Unclear | unclear | No annotation, Bounding box | unclear | Unclear | n/a | CNN, TM | "forward and backward prediction in conjunction with a Siamese network" | Self-supervised |
| Jaspers, 2024 [54] | Engineering Conference Paper | Interdisciplinary | Visceral / General Surgery, Urological Surgery | Laparoscopic cholecystectomy, Robotic Esophagectomy, Robotic Prostatectomy, Robotic Rectal Resection, Laparoscopic Gynecological Surgery, Laparoscopic Gastric Bypass, Robotic Gastrectomy | Organs/Tissue | Real | Private dataset(s), Public dataset(s) | CholecSeg8k | Semantic | unclear | Unclear | n/a | CNN, TM | not reported | Supervised, Unsupervised |
| Jearanai, 2023 [55] | Surgery | Interdisciplinary | Visceral / General Surgery | Unclear | Organs/Tissue | Real | Private dataset(s) | n/a | Bounding box | Roboflow | Unclear | n/a | CNN | not reported | Supervised |
| Jha, 2021 [56] | Preprint | Computer Science | Visceral / General Surgery | Laparoscopic Colectomy, Laparoscopic Rectal Resection, Laparoscopic Hernia Repair, Laparoscopic Sigmoid Resection | Surgical Instruments | Real | Public dataset(s) | ROBUST-MIS | Semantic | understand.ai | Unclear | n/a | CNN | not reported | Supervised |
| Jin, 2020 [57] | Radiology, Nuclear Medicine & Medical Imaging; Engineering, Biomedical; Computer Science, Interdisciplinary Applications; Computer Science, Artificial Intelligence | Computer Science | Visceral / General Surgery | Laparoscopic cholecystectomy | Surgical Instruments | Real | Public dataset(s) | Cholec80 | Classification | unclear | No | n/a | CNN | RNN (for phase recognition) | Supervised |
| Jin, 2018 [58] | Preprint | Computer Science | Visceral / General Surgery | Laparoscopic cholecystectomy | Surgical Instruments | Real | Public dataset(s) | m2cai-tl | Bounding box, Classification | unclear | Unclear | n/a | CNN | not reported | Supervised |
| Kamrul Hasan, 2021 [59] | Radiology, Nuclear Medicine & Medical Imaging; Engineering, Biomedical; Computer Science, Interdisciplinary Applications; Computer Science, Artificial Intelligence | Computer Science | Gynecological Surgery | Laparoscopic Hysterectomy | Surgical Instruments | Real, Also non-human data included | Private dataset(s) | n/a | Semantic, Line, Dot, Classification | ImageJ | Unclear | n/a | CNN | not reported | Supervised |
| Kanakatte, 2020 [60] | Engineering Conference Paper | Computer Science | Visceral / General Surgery | Laparoscopic cholecystectomy | Surgical Instruments | Real | Public dataset(s) | Cholec80 | Semantic | LabelMe | Unclear | n/a | CNN | Spatio-temporal deep network (ST-LSTM) | Supervised |
| Kawamura, 2023 [61] | Surgery | Interdisciplinary | Visceral / General Surgery | Laparoscopic cholecystectomy | Organs/Tissue, Technical guidance | Real | Private dataset(s) | n/a | Classification | own | Yes (clear criteria) | "excluded patients with severe cholecystitis that did not meet the CVS requirements due to the inability to identify arteries and those forced to proceed to bailout surgery or laparotomy" | CNN | not reported | Supervised |
| Khalid, 2023 [62] | Surgery | Interdisciplinary | Visceral / General Surgery | Laparoscopic cholecystectomy | Organs/Tissue, Technical guidance | Real | Private dataset(s), Unclear | n/a | Semantic | GoNoGoNet | Yes (clear criteria) | "only included if all tool-tissue interactions were deemed to have occurred within the surgeon-assessed Go zone." | CNN | not reported | Others/unclear |
| Khalid, 2023 [63] | Preprint | Computer Science | Visceral / General Surgery | Laparoscopic cholecystectomy, Laparoscopic Appendectomy, Laparoscopic Hernia Repair | Surgical Instruments | Real | Private dataset(s) | unclear | No annotation, Semantic | unclear | Unclear | n/a | Others/Unclear (GNN) | "radiance fields assisted 3D feature extraction" | Self-supervised, Supervised |
| Kim, 2024 [64] | Engineering Conference Paper | Computer Science | Visceral / General Surgery | Laparoscopic cholecystectomy | Surgical Instruments | Real | Public dataset(s) | m2cai-tl | Bounding box | unclear | Unclear | n/a | CNN | not reported | Supervised |
| Kinoshita, 2024 [65] | Surgery | Interdisciplinary | Visceral / General Surgery | Robotic Rectal Resection, Laparoscopic Rectal Resection | Organs/Tissue | Real | Private dataset(s) | n/a | Semantic | unclear | Unclear | n/a | CNN | not reported | Supervised |
| Kitaguchi, 2023 [66] | Surgery | Interdisciplinary | Visceral / General Surgery | Laparoscopic Colorectal Resection | Organs/Tissue | Real | Private dataset(s) | n/a | Semantic | unclear | Yes (clear criteria) | "he study cohort did not include patients with a high BMI or severe intra-abdominal adhesions due to previous surgery" | CNN | not reported | Supervised |
| Kitaguchi, 2022 [67] | Medicine, General & Internal | Interdisciplinary | Visceral / General Surgery | Laparoscopic Colorectal Resection | Surgical Instruments | Real | Private dataset(s) | n/a | Semantic | unclear | Unclear | n/a | CNN | not reported | Supervised |
| Kitaguchi, 2022 [68] | Multidisciplinary Sciences | Interdisciplinary | Visceral / General Surgery | Laparoscopic Colorectal | Surgical Instruments | Real | Private dataset(s) | n/a | Semantic | unclear | Yes (clear criteria) | "out-of-focus images and/or images with mist were excluded" | CNN | not reported | Supervised |

| | | | | | | | | | | | | | | | |
|---|---|---|---|---|---|---|---|---|---|---|---|---|---|---|---|
| | | | | Resection, Laparoscopic Distal Gastrectomy, Laparoscopic cholecystectomy, Laparoscopic Partial Hepatectomy | | | | | | | | | | | |
| Kletz, 2019 [69] | Engineering Conference Paper | Interdisciplinary | Gynecological Surgery | Laparoscopic Gynecological Surgery | Surgical Instruments | Real | Private dataset(s) | n/a | Semantic | unclear | Unclear | n/a | CNN | not reported | Supervised |
| Kolbinger, 2023 [70] | Surgery | Interdisciplinary | Visceral / General Surgery | Robotic Rectal Resection | Organs/Tissue, Technical guidance | Real | Public dataset(s) | DSAD | Semantic, Bounding box | 3DSlicer, CVAT | No | n/a | CNN | not reported | Supervised |
| Kolbinger, 2024 [71] | Oncology; Surgery | Interdisciplinary | Visceral / General Surgery | Robotic Rectal Resection | Organs/Tissue | Real | Private dataset(s) | n/a | Semantic | 3DSlicer | No | n/a | CNN | not reported | Supervised |
| Kolbinger, 2024 [72] | Engineering Conference Paper | Interdisciplinary | Visceral / General Surgery | Robotic Rectal Resection | Organs/Tissue | Real | Public dataset(s) | DSAD | Semantic | 3DSlicer | No | n/a | TM | not reported | Supervised |
| Kondo, 2021 [73] | Engineering, Biomedical | Computer Science | Visceral / General Surgery | Laparoscopic cholecystectomy | Surgical Instruments | Real | Public dataset(s) | Cholec80 | Classification | unclear | No | n/a | TM, CNN | "Transformer architecture to analyse inter-frame correlation" | Supervised |
| Konduri, 2024 [74] | Engineering, Biomedical | Computer Science | Visceral / General Surgery, Gynecological Surgery | Laparoscopic cholecystectomy, Laparoscopic Gynecological Surgery, Laparoscopic Colectomy, Laparoscopic Rectal Resection, Laparoscopic Sigmoid Resection | Surgical Instruments | Real | Public dataset(s) | CholecSeg8k, LapGyn4, HeiCo | Classification | PixelAnnotaion Tool | Unclear | n/a | CNN | not reported | Supervised |
| Kong, 2021 [75] | Surgery; Engineering, Biomedical; Radiology, Nuclear Medicine & Medical Imaging | Computer Science | Gynecological Surgery | Robotic Hysterectomy | Surgical Instruments | Real, Also non-human data included | Private dataset(s) | n/a | Semantic | Labelme | Unclear | n/a | CNN | not reported | Supervised |
| Kumazu, 2021 [76] | Multidisciplinary Sciences | Interdisciplinary | Visceral / General Surgery | Robotic Gastrectomy | Organs/Tissue, Technical guidance | Real | Private dataset(s) | n/a | Semantic | unclear | Unclear | n/a | CNN | not reported | Supervised |
| Kumazu, 2025 [77] | Multidisciplinary Sciences | Interdisciplinary | Visceral / General Surgery | Robotic Gastrectomy, Laparoscopic Gastrectomy, Laparoscopic Colorectal Surgery, Laparoscopic Hernia Repair | Organs/Tissue, Technical guidance | Real | Private dataset(s) | n/a | Semantic | unclear | Yes (clear criteria) | "Conversely, we excluded images in which the LCT was not clearly visible due to bleeding, smoking, or artifacts, and in which the LCT had degenerated due to inflammation or prior treatment. Additionally, we excluded evaluation scenes in which the surgical procedure was not progressing smoothly." | CNN | not reported | Supervised |
| Labrunie, 2022 [78] | Surgery; Engineering, Biomedical; Radiology, Nuclear Medicine & Medical Imaging | Interdisciplinary | Visceral / General Surgery | Laparoscopic Liver Resection | Organs/Tissue | Real | Private dataset(s) | n/a | Semantic | unclear | Unclear | n/a | CNN | not reported | Supervised |
| Lam, 2022 [79] | Robotics; Engineering, Biomedical | Computer Science | Visceral / General Surgery | Laparoscopic Gastric Band Insertion | Surgical Instruments | Real | Private dataset(s) | n/a | Semantic | VGG Image annotator | Unclear | n/a | CNN | Markov-chain | Supervised |
| Laplante, 2022 [80] | Surgery | Interdisciplinary | Visceral / General Surgery | Laparoscopic cholecystectomy | Organs/Tissue, Technical guidance | Real | Private dataset(s) | n/a | Semantic | Think Like A Surgeon | Yes (clear criteria) | "Exclusion criteria included: 1) top-down approach LC, and 2) poor visualization of the hepatocystic triangle whereby the entire triangle was never in full view" | CNN | not reported | Supervised |
| Lavanchy, 2021 [81] | Multidisciplinary Sciences | Interdisciplinary | Visceral / General Surgery | Laparoscopic cholecystectomy | Surgical Instruments | Real | Private dataset(s) | n/a | Bounding box, Classification | unclear | Unclear | n/a | CNN | not reported | Supervised |
| Le, 2023 [82] | Engineering Conference Paper | Computer Science | Visceral / General Surgery | Laparoscopic cholecystectomy | Surgical Instruments | Real | Public dataset(s) | m2cai-tl | Bounding box | unclear | No | n/a | CNN | not reported | Supervised |
| Lee, 2024 [83] | Computer Science, Interdisciplinary Applications; Computer Science, Information Systems; Mathematical & Computational Biology; Medical Informatics | Computer Science | Visceral / General Surgery | Laparoscopic cholecystectomy, Laparoscopic Colectomy, Laparoscopic Rectal Resection, Laparoscopic Sigmoid Resection | Organs/Tissue, Surgical Instruments | Real | Public dataset(s) | CholecSeg8K, ROBUST-MIS | Semantic, Bounding box | PixelAnnotaion Tool, understand.ai | Unclear | n/a | CNN, Others/Unclear (Generative Model (DIPO)) | not reported | Supervised |
| Leifman, 2024 [84] | Computer Science; Artificial Intelligence; Computer Science Applications | Interdisciplinary | Visceral / General Surgery | Laparoscopic cholecystectomy | Organs/Tissue, Technical guidance | Real | Private dataset(s), Public dataset(s) | Cholec80 | Classification, Semantic | unclear | Yes (unclear criteria) | "Eligibility criteria were laparoscopic cholecystectomy for biliary colic or acute and chronic cholecystitis in patients 18 years of age or older." | CNN | "Inspired by the success of the above methods, we propose an approach similar in spirit to DeepCVS, but we add the temporal aggregation stage, and our approach can work in real-time settingsapproach similar in spirit to DeepCVS, but we add the temporal aggregation stage, and our approach can work in real-time settings" | Supervised |
| Leifmann, 2022 [85] | Engineering Conference Paper | Computer Science | Visceral / General Surgery | Laparoscopic Colectomy, Laparoscopic Rectal Resection, Laparoscopic | Surgical Instruments | Synthetic, Real, Also non-human data included | Public dataset(s) | Robotool | Bounding box | Understand.ai | Unclear | n/a | CNN | not reported | Supervised |

| | | | | | | | | | | | | | | | |
|---|---|---|---|---|---|---|---|---|---|---|---|---|---|---|---|
| | | | | Sigmoid Resection, Unclear | | | | | | | | | | | |
| Li, 2025 [86] | Preprint | Interdisciplinary | Urological Surgery, Gynecological Surgery, Visceral / General Surgery | Laparoscopic cholecystectomy, Unclear | Organs/Tissue, Surgical Instruments, Bleeding | Real | Public dataset(s) | CholecSeg8K | Classification, No annotation | unclear | Unclear | n/a | TM | Spatial-temporal fusion layer | Weakly-supervised |
| Li, 2023 [87] | Engineering Conference Paper | Interdisciplinary | Visceral / General Surgery | Laparoscopic cholecystectomy | Organs/Tissue, Technical guidance | Real | Public dataset(s) | CholecT50, Cholec80, open source videos | Semantic | PixelAnnotaion Tool | Unclear | n/a | TM | not reported | Supervised |
| Liao, 2024 [88] | Preprint | Interdisciplinary | Visceral / General Surgery | Laparoscopic cholecystectomy | Surgical Instruments | Real | Public dataset(s) | Cholec80, CholecSeg8k | Semantic | PixelAnnotaion Tool | Unclear | n/a | CNN | "Both modules utilize a spatio-temporal class activation map (ST-CAM) and share input from ViT [4] encoded frames and video-level class labels as the supervision signal." | Weakly-supervised |
| Lin, 2024 [89] | Engineering, Biomedical; Radiology, Nuclear Medicine & Medical Imaging; Engineering, Electrical & Electronic; Imaging Science & Photographic Technology; Computer Science, Interdisciplinary Applications | Interdisciplinary | Visceral / General Surgery | Laparoscopic cholecystectomy | Organs/Tissue, Surgical Instruments | Real | Public dataset(s) | CholecQ | Bounding box, Classification | unclear | Unclear | n/a | CNN | "Global Average Pooling Layer and Long Short-Term Memory (LSTM) Layer to obtain the snippet context features. These obtained snippet context features include temporal information about the key frame and reference frames." | Supervised |
| Lin, 2021 [90] | Robotics | Interdisciplinary | Visceral / General Surgery | Laparoscopic Proctocolectomy | Surgical Instruments | Real, Synthetic | Public dataset(s) | ROBUST-MIS | Semantic | Understand.ai | Yes (unclear criteria) | "As an initial exploration, we evaluated our method only on the proctocolectomy dataset for the binary segmentation task." | CNN | "Multi-frame Feature Aggregation (MFFA) module that aggregates features both temporally and spatially for segmentation" | Others/unclear |
| Liu, 2022 [91] | Surgery | Interdisciplinary | Visceral / General Surgery | Laparoscopic cholecystectomy | Organs/Tissue, Technical guidance | Real | Private dataset(s) | n/a | Bounding box | unclear | Yes (clear criteria) | "excluding videos that were incomplete (n = 1), pixels or format incompatible (n = 17), or gallbladder gangrene (n = 1)" | CNN | not reported | Supervised |
| Liu, 2024 [92] | Engineering, Biomedical; Radiology, Nuclear Medicine & Medical Imaging; Engineering, Electrical & Electronic; Imaging Science & Photographic Technology; Computer Science, Interdisciplinary Applications | Computer Science | Visceral / General Surgery | Robotic Rectal Resection | Surgical Instruments | Real | Private dataset(s) | n/a | Semantic | unclear | Unclear | n/a | CNN, TM | not reported | Supervised |
| Liu, 2022 [93] | Robotics; Engineering, Biomedical | Interdisciplinary | Visceral / General Surgery | Laparoscopic cholecystectomy | Surgical Instruments | Real, Also non-human data included | Public dataset(s) | Cholec80-locations | Bounding box | unclear | Unclear | n/a | CNN | "Given that the input image is a continuous video frame, which is time-dependent, RNN was added to the CNN framework. On the other hand, since the input multi-channel color image is a tensor, the convolutional LSTM structure was used to extract the image's temporal information. At this stage, we formed the DSCNet-CLSTM network, which could utilize the temporal and spatial context information of laparoscopic video frames" | Supervised |
| Lou, 2023 [94] | Engineering, Biomedical; Radiology, Nuclear Medicine & Medical Imaging; Engineering, Electrical & Electronic; Imaging Science & Photographic Technology; Computer Science, Interdisciplinary Applications | Interdisciplinary | Gynecological Surgery | Unclear | Surgical Instruments | Real | Public dataset(s) | ART-NET | Semantic | unclear | Unclear | n/a | CNN | not reported | Semi-supervised |
| Loukas, 2020 [95] | Surgery | Computer Science | Visceral / General Surgery | Laparoscopic cholecystectomy | Organs/Tissue, Others | Real | Public dataset(s) | m2cai-w | Classification | unclear | Unclear | n/a | Others/Unclear (Bayesian gaussian mixture models (VBGMM)) | not reported | Others/unclear, Unsupervised |
| Loza, 2024 [96] | Engineering, Biomedical | Computer Science | Visceral / General Surgery | Laparoscopic cholecystectomy | Surgical Instruments | Real | Public dataset(s) | m2cai-tl | Bounding box | unclear | Unclear | n/a | TM | not reported | Supervised |
| Maack, 2024 [97] | Engineering Conference Paper | Computer Science | Visceral / General Surgery | Robotic Rectal Resection | Organs/Tissue | Real | Public dataset(s) | DSAD | Semantic | 3DSlicer | No | n/a | CNN, TM | not reported | Supervised |

| | | | | | | | | | | | | | | | |
|---|---|---|---|---|---|---|---|---|---|---|---|---|---|---|---|
| Madad Zadeh, 2020 [98] | Surgery | Interdisciplinary | Gynecological Surgery | Laparoscopic Hysterectomy | Organs/Tissue, Surgical Instruments | Real | Public dataset(s), Private dataset(s) | SurgAI | Semantic | Supervisely | Unclear | n/a | CNN | not reported | Supervised |
| Madani, 2022 [99] | Surgery | Interdisciplinary | Visceral / General Surgery | Laparoscopic cholecystectomy | Organs/Tissue, Technical guidance | Real | Public dataset(s) | Cholec80, m2cai-w | Semantic | Think Like A Surgeon | Yes (unclear criteria) | "Frames wherein the camera was outside & 2900 frames were extracted, 273 of which were excluded" | CNN | not reported | Supervised |
| Maqbool, 2020 [100] | Preprint | Computer Science | Visceral / General Surgery | Laparoscopic cholecystectomy | Organs/Tissue, Surgical Instruments, Bleeding | Real | Public dataset(s) | m2cai-t | Semantic | MatLab | Unclear | n/a | CNN | not reported | Supervised |
| Marullo, 2023 [101] | Pharmacology & Pharmacy | Interdisciplinary | Urological Surgery | Robotic Prostatectomy | Surgical Instruments, Bleeding | Real | Private dataset(s) | n/a | Semantic, Classification | unclear | Yes (clear criteria) | "due to poor resolution, or the absence of surgical equipment were dataset, which comprised 318 images. The dataset was then divided into train, validation, removed" | CNN | not reported | Supervised |
| Mascagni, 2022 [102] | Surgery | Interdisciplinary | Visceral / General Surgery | Laparoscopic cholecystectomy | Organs/Tissue, Technical guidance | Real | Private dataset(s) | n/a | Semantic | Pixel Annotation Tool | Yes (unclear criteria) | "After manual screening, 2854 images of CVS were annotated and included in the CVS dataset." | CNN | not reported | Supervised |
| Matsumoto, 2024 [103] | Multidisciplinary Sciences | Interdisciplinary | Visceral / General Surgery | Laparoscopic Gastrectomy | Surgical Instruments | Real | Private dataset(s) | n/a | Semantic | unclear | Unclear | n/a | CNN | not reported | Supervised |
| Mehta, 2024 [104] | Surgery; Engineering, Biomedical; Radiology, Nuclear Medicine & Medical Imaging | Computer Science | Visceral / General Surgery | Laparoscopic cholecystectomy | Organs/Tissue, Technical guidance | Real | Unclear | n/a | Semantic | unclear | Unclear | n/a | TM | not reported | Supervised |
| Mishra, 2017 [105] | Engineering Conference Paper | Computer Science | Visceral / General Surgery | Laparoscopic cholecystectomy | Surgical Instruments | Real | Public dataset(s) | m2cai-t | Classification | unclear | Unclear | n/a | CNN | "long short-term memory (LSTM) on the extracted spatial features of the video sequence to capture the temporal connectionism across deep residual visual features and thereby increase the accuracy in prediction." | Supervised |
| Murali, 2024 [106] | Preprint | Computer Science | Visceral / General Surgery | Laparoscopic cholecystectomy | Organs/Tissue, Surgical Instruments, Technical guidance | Real | Public dataset(s) | Endoscapes-Seg50 | Semantic | unclear | Unclear | n/a | CNN, TM | CycleSelect | Supervised |
| Murali, 2023 [107] | Preprint | Interdisciplinary | Visceral / General Surgery | Laparoscopic cholecystectomy | Organs/Tissue, Technical guidance | Real | Public dataset(s) | Endoscapes+ | Classification, Semantic, Bounding box | unclear | Unclear | n/a | CNN, TM | "latent spatiotemporal graph representations of entire surgical videos, with each node representing a surgical tool or anatomical structure and edges representing relationships between nodes across space and time." | Supervised |
| Murali, 2022 [108] | Engineering, Biomedical; Radiology, Nuclear Medicine & Medical Imaging; Engineering, Electrical & Electronic; Imaging Science & Photographic Technology; Computer Science, Interdisciplinary Applications | Interdisciplinary | Visceral / General Surgery | Laparoscopic cholecystectomy | Organs/Tissue, Technical guidance, Surgical Instruments | Real | Public dataset(s) | DeepCVS, Endoscapes+ | Classification, Semantic, Bounding box | unclear | Unclear | n/a | CNN, TM | not reported | Semi-supervised |
| Myo, 2024 [109] | Telecommunications; Computer Science, Information Systems; Engineering, Electrical & Electronic | Interdisciplinary | Visceral / General Surgery | Laparoscopic Colectomy, Laparoscopic Rectal Resection, Laparoscopic Sigmoid Resection | Surgical Instruments | Real | Public dataset(s) | ROBUST-MIS | Semantic | Roboflow online annotation tool | No | n/a | CNN | "Modified Y+BT algorithm consists of YOLOv8 as the core segmentation and ByteTrack as an adapted module. Two confidence thresholds (high and low) have to be defined for ByteTrack in order to take advantage of having 2 associations between the current frame and the previous frame" | Supervised |
| Nakanuma, 2023 [110] | Surgery | Interdisciplinary | Visceral / General Surgery | Laparoscopic cholecystectomy | Organs/Tissue, Technical guidance | Real | Private dataset(s) | n/a | Semantic | unclear | Unclear | n/a | CNN | not reported | Supervised |
| Namazi, 2022 [111] | Surgery | Interdisciplinary | Visceral / General Surgery | Laparoscopic cholecystectomy | Surgical Instruments | Real | Public dataset(s) | m2cai-td, Cholec80 | Classification | unclear | Unclear | n/a | CNN | not reported | Supervised |
| Nema , 2023 [112] | Surgery | Computer Science | Visceral / General Surgery | Robotic Colorectal Surgery | Surgical Instruments | Real, Also non-human data included, Synthetic | Public dataset(s) | EndoVis15 | Semantic | unclear | No | n/a | CNN, Others/Unclear (Generative Adversarial Network (GAN)) | not reported | Supervised |
| Nwoye, 2024 [113] | Preprint | Computer Science | Visceral / General Surgery | Laparoscopic cholecystectomy | Surgical Instruments | Real | Public dataset(s) | CholecTrack20 | Bounding box | unclear | Unclear | n/a | CNN | Multi-Class Multi-Object Tracking (MC-MOT) | Weakly-supervised, Self-supervised, Supervised |

| | | | | | | | | | | | | | | | |
|---|---|---|---|---|---|---|---|---|---|---|---|---|---|---|---|
| Nwoye, 2019 [114] | Surgery; Engineering, Biomedical; Radiology, Nuclear Medicine & Medical Imaging | Computer Science | Visceral / General Surgery | Laparoscopic cholecystectomy | Surgical Instruments | Real | Public dataset(s) | Cholec80 | Classification, Bounding box | unclear | Unclear | n/a | CNN | "As temporal model, we propose to use a recurrent neural network (RNN), with the aim to determine the current position of each tool from the input feature map along with information from prior images captured in RNN's state." | Weakly-supervised |
| Oh, 2024 [115] | Multidisciplinary Sciences | Interdisciplinary | Visceral / General Surgery | Laparoscopic Hepatectomy | Organs/Tissue | Real | Private dataset(s) | n/a | Semantic | CVAT | Yes (clear criteria) | "Frames with obscured fields due to smoke, completely obscured biliary structures by surgical instruments, or camera positioned outside the surgical field were excluded." | CNN | not reported | Supervised |
| Owen, 2022 [116] | Surgery; Engineering, Biomedical; Radiology, Nuclear Medicine & Medical Imaging | Computer Science | Visceral / General Surgery | Laparoscopic cholecystectomy | Organs/Tissue, Technical guidance | Real | Private dataset(s) | unclear | Semantic | unclear | Unclear | n/a | CNN | not reported | Semi-supervised |
| Ozbulak, 2025 [117] | Preprint | Interdisciplinary | Visceral / General Surgery | Laparoscopic cholecystectomy | Organs/Tissue, Surgical Instruments | Real | Public dataset(s) | CholecSeg8k | Semantic | PixelAnnotaion Tool | Unclear | n/a | TM | not reported | Others/unclear |
| Pan, 2023 [118] | Surgery; Engineering, Biomedical; Radiology, Nuclear Medicine & Medical Imaging | Interdisciplinary | Visceral / General Surgery | Laparoscopic cholecystectomy, Laparoscopic Gastrectomy | Surgical Instruments | Real, Also non-human data included | Public dataset(s), Private dataset(s) | m2cai-tl | Bounding box | VoTT, LabelMe | Unclear | n/a | CNN | not reported | Supervised |
| Penza, 2018 [119] | Radiology, Nuclear Medicine & Medical Imaging; Engineering, Biomedical; Computer Science, Interdisciplinary Applications; Computer Science, Artificial Intelligence | Computer Science | Visceral / General Surgery | Unclear | Organs/Tissue | Real, Also non-human data included | Private dataset(s) | n/a | Semantic, Classification | unclear | Unclear | n/a | Others/Unclear (Kanade-Lucas-Tomasi (KLT) Tracker and SURF (Speeded-Up Robust Features)) | Optical Flow | Others/unclear |
| Pradeep, 2022 [120] | Engineering Conference Paper | Computer Science | Visceral / General Surgery | Laparoscopic cholecystectomy | Surgical Instruments | Real | Public dataset(s) | m2cai-tl | Bounding box | unclear | Unclear | n/a | CNN | "Surgical Phase Aware Encoder-Decoder Architecture for Surgical Tool Presence Detection and Tool Localization" | Supervised |
| Prokopetc, 2015 [121] | Engineering Conference Paper | Computer Science | Gynecological Surgery | Laparoscopic Gynecological Surgery | Organs/Tissue | Real | Unclear | unclear | Bounding box | unclear | Unclear | n/a | Others/Unclear (DPM Model) | not reported | Supervised |
| Protserov, 2024 [122] | Health Care Sciences & Services; Medical Informatics | Interdisciplinary | Visceral / General Surgery | Laparoscopic cholecystectomy | Organs/Tissue, Technical guidance | Real | Public dataset(s), Private dataset(s) | open source videos | Semantic | unclear | Unclear | n/a | CNN, TM | not reported | Supervised |
| Rahbar, 2020 [123] | Surgery | Computer Science | Urological Surgery, Others | Robotic Lobectomy, Robotic Thoracoscopy, Robotic Prostectomy | Bleeding | Real | Public dataset(s) | US NLM | Dot | Adobe After Effects 2020 | No | n/a | Others/Unclear | "For real-time detection of arterial-type bleeding, the algorithm tracks the change in the entropy map distribution. That is, it tracks the local entropy maps from the prior step for each frame over a period of time. The local entropy map localizes the regions with a high degree of homogeneity. For the sake of quantification, we binarize the entropy map and compute the number of non-zeropixels as an indicator of uniformity of the different regions of the video's content with respect to time." | Supervised |
| Rahbar, 2024 [124] | Health Care Sciences & Services | Computer Science | Visceral / General Surgery | Laparoscopic cholecystectomy, Robotic Lobectomy, Robotic Thoracoscopy, Robotic Prostectomy | Surgical Instruments | Real, Also non-human data included | Public dataset(s) | US NLM, ROBUST-MIS, Endoscapes | Semantic | unclear | Unclear | n/a | CNN | not reported | Supervised |
| Raja, 2024 [125] | Engineering Conference Paper | Computer Science | Visceral / General Surgery | Laparoscopic cholecystectomy | Surgical Instruments | Real | Public dataset(s) | m2cai-tl | Bounding box | unclear | Unclear | n/a | CNN | not reported | Supervised |
| Ryu, 2024 [126] | Surgery | Interdisciplinary | Visceral / General Surgery | Laparoscopic Hemicolectomy | Organs/Tissue | Real | Private dataset(s) | n/a | Semantic | CVAT | Unclear | n/a | CNN | not reported | Supervised |
| Sahu, 2017 [127] | Surgery; Engineering, Biomedical; Radiology, Nuclear Medicine & Medical Imaging | Computer Science | Visceral / General Surgery | Laparoscopic cholecystectomy | Surgical Instruments | Real | Public dataset(s) | m2cai-t | Classification | unclear | No | n/a | CNN | "Temporal smoothing: It assumes that each tool transition within the endoscopic videos is | Supervised |

| | | | | | | | | | | | | | | | |
|---|---|---|---|---|---|---|---|---|---|---|---|---|---|---|---|
| | | | | | | | | | | | | | | smooth and takes previous frame detections into account in a weighted scheme. A window of five frames (including current and four previous frames) with normalized linear weights determines the current output detection." | |
| Samuel, 2021 [128] | Robotics | Computer Science | Visceral / General Surgery | Laparoscopic cholecystectomy | Surgical Instruments | Real, Also non-human data included | Public dataset(s) | Cholec80 | Classification | unclear | Unclear | n/a | CNN | not reported | Unsupervised |
| Sanchez-Matilla, 2022 [129] | Surgery; Engineering, Biomedical; Radiology, Nuclear Medicine & Medical Imaging | Computer Science | Visceral / General Surgery | Laparoscopic cholecystectomy | Organs/Tissue, Surgical Instruments | Real | Public dataset(s) | CholecSeg8k | Semantic | PixelAnnotaion Tool | Unclear | n/a | CNN | Multi-Stage TCN | Supervised |
| Satyanaik, 2024 [130] | Surgery; Engineering, Biomedical; Radiology, Nuclear Medicine & Medical Imaging | Computer Science | Visceral / General Surgery | Laparoscopic cholecystectomy | Organs/Tissue, Technical guidance | Real | Public dataset(s) | Endoscapes2023, Endoscapes-WC70 | Semantic | unclear | Unclear | n/a | CNN | not reported | Supervised, Others/unclear, Semi-supervised |
| Satyanaik, 2024 [131] | Engineering Conference Paper | Interdisciplinary | Visceral / General Surgery | Laparoscopic cholecystectomy | Organs/Tissue, Technical guidance | Real | Public dataset(s) | Endoscapes2023, Endoscapes-WC70 | Semantic, Classification | unclear | Unclear | n/a | CNN | not reported | Semi-supervised |
| Seenivasan, 2023 [132] | Preprint | Computer Science | Visceral / General Surgery | Laparoscopic cholecystectomy, Robotic Prostatectomy | Surgical Instruments | Real | Public dataset(s) | Cholec80, PSI-AVA | Classification | unclear | Unclear | n/a | CNN, TM | not reported | Supervised |
| Sengun, 2023 [133] | Surgery | Interdisciplinary | Visceral / General Surgery | Laparoscopic Adrenalectomy | Organs/Tissue | Real | Private dataset(s) | n/a | Semantic | CVAT | Yes (unclear criteria) | "of 66 videos, 40 met the inclusion criteria" | CNN | not reported | Supervised |
| Sengun, 2024 [134] | Surgery | Interdisciplinary | Visceral / General Surgery | Laparoscopic Adrenalectomy | Organs/Tissue | Real | Private dataset(s) | n/a | Semantic | LabelBox | No | n/a | CNN, TM | not reported | Supervised |
| Shen, 2023 [135] | Engineering, Biomedical; Radiology, Nuclear Medicine & Medical Imaging; Engineering, Electrical & Electronic; Imaging Science & Photographic Technology; Computer Science, Interdisciplinary Applications | Computer Science | Visceral / General Surgery | Unclear | Surgical Instruments | Real, Also non-human data included | Private dataset(s) | n/a | Semantic | unclear | Unclear | n/a | CNN, TM | not reported | Supervised |
| Sheng, 2024 [136] | Preprint | Interdisciplinary | Visceral / General Surgery | Laparoscopic cholecystectomy | Surgical Instruments, Organs/Tissue | Real, Also non-human data included, Synthetic | Public dataset(s) | CholecSeg8k | Semantic, No annotation | PixelAnnotaion Tool | Unclear | n/a | TM | not reported | Unsupervised |
| Shi, 2020 [137] | Telecommunications; Computer Science, Information Systems; Engineering, Electrical & Electronic | Interdisciplinary | Visceral / General Surgery | Laparoscopic cholecystectomy | Surgical Instruments | Real | Public dataset(s) | Cholec80 | Bounding box, Classification | unclear | Unclear | n/a | CNN | not reported | Supervised |
| Shimgekar, 2021 [138] | Engineering Conference Paper | Computer Science | Visceral / General Surgery | Unclear | Surgical Instruments | Real | Private dataset(s) | n/a | Semantic | LabelMe | Unclear | n/a | CNN | not reported | Supervised |
| Silva, 2022 [139] | Engineering Conference Paper | Computer Science | Visceral / General Surgery | Laparoscopic cholecystectomy | Organs/Tissue, Surgical Instruments | Real | Public dataset(s) | CholecSeg8k | Semantic | PixelAnnotaion Tool | Unclear | n/a | CNN, TM | not reported | Supervised |
| Smithmaitrie, 2024 [140] | Multidisciplinary Sciences | Interdisciplinary | Visceral / General Surgery | Unclear | Organs/Tissue | Real | Private dataset(s) | n/a | Bounding box | LabelImg | Yes (clear criteria) | "no cases involving cancerous lesions were included" | CNN | not reported | Supervised |
| Sonsilphong, 2022 [141] | Engineering Conference Paper | Interdisciplinary | Gynecological Surgery | Laparoscopic Hysterectomy | Organs/Tissue, Surgical Instruments | Real | Private dataset(s) | n/a | Bounding box | unclear | Unclear | n/a | CNN | not reported | Supervised |
| Streckert, 2023 [142] | Engineering Conference Paper | Computer Science | Visceral / General Surgery | Robotic Colorectal Surgery | Surgical Instruments | Real, Synthetic, Also non-human data included | Public dataset(s) | EndoVis15 | Semantic | unclear | Unclear | n/a | CNN | not reported | Supervised |
| Strong, 2024 [143] | Gastroenterology & Hepatology | Interdisciplinary | Visceral / General Surgery | Robotic Gastrectomy | Surgical Instruments | Real | Private dataset(s) | n/a | Semantic | unclear | Unclear | n/a | CNN | not reported | Supervised |
| Sun, 2022 [144] | Computer Science | Computer Science | Visceral / General Surgery | Unclear | Surgical Instruments | Real | Private dataset(s) | n/a | Classification | unclear | Unclear | n/a | CNN | not reported | Supervised |
| Takeuchi, 2023 [145] | Surgery | Interdisciplinary | Visceral / General Surgery | Laparoscopic Hernia Repair | Organs/Tissue, Technical guidance | Real | Private dataset(s) | n/a | Bounding box | CVAT | Yes (unclear criteria) | "set consisting of both positive examples (images where anatomical structured were present) and negative examples (images where structures were not present)." | CNN | not reported | Supervised |
| Tao, 2023 [146] | Engineering, Biomedical; Radiology, Nuclear Medicine & Medical Imaging; Engineering, Electrical & Electronic; Imaging Science & Photographic Technology; Computer Science, Interdisciplinary Applications | Computer Science | Visceral / General Surgery | Laparoscopic cholecystectomy | Surgical Instruments | Real | Public dataset(s) | Cholec80, m2cai-t | Classification | unclear | No | n/a | TM, CNN | "we propose to use a banded causal mask to limit the dependency range between frames while ensuring that future information is masked, thus supporting on-line surgical workflow analysis." | Supervised |
| Tokuyasu, 2021 [147] | Surgery | Interdisciplinary | Visceral / General Surgery | Laparoscopic cholecystectomy | Organs/Tissue | Real | Private dataset(s) | n/a | Semantic | unclear | Yes (clear criteria) | "As the degree of difficulty in LC increases in tandem with the extent of fibrosis and/or scarring inside the abdominal cavity, the technical platform of our system was established using videos with minimal fibrosis and/or scarring; videos with bleeding or less-visible landmarks were also excluded." | CNN | not reported | Supervised |
| Urrea, 2024 [148] | Pharmacology & Pharmacy; Biochemistry & Molecular Biology; Medicine, Research & Experimental | Computer Science | Visceral / General Surgery | Laparoscopic cholecystectomy | Organs/Tissue, Surgical Instruments, | Real | Public dataset(s) | CholecSeg8K | Semantic | PixelAnnotaion Tool | Unclear | n/a | CNN | Spatial–Temporal Convolutional Network (SP-TCN) | Supervised |

| | | | | | | | | | | | | | | | |
|---|---|---|---|---|---|---|---|---|---|---|---|---|---|---|---|
| | | | | | Technical guidance | | | | | | | | | | |
| Valderrama, 2022 [149] | Preprint | Interdisciplinary | Urological Surgery | Robotic Prostatectomy | Surgical Instruments | Real | Private dataset(s) | PSI-AVA | Bounding box, Classification | unclear | Yes (unclear criteria) | "removing video segments with non-surgical content and potential patient identifiers" | TM, CNN | "We build upon the Multiscale Vision Transformer (MViT) model developed for video and image recognition tasks [9]. The connection between multiscale feature hierarchies with a transformer model, provides rich features to model the complex temporal cues of the surgical scene around each keyframe" | Supervised |
| Vardazaryan, 2018 [150] | Preprint | Computer Science | Visceral / General Surgery | Laparoscopic cholecystectomy | Surgical Instruments | Real | Public dataset(s) | Cholec80 | Classification, Bounding box | unclear | Unclear | n/a | CNN | not reported | Weakly-supervised |
| Wagner, 2023 [151] | Radiology, Nuclear Medicine & Medical Imaging; Engineering, Biomedical; Computer Science, Interdisciplinary Applications; Computer Science, Artificial Intelligence | Interdisciplinary | Visceral / General Surgery | Laparoscopic cholecystectomy | Surgical Instruments | Real | Public dataset(s), Private dataset(s) | ROBUST-MIS | Classification | Anvil | Unclear | n/a | CNN | not reported | Supervised |
| Wang, 2023 [152] | Engineering Conference Paper | Computer Science | Visceral / General Surgery | Laparoscopic cholecystectomy | Surgical Instruments | Real | Public dataset(s) | Cholec80 | Classification | unclear | Unclear | n/a | CNN | not reported | Supervised |
| Wang, 2017 [153] | Engineering Conference Paper | Computer Science | Visceral / General Surgery | Laparoscopic cholecystectomy | Surgical Instruments | Real | Public dataset(s) | m2cai-t | Classification | unclear | Unclear | n/a | CNN | not reported | Supervised |
| Wang, 2024 [154] | Preprint | Interdisciplinary | Visceral / General Surgery | Laparoscopic cholecystectomy | Surgical Instruments | Real | Public dataset(s) | Cholec80, m2cai-tl | Classification, Bounding box | unclear | Unclear | n/a | CNN | "HMM-stabilized deep learning model; LSTM-enhanced version of EndoNet and FCN compared to 4 frame-independent methods, ToolNet, EndoNet, SwinNet, and FR-CNN" | Semi-supervised |
| Wang, 2023 [155] | Surgery | Interdisciplinary | Urological Surgery | Robotic Partial Nephrectomy | Surgical Instruments | Real | Private dataset(s) | n/a | Semantic | unclear | Unclear | n/a | CNN | "mtCNN processes the sequence of instrument features across all the video frames" | Supervised |
| Wang, 2025 [156] | Radiology, Nuclear Medicine & Medical Imaging; Engineering, Biomedical; Computer Science, Interdisciplinary Applications; Computer Science, Artificial Intelligence | Computer Science | Urological Surgery | Robotic Prostatectomy | Surgical Instruments | Real | Public dataset(s) | GraSP | Bounding box, Semantic | unclear | Unclear | n/a | CNN, TM | Stereo-temporal set classifier (STSCls) | Supervised |
| Wang, 2021 [157] | Engineering Conference Paper | Computer Science | Visceral / General Surgery | Laparoscopic cholecystectomy | Surgical Instruments | Real | Public dataset(s) | m2cai-tl | Bounding box | unclear | Unclear | n/a | CNN | not reported | Supervised |
| Wang, 2020 [158] | Telecommunications; Computer Science, Information Systems; Engineering, Electrical & Electronic | Computer Science | Visceral / General Surgery | Laparoscopic cholecystectomy | Surgical Instruments | Real | Private dataset(s), Public dataset(s) | m2cai-tl | Bounding box | unclear | Unclear | n/a | CNN | not reported | Supervised |
| Wang, 2024 [159] | Preprint | Interdisciplinary | Visceral / General Surgery | Laparoscopic cholecystectomy, Robotic Liver Surgery | Surgical Instruments | Real | Public dataset(s) | Cholec80 | Semantic, Classification | unclear | Unclear | n/a | TM | "We take temporal properties into account by inducing two temporal constraints for traditional paradigm, temporal equivariance and class-aware temporal semantic continuity constraints, together with temporal-enhanced pseudo masks generation; CAM" | Weakly-supervised |
| Wang, 2023 [160] | Robotics; Engineering, Biomedical | Computer Science | Visceral / General Surgery | Laparoscopic cholecystectomy | Surgical Instruments | Real | Public dataset(s) | m2cai-tl | Bounding box | unclear | Unclear | n/a | CNN | not reported | Supervised |
| Ward, 2022 [161] | Surgery | Interdisciplinary | Visceral / General Surgery | Laparoscopic cholecystectomy | Organs/Tissue, Others | Real | Private dataset(s) | n/a | Classification | unclear | Unclear | n/a | CNN | not reported | Supervised |
| Wei, 2024 [162] | Surgery; Engineering, Biomedical; Radiology, Nuclear Medicine & Medical Imaging | Computer Science | Visceral / General Surgery | Laparoscopic cholecystectomy, Laparoscopic Colectomy, Laparoscopic Rectal Resection, Laparoscopic Hernia Repair, Laparoscopic Sigmoid Resection | Surgical Instruments | Real | Public dataset(s) | ROBUST-MIS | Semantic | understand.ai | Unclear | n/a | TM, CNN | not reported | Supervised |
| Wei, 2023 [163] | Preprint | Interdisciplinary | Visceral / General Surgery | Laparoscopic Colectomy, Laparoscopic Rectal Resection, Laparoscopic Hernia Repair, Laparoscopic Sigmoid Resection | Surgical Instruments | Real, Also non-human data included | Public dataset(s) | ROBUST-MIS | Semantic | Understand.ai | Unclear | n/a | CNN | not reported | Semi-supervised |

| | | | | | | | | | | | | | | | |
|---|---|---|---|---|---|---|---|---|---|---|---|---|---|---|---|
| Wu, 2024 [164] | Computer Science, Interdisciplinary Applications; Biology; Mathematical & Computational Biology; Engineering, Biomedical | Interdisciplinary | Gynecological Surgery | Laparoscopic Hysterectomy | Surgical Instruments | Real | Public dataset(s) | AutoLaparo | Semantic | LabelMe | Unclear | n/a | CNN | not reported | Supervised |
| Xi, 2022 [165] | Engineering, Electrical & Electronic | Computer Science | Visceral / General Surgery | Laparoscopic cholecystectomy | Surgical Instruments | Real | Public dataset(s) | CholecT50 | Classification | unclear | Unclear | n/a | Others/Unclear (Graph Convolutional Network (GCN)) | not reported | Supervised |
| Xu, 2024 [166] | Preprint | Computer Science | Urological Surgery | Robotic Prostatectomy | Surgical Instruments | Real, Also non-human data included | Public dataset(s) | SAR-RARP | Semantic | unclear | Unclear | n/a | CNN, TM | not reported | Supervised, Others/unclear |
| Xue, 2022 [167] | Engineering, Biomedical | Computer Science | Visceral / General Surgery | Laparoscopic cholecystectomy | Surgical Instruments | Real, Also non-human data included | Public dataset(s) | m2cai-tl | Bounding box | unclear | Unclear | n/a | CNN | not reported | Supervised |
| Yamazaki, 2020 [168] | Surgery | Interdisciplinary | Visceral / General Surgery | Laparoscopic Gastrectomy | Surgical Instruments | Real | Private dataset(s) | n/a | Bounding box | unclear | Unclear | n/a | CNN | not reported | Supervised |
| Yamlahi, 2023 [169] | Preprint | Computer Science | Visceral / General Surgery | Laparoscopic cholecystectomy | Surgical Instruments | Real | Public dataset(s) | CholecT45 | Classification | unclear | Unclear | n/a | TM | not reported | Supervised |
| Yang, 2024 [170] | Engineering Conference Paper | Computer Science | Visceral / General Surgery | Laparoscopic cholecystectomy | Organs/Tissue | Real | Private dataset(s) | n/a | Bounding box | Labelimg | Yes (unclear criteria) | "Out of the 101 surgical videos, 80 were selected based on their good visual effects and compliance with the selection criteria." | CNN | not reported | Supervised |
| Yang, 2023 [171] | Preprint | Interdisciplinary | Visceral / General Surgery | Laparoscopic cholecystectomy | Organs/Tissue, Surgical Instruments | Real | Public dataset(s) | CholecSeg8K | Semantic | PixelAnnotaion Tool | Unclear | n/a | CNN | not reported | Supervised |
| Yang, 2021 [172] | Engineering Conference Paper | Interdisciplinary | Visceral / General Surgery | Laparoscopic cholecystectomy | Surgical Instruments | Real | Public dataset(s) | Cholec80 | Unclear | unclear | Yes (unclear criteria) | "13 videos from the dataset are eliminated due to bad segmentation performance." | CNN | not reported | Others/unclear |
| Yin, 2024 [173] | Preprint | Interdisciplinary | Visceral / General Surgery | Laparoscopic cholecystectomy | Organs/Tissue, Surgical Instruments, Technical guidance | Real | Public dataset(s) | CholecT45, Cholec80-CVS | Classification | unclear | Unclear | n/a | TM | "Prediction of following steps in the next frames" | Supervised |
| Yuan, 2024 [174] | Surgery; Engineering, Biomedical; Radiology, Nuclear Medicine & Medical Imaging | Computer Science | Visceral / General Surgery | Laparoscopic cholecystectomy | Surgical Instruments, Organs/Tissue | Real | Public dataset(s) | CholecT45, m2cai-tl, CholecSeg8k | Bounding box, Semantic, Classification | PixelAnnotaion Tool | Yes (unclear criteria) | "To reduce class-imbalance" | CNN, TM | not reported | Supervised |
| Zhang, 2024 [175] | Robotics | Interdisciplinary | Visceral / General Surgery | Laparoscopic cholecystectomy | Organs/Tissue | Real | Private dataset(s) | n/a | Semantic, Bounding box | unclear | Unclear | n/a | Others/Unclear (TAI-GNN) | "Temporal Graph: During the CTS process, video frame data is transformed into a sequence of spatial graphs. These graphs are then cascaded by connecting the same-type target nodes to each graph, forming a spatial-temporal graph" | Supervised |
| Zhang, 2023 [176] | Engineering Conference Paper | Computer Science | Visceral / General Surgery | Laparoscopic cholecystectomy | Surgical Instruments | Real | Public dataset(s) | Cholec80 | Classification, Bounding box | LabelIMG | Unclear | n/a | CNN | DeepSORT (Simple Online and Realtime Tracker mit Deep Learning) | Supervised |
| Zhang, 2023 [177] | Engineering, Biomedical | Computer Science | Visceral / General Surgery | Laparoscopic cholecystectomy | Surgical Instruments | Real | Public dataset(s) | Cholec80 | Classification | unclear | Unclear | n/a | TM, CNN | "MS-TCN, a recent state-of-the-art architecture in action segmentation, improved on previous approaches by adopting a fully convolutional architecture for processing the temporal dimension of the video" | Supervised |
| Zhang, 2020 [178] | Telecommunications; Computer Science, Information Systems; Engineering, Electrical & Electronic | Interdisciplinary | Visceral / General Surgery | Laparoscopic cholecystectomy | Surgical Instruments | Real | Public dataset(s), Private dataset(s) | m2cai-tl | Bounding box | unclear | Unclear | n/a | CNN | not reported | Supervised |
| Zhang, 2023 [179] | Computer Science, Interdisciplinary Applications; Computer Science, Information Systems; Mathematical & Computational Biology; Medical Informatics | Interdisciplinary | Visceral / General Surgery | Laparoscopic cholecystectomy | Surgical Instruments, Technical Guidance | Real | Private dataset(s) | n/a | Classification | LabelImg | Unclear | n/a | CNN | Spatial-temporal patterns | Supervised |
| Zhang, 2020 [180] | Surgery; Engineering, Biomedical; Radiology, Nuclear Medicine & Medical Imaging | Computer Science | Visceral / General Surgery | Laparoscopic cholecystectomy | Surgical Instruments | Real | Public dataset(s) | m2cai-t | Classification, Dot | unclear | Unclear | n/a | CNN | not reported | Supervised |
| Zhao, 2021 [181] | Radiology, Nuclear Medicine & Medical Imaging; Engineering, Biomedical; Computer Science, Interdisciplinary Applications; Computer Science, Artificial Intelligence | Interdisciplinary | Urological Surgery, Others | Robotic Prostatectomy | Surgical Instruments | Real, Also non-human data included | Private dataset(s) | n/a | Semantic | Viame | Unclear | n/a | CNN | Anchor matching mechanism | Supervised, Unsupervised |
| Zhao, 2023 [182] | Medical Informatics | Interdisciplinary | Visceral / General Surgery, Others | Laparoscopic cholecystectomy | Surgical Instruments | Real | Public dataset(s) | Cholec80 | Bounding box | unclear | Unclear | n/a | CNN | "cross-layer aggregated attention detection network (CLAD-Net)" | Supervised |

| | | | | | | | | | | | | | | | |
|---|---|---|---|---|---|---|---|---|---|---|---|---|---|---|---|
| Zhao, 2021 [183] | Preprint | Interdisciplinary | Urological Surgery | Robotic Prostatectomy | Surgical Instruments | Real, Also non-human data included | Public dataset(s) | HKPWH | Semantic | unclear | Unclear | n/a | CNN | "In order to avoid the negative effect of the noisy supervision and increase the model robustness to tackle large temporal variation, we propose to selectively update the model by leveraging the available label of first frame to assess the quality of each pseudo mask." | Semi-supervised |
| Zhao, 2019 [184] | Surgery | Computer Science | Others, Visceral / General Surgery | Unclear | Surgical Instruments | Real, Also non-human data included | Unclear | unclear | Bounding box | unclear | Unclear | n/a | CNN, TM | "spatial transformer network (STN) for the location-determination process and a spatio-temporal context learning algorithm (STC) for the process of tracking frame by frame" | Supervised |
| Zheng, 2023 [185] | Surgery | Interdisciplinary | Urological Surgery, Visceral / General Surgery | Laparoscopic cholecystectomy, Laparoscopic Prostatectomy, Laparoscopic Partial Adrenal Resection, Laparoscopic Bladder Resection | Surgical Instruments | Real | Private dataset(s), Public dataset(s) | Cholec80 | Semantic | LabelMe | Yes (clear criteria) | "Exclusion criteria included any of the following, (i) A visual pedestal, (ii) smoke, (iii) blurred image, or (iv) specular reflection." | CNN | not reported | Supervised |
| Zhou, 2024 [186] | Engineering Conference Paper | Interdisciplinary | Visceral / General Surgery | Laparoscopic cholecystectomy | Organs/Tissue, Technical guidance | Real | Public dataset(s) | Cholec80 | Semantic | unclear | Unclear | n/a | TM | not reported | Supervised |
| Zhou, 2022 [187] | Engineering Conference Paper | Computer Science | Visceral / General Surgery | Laparoscopic cholecystectomy | Surgical Instruments | Real | Public dataset(s) | m2cai-tl | Bounding box | unclear | Unclear | n/a | CNN | not reported | Supervised |
| Zygomalas, 2024 [188] | Surgery | Interdisciplinary | Visceral / General Surgery | Laparoscopic Hernia Repair | Organs/Tissue, Surgical Instruments | Real | Private dataset(s), unclear | unclear | Bounding box | SuperAnnotate | Unclear | n/a | CNN | not reported | Supervised |

Abbreviations: *n/a: not applicable, CNN: Convolutional Neural Network, TM: Transformer Model*

**Footnotes:**

a. Searched on: https://clarivate.com/academia-government/scientific-and-academic-research/research-funding-analytics/journal-citation-reports/. If the journal was not available, for example, because it is too new, the criteria from the official journal website were used. If it is a preprint journal, it was marked as "Preprint." If it is a conference paper, it was labeled as "Engineering Conference Paper," since they are exclusively in the field of engineering.
b. If the name was provided and if it is a dataset consisting of human data.

**Supplementary Table 4**: Studies and study characteristics included in the systematic review and meta-analysis (2)

The following table presents an overview of all included studies, detailing key characteristics across four main categories: **Validation Strategies, Dataset Size, Model Performance Assessment, Uncertainty Estimation**.

| Publication | Validation Strategies | | | | | Dataset Size | | | | | | | Model Performance Assessment | | | | | | Uncertainty Estimation | | | |
|---|---|---|---|---|---|---|---|---|---|---|---|---|---|---|---|---|---|---|---|---|---|---|
| First author, year [reference] | Validation method | If cross-validation X Fold | Validation distribution[a] | Dataset Validation[b] | Independence of test samples when splitting the data | Number of Patients[c] | Number of institutions[c] | Dataset Size[d] | No. videos training + validation | No. frames training + validation | No. videos testing | No. frames testing | Object detection metrics | Calculation of performance metrics[e] | Temporal metrics use | Visual Reporting | Additional Imaging Modality | Real-time modeling (inference time) | Metrics for reporting model uncertainty and variability | Mode of reporting model uncertainty and prediction variability | Calculation of model uncertainty and prediction variability[e] | Other statistical tests[f] |
| Acharya, 2022 [1] | Cross-Validation, Hold-out | 10 | unclear | Internal Retrospective | "Yes, on frame level" | 102 | 1 | 25682 frames | unclear | unclear | unclear | unclear | Accuracy, Recall, Precision | Unclear | no | no | No | unclear | SD | Exact numbers | Unclear | none |
| Al Hajj, 2018 [2] | Hold-out | n/a | 45/5/50 | Internal Retrospective | "Yes, on patient and frame level" | 80 | 1 | 80 videos | 40 | unclear | 40 | unclear | ROC Curve, mAP | Unclear | no | others | No | 2.87 ms | None | None | none | "Frequentist parametric (e.g., t-test)" |
| Alapatt, 2021 [3] | Hold-out | n/a | unclear | Internal Retrospective | "Yes, on frame level" | 201 | 1 | 1933 frames | unclear | unclear | unclear | unclear | F1-Score, mIoU, Accuracy | Unclear | no | Qualitative results | No | n/a | None | None | none | none |
| Ali, 2022 [4] | Hold-out | n/a | unclear | Internal Retrospective | "Yes, on frame level" | 15 | 1 | 2812 frames | unclear | 2531 | unclear | 281 | mAP | Unclear | no | Qualitative results | No | n/a | None | None | none | "Frequentist parametric (e.g., t-test)" |
| Alkhamaiseh, 2023 [5] | Cross-Validation | 5 | unclear | Internal Retrospective | "Yes, on frame level" | 200 | unclear | 1550 frames | unclear | 1395 | unclear | 155 | DSC, Precision, Recall, IoU, Others | Over frame-level averages of n cross-validation folds | no | Qualitative results | No | 24 ms | None | None | none | none |
| Alshirbaji, 2021 [6] | Cross-Validation | 6 | 50/50 | Internal Retrospective | "Yes, on patient and frame level" | 80 | 1 | 80 videos | 40 | unclear | 40 | unclear | mAP | Over sample-level averages of n cross-validation folds | no | Class activation map | No | n/a | SD | Graph | Over sample-level averages of n cross-validation folds | none |
| Alshirbaji, 2021 [7] | Hold-out | n/a | unclear | Internal Retrospective | Unclear | 100 | 3 | 92305 frames | 80, 20 | unclear | 20/80 | unclear | AP | Unclear | no | no | No | n/a | SD | Graph | Unclear | none |
| Alshirbaji, 2018 [8] | Hold-out | n/a | 50/50 | Internal Retrospective | "Yes, on patient and frame level" | 80 | 1 | 80 videos | 40 | unclear | 40 | unclear | Others, Accuracy, Recall, Precision | Unclear | no | others | No | n/a | None | None | none | none |
| Ángeles-Cerón, 2021 [9] | Hold-out | n/a | unclear | Internal Retrospective | "Yes, on frame level", "Yes, on patient and frame level" | unclear | 1 | 4987 frames | unclear | 4239 | unclear | 748 | Others | Unclear | no | Qualitative results | No | >45 FPS | IQR | Graph | Unclear | none |
| Ángeles-Cerón, 2022 [10] | Hold-out | n/a | unclear | Internal Retrospective | "Yes, on patient and frame level", "Yes, on frame level" | 30 | 1 | 9044 frames | unclear | 4987 | unclear | 4057 | Others | Over n frames in test set | no | Qualitative results, others, Feature Map | No | 24 FPS | IQR | Graph | Over n frames in test set | none |
| Aoyama, 2024 [11] | Hold-out | n/a | 45/5 | Prospective Interventional | "Yes, on patient and frame level" | 60 | 1 | 2771 frames | unclear | 2493 | unclear | 278 | DSC | Unclear | no | Qualitative results | No | 210 ms | None | None | none | "Frequentist non-parametric (e.g., Mann-Whitney)" |
| Arabian, 2022 [12] | Hold-out | n/a | 50/x/50 | Internal Retrospective | "Yes, on patient and frame level" | 80 | 1 | 80 Videos | 40 | unclear | 40 | unclear | mAP | Over n frames in test set | no | Class activation map | No | n/a | SD | Graph | Unclear | none |
| Arabian, 2023 [13] | Hold-out | n/a | 50/50 | Internal Retrospective, External Retrospective | "Yes, on patient and frame level" | 80 | 1 | 80 videos | 40 | unclear | 40 | unclear | mAP | Over n frames in test set | no | no | No | n/a | SD | Exact numbers, Graph | Unclear | none |
| Aspart, 2022 [14] | Cross-Validation | 5 | 90/10 | Internal Retrospective | "Yes, on patient and frame level" | 300 | 1 | 122470 frames | 271 | 111154 | 29 | 11316 | ROC Curve, Sensitivity, Specificity | Unclear | no | Class activation map | No | 16 FPS | SD | Exact numbers | Unclear | none |
| Attia, 2017 [15] | Hold-out | n/a | unclear | Internal Retrospective | "Yes, on frame level" | 4 | 1 | unclear | unclear | unclear | unclear | unclear | IoU, Sensitivity, Specificity, Accuracy | Unclear | no | Qualitative results | No | n/a | None | None | none | none |
| Ayobi, 2024 [16] | Cross-Validation | 2 | unclear | Prospective Observational | "Yes, on patient and frame level" | 13 | 1 | 13 videos | 8 | unclear | 5 | unclear | mAP, mIoU, Others | Unclear | unclear | Qualitative results | No | n/a | SD | Exact numbers | Unclear | none |
| Bai, 2023 [17] | Hold-out | n/a | unclear | Internal Retrospective | "Yes, on frame level" | 15 | 1 | 2811 frames | unclear | unclear | unclear | unclear | Accuracy, mIoU | Unclear | no | Qualitative results | No | n/a | None | None | none | none |
| Bakker, 2024 [18] | Hold-out | n/a | 90/10 | Internal Retrospective | "Yes, on patient and frame level" | 111 | 2 | 1694 frames | unclear | 1522 | unclear | 172 | DSC, Others | Unclear | no | Qualitative results | No | n/a | SD | Exact numbers, Graph | Unclear | Other |
| Bamba, 2021 [19] | Hold-out | n/a | unclear | Internal Retrospective | "Yes, on patient and frame level", "Yes, on frame level" | 10 | 1 | 1270 frames | unclear | 1070 | unclear | 200 | Accuracy, Precision, Recall, mAP, IoU | Unclear | no | Qualitative results | No | unclear | CI | Exact numbers | Unclear | none |

| | | | | | | | | | | | | | | | | | | | | | | |
|---|---|---|---|---|---|---|---|---|---|---|---|---|---|---|---|---|---|---|---|---|---|---|
| Bamba, 2021 [20] | Hold-out | n/a | unclear | Prospective Observational | "Yes, on patient and frame level" | 17 | 1 | 1673 frames | 11 | 1173 | 6 | 500 | Accuracy, mAP, Precision, Recall, IoU, Covidence Score | Unclear | no | Qualitative results | No | n/a | CI | Exact numbers | Unclear | none |
| Ban, 2023 [21] | Cross-Validation, Hold-out | 3, 5 | 90/10 | Internal Retrospective | "Yes, on patient and frame level" | 345 | unclear | 345 videos | unclear | unclear | unclear | unclear | AP, Accuracy, Others | Unclear | no | others | No | n/a | SD | Exact numbers | Unclear | none |
| Batić, 2024 [22] | unclear | unclear | unclear | External Retrospective | "Yes, on frame level" | unclear | unclear | 743724 frames | unclear | unclear | unclear | unclear | mIoU, mAP | Unclear | no | Qualitative results | No | n/a | SD | Exact numbers | Unclear | none |
| Batić, 2023 [23] | Hold-out | n/a | unclear | External Retrospective | "Yes, on frame level" | unclear | unclear | >700000 frames | unclear | unclear | unclear | unclear | mAP | Unclear | no | Qualitative results | No | unclear | SD | Exact numbers | Unclear | none |
| Boonkong, 2022 [24] | unclear | n/a | unclear | Internal Retrospective | "Yes, on frame level" | unclear | unclear | 870 frames | unclear | unclear | unclear | unclear | Precision, Recall, F1-Score | Unclear | no | Qualitative results | No | n/a | None | None | none | none |
| Brandenburg, 2023 [25] | Hold-out | n/a | ca 85/15 | Prospective Observational, Prospective Interventional | "Yes, on patient and frame level" | 26 | 2 | 14004 frames | 22 | 13400 | 4 | 604 | F1-Score, Precision, Recall | Over n frames in test set | no | others | No | 12 FPS | SD | Exact numbers, Graph | Over n frames in test set | "Frequentist parametric (e.g., t-test)" |
| Casella, 2021 [26] | Hold-out | n/a | 75/12.5/12.5 | Internal Retrospective | "Yes, on patient and frame level" | 8 | 1 | 1871 frames | 7 | 1631 | 1 | 240 | Precision, Recall, DSC | Over n frames in test set | no | Qualitative results | No | n/a | Standard Error | Exact numbers | Over n frames in test set | "Frequentist non-parametric (e.g., Mann-Whitney)" |
| Chen, 2024 [27] | Hold-out | n/a | unclear | Internal Retrospective, External Retrospective | "Yes, on frame level" | unclear | 2 | unclear (0ver 3 mio. frames) | unclear | unclear | unclear | unclear | mIoU, mDSC | Unclear | no | Qualitative results | No | n/a | None | None | none | "Frequentist non-parametric (e.g., Mann-Whitney)" |
| Chen, 2013 [28] | Hold-out | n/a | n/a | Prospective Observational | "Yes, on patient and frame level" | 1 | 1 | 907 frames | 1 | 10 | 1 | 897 | Others | Over n frames in test set | no | Qualitative results | No | 8 FPS | None | None | none | none |
| Chen, 2017 [29] | Hold-out | n/a | unclear | Internal Retrospective | "Yes, on patient and frame level" | unclear | unclear | 1000 frames | unclear | unclear | unclear | unclear | Accuracy | Over n frames in test set | no | Qualitative results | No | 25.41 FPS | None | None | none | none |
| Choi, 2017 [30] | Hold-out | n/a | 70/30 | Internal Retrospective | "Yes, on patient and frame level" | 10 | 1 | 10 videos | 7 | unclear | 3 | unclear | Recall, Precision, mAP | "Over n independent samples (i.e., one patient/video) in independent test set" | no | Qualitative results | No | 48.9 FPS | Other | Exact numbers | Other | none |
| Ciaparrone, 2020 [31] | Hold-out | n/a | unclear | Internal Retrospective | "Yes, on patient and frame level" | 14 | unclear | 4955 frames | unclear | 3485 | unclear | 1470 | AP | Over n frames in test set | no | Qualitative results | No | 400 ms | None | None | none | none |
| Colleoni, 2024 [32] | Hold-out | n/a | unclear | Internal Retrospective | "Yes, on frame level" | unclear | unclear | 254678 frames | 1772 | 230455 | 183 | 24223 | mIoU | Over n frames in test set | no | Qualitative results | No | n/a | SD | Exact numbers | Over n frames in test set | none |
| Colleoni, 2022 [33] | Hold-out | n/a | unclear | Internal Retrospective | Unclear | 23 | 1 | 7236 frames | unclear | 6248 | unclear | 988 | IoU | Unclear | no | Qualitative results | No | n/a | None | None | none | none |
| Daneshgar Rahbar, 2023 [34] | Hold-out | n/a | unclear | External Retrospective | "Yes, on frame level" | unclear | unclear | unclear | unclear | unclear | unclear | unclear | mDSC, mIoU, Accuracy | Over n frames in test set | no | Qualitative results | No | 34.2 ms | None | None | none | none |
| Davila, 2023 [35] | unclear | unclear | unclear | Internal Retrospective | Unclear | 80 | 1 | 80 videos | unclear | 86331 | unclear | unclear | AP, Accuracy | Unclear | no | no | No | n/a | Standard Error | Graph | Unclear | none |
| De Backer, 2023 [36] | Hold-out | n/a | 70/20/10 | Internal Retrospective | "Yes, on patient and frame level" | 67 | 1 | 15100 frames | 52 | 13592 | 5 | 1508 | IoU, DSC | Over n frames in test set | no | Qualitative results | No | 500 ms | None | None | none | none |
| den Boer, 2023 [37] | Cross-Validation | 5 | 80/20 | Internal Retrospective | "Yes, on patient and frame level" | 83 | 1 | 1050 frames | 66 | 850 | 17 | 200 | DSC, Others | Unclear | no | Qualitative results | No | 39 FPS | IQR | Exact numbers | Unclear | "Frequentist non-parametric (e.g., Mann-Whitney)" |
| Derathé, 2020 [38] | Cross-Validation | 10 | unclear | Internal Retrospective | "Yes, on patient and frame level" | 29 | 1 | 29 videos | unclear | unclear | unclear | unclear | Accuracy, Sensitivity, Specificity | Unclear | no | no | No | n/a | SD | Exact numbers, Graph | Unclear | none |
| Du, 2019 [39] | unclear | n/a | unclear | External Retrospective, Internal Retrospective | Unclear | unclear | unclear | unclear | unclear | unclear | unclear | unclear | Others | Unclear | no | Qualitative results | No | unclear | None | None | none | none |
| Endo, 2023 [40] | unclear | n/a | 80/20 | Internal Retrospective | "Yes, on patient and frame level" | 115 | 1 | 1754 frames | unclear | unclear | unclear | unclear | Others | Unclear | no | Qualitative results | No | unclear | None | None | none | none |
| Fernandez-Rodríguez, 2024 [41] | Hold-out | n/a | 58/22/20 | Internal Retrospective | "Yes, on patient and frame level" | 80 | 1 | 8080 frames | 13 | 6480 | 4 | 1600 | DSC, Recall, Precision | Over sample-level averages of n cross-validation folds | no | Qualitative results | No | n/a | SD | Exact numbers | Over sample-level averages of n cross-validation folds | none |
| Fuentes-Hurtado, 2019 [42] | Cross-Validation, Hold-out | 5 | unclear | Internal Retrospective | "Yes, on patient and frame level" | 29 | 2 | 50366 frames | unclear | unclear | unclear | unclear | IoU, Others | Over sample-level averages of n cross-validation folds | no | Qualitative results, others | No | n/a | None | None | none | none |
| Fujinaga, 2023 [43] | unclear | unclear | unclear | Prospective Interventional | "Yes, on patient and frame level" | 122 | 1 | 1826 frames | unclear | unclear | unclear | unclear | DSC | Unclear | no | Qualitative results | No | 30 FPS | SD | Exact numbers | Unclear | Other |
| Ghamsarian, 2024 [44] | Cross-Validation | 4 | 75/x/25 | Internal Retrospective | Unclear | 108 | 1 | 158 frames | unclear | 119 | unclear | 39 | IoU, DSC | Unclear | no | no | No | n/a | None | None | none | none |
| Gitau, 2024 [45] | Hold-out | n/a | unclear | Internal Retrospective | "Yes, on frame level" | unclear | 1 | 3299 frames | unclear | unclear | unclear | unclear | Accuracy | Over n frames in test set | no | no | No | n/a | None | None | none | none |
| Grammatikopoulou, 2023 [46] | Hold-out | n/a | 85/5/10 (PN) 75/25 (CholecSeg8k) | Internal Retrospective | "Yes, on patient and frame level" | 154 | 2 | 61080 frames | 126 | unclear | 18 | unclear | mIoU | Over n frames in test set | TC: 56.13 (PN), 87.26 (Cholec) | Qualitative results | No | n/a | None | None | none | none |

| | | | | | | | | | | | | | | | | | | | | | | |
|---|---|---|---|---|---|---|---|---|---|---|---|---|---|---|---|---|---|---|---|---|---|---|
| Guédon, 2021 [47] | Cross-Validation | 5 | 24/9/x (LC) 26/9/x (TLH) | Internal Retrospective | "Yes, on patient and frame level" | 68 | 1 | 68 videos | 50 | unclear | 18 | unclear | Accuracy, Precision, Recall | Unclear | no | no | No | n/a | None | None | none | none |
| Hasan, 2021 [48] | Hold-out | n/a | unclear | Internal Retrospective | Unclear | 4 | 1 | unclear | unclear | unclear | unclear | unclear | DSC | Over n frames in test set | no | Qualitative results | No | n/a | SD | Exact numbers | Over n frames in test set | none |
| Huang, 2022 [49] | Hold-out | n/a | unclear | External Retrospective | "Yes, on frame level" | unclear | unclear | 48702 frames | unclear | 34320 | unclear | 14382 | IoU, DSC, Others | Unclear | no | Qualitative results, Depth map | No | 5.8 ms / 172 FPS | SD | Exact numbers | Unclear | none |
| Jalal, 2022 [50] | Hold-out | n/a | 50/50 | Internal Retrospective | "Yes, on patient and frame level" | 80 | 1 | 15691 frames (bounding boxes) | 40 | unclear | 40 | unclear | mDSC, Precision, Others | Over n frames in test set | no | no | No | n/a | None | None | none | none |
| Jalal, 2023 [51] | Hold-out | n/a | 50/50 | Internal Retrospective | "Yes, on patient and frame level" | 80 | 1 | unclear | 40 | unclear | 40 | unclear | mAP, F1-Score | Over n frames in test set | no | Qualitative results, Localisation Map | No | 20 ms | None | None | none | none |
| Jamal, 2024 [52] | Hold-out | n/a | unclear | Internal Retrospective | "Yes, on patient and frame level", "Yes, on frame level" | 88 | unclear | 26130 frames | unclear | unclear | unclear | unclear | IoU | Unclear | no | no | Other | n/a | None | None | none | none |
| Jang, 2023 [53] | unclear | n/a | unclear | Internal Retrospective | Unclear | unclear | unclear | 157 frames | unclear | 137 | unclear | 20 | Others | Unclear | no | Qualitative results | No | n/a | None | None | none | none |
| Jaspers, 2024 [54] | Cross-Validation | 5 | 77/23 | External Retrospective | "Yes, on patient and frame level" | 162 | 3 | 9231 frames | unclear | 7801 | unclear | 1430 | DSC | Unclear | no | Qualitative results | No | n/a | SD | Graph | Unclear | none |
| Jearanai, 2023 [55] | Hold-out | n/a | unclear | Internal Retrospective, Prospective Interventional | "Yes, on frame level" | unclear | 1 | 3600 frames | unclear | 3200 | unclear | 400 | Precision, Recall, mAP | Over n frames in test set | no | Qualitative results | No | unclear | None | None | none | none |
| Jha, 2021 [56] | Hold-out | n/a | unclear | Internal Retrospective | "Yes, on frame level" | 30 | 1 | 5983 frames | unclear | 5385 | unclear | 598 | mIoU, DSC, Precision, Recall, Accuracy, Others | Unclear | no | Qualitative results | No | 101.36 FPS | None | None | none | none |
| Jin, 2020 [57] | Hold-out | n/a | 50/50 | Internal Retrospective | "Yes, on patient and frame level" | 80 | 1 | 80 videos | 40 | unclear | 40 | unclear | mAP | "Over n independent samples (i.e., one patient/video) in independent test set" | no | Qualitative results | No | n/a | SD | Exact numbers, Graph | "Over n independent samples (i.e., one patient/video) in independent test set" | none |
| Jin, 2018 [58] | Hold-out | n/a | 67/33 (class) | Internal Retrospective | Unclear | 15 | 1 | 2532 frames (bounding box), 23000 (classification) | 10 (classification) | 2026 (bounding box) | 5 (classification) | 506 (bounding box) | mAP | Unclear | no | Qualitative results, Heat map, others | No | 5 FPS | None | None | none | none |
| Kamrul Hasan, 2021 [59] | Hold-out | n/a | unclear | Internal Retrospective | "Yes, on frame level" | 30 | 1 | Semantic: 635 frames Classification: 4270 frames | unclear | Semantic: 508, Classification: 1016 | unclear | Semantic: 127, Classification: 3254 | AP, Accuracy, DSC, IoU, Sensitivity, Specificity, Others | Unclear | no | Qualitative results | No | unclear | Other | Exact numbers, Graph | Unclear | none |
| Kanakatte, 2020 [60] | Hold-out | n/a | 50/50 | Internal Retrospective | "Yes, on frame level" | 10 | 1 | 1210 frames | unclear | 960 | unclear | 250 | mAP | Unclear | no | Qualitative results | No | n/a | None | None | none | none |
| Kawamura, 2023 [61] | Hold-out | n/a | unclear | Internal Retrospective | "Yes, on patient and frame level" | 72 | 1 | 23793 frames | 52 | unclear | 20 | unclear | Precision, Recall, F1-Score, Specificity, Accuracy | Unclear | no | Class activation map | No | 6 FPS | None | None | none | none |
| Khalid, 2023 [62] | Others | n/a | unclear | External Retrospective | "Yes, on patient and frame level" | 22 | unclear | 22 videos | n/a | n/a | n/a | n/a | Others | Unclear | no | Qualitative results | No | unclear | CI | Exact numbers | Unclear | none |
| Khalid, 2023 [63] | unclear | n/a | unclear | Internal Retrospective | Unclear | unclear | 2 | unclear | unclear | unclear | unclear | unclear | Precision, Recall, F1-Score, Others | Unclear | no | Qualitative results, Depth map | No | n/a | None | None | none | none |
| Kim, 2024 [64] | Hold-out | n/a | 50/50 | Internal Retrospective | "Yes, on frame level" | unclear | 1 | 1300 frames | unclear | 1000 | unclear | 300 | IoU, Precision, Recall, mAP, Others | Unclear | no | Qualitative results | No | unclear | None | None | none | none |
| Kinoshita, 2024 [65] | Hold-out | n/a | unclear | Internal Retrospective | "Yes, on patient and frame level" | 75 | 2 | 800 frames | 70 | 740 | 5 | 60 | IoU, DSC | Unclear | no | Qualitative results | No | 60 FPS | None | None | none | Other |
| Kitaguchi, 2023 [66] | Hold-out | n/a | unclear | Prospective Interventional | "Yes, on patient and frame level" | Ureter: 344, Nerves: 249 | 1 | 25122 frames (Ureter), 18176 frames (Nerve) | 288 (Ureter), 194 (Nerve) | 20506 (Ureter), 14577 (Nerve) | 56 (Ureter) 55 (Nerve) | 4616 (Ureter), 3599 (Nerve) | Precision, Recall, DSC | Over n frames in test set | no | Qualitative results | No | 8-10 FPS | None | None | none | Other |
| Kitaguchi, 2022 [67] | Cross-Validation, Hold-out | 5 | unclear | Internal Retrospective | "Yes, on patient and frame level" | 337 | unclear | 38628 frames | unclear | unclear | unclear | unclear | mAP | Unclear | no | Qualitative results | No | unclear | SD | Exact numbers | Unclear | none |

| | | | | | | | | | | | | | | | | | | | | | | |
|---|---|---|---|---|---|---|---|---|---|---|---|---|---|---|---|---|---|---|---|---|---|---|
| Kitaguchi, 2022 [68] | Hold-out | n/a | 4 different | Internal Retrospective | "Yes, on patient and frame level" | 128 | 5 | 5238 frames | 94 | 4419 | 34 | 819 | IoU, AP, mAP | Over n frames in test set | no | Qualitative results | No | n/a | SD | Exact numbers, Graph | Over n frames in test set | none |
| Kletz, 2019 [69] | Hold-out | n/a | unclear | Internal Retrospective | "Yes, on frame level" | unclear | unclear | 333 frames | unclear | 267 | unclear | 66 | AP, Recall | Unclear | no | Qualitative results | No | n/a | None | None | none | none |
| Kolbinger, 2023 [70] | Hold-out, Cross-Validation | 4 | unclear | Internal Retrospective | "Yes, on patient and frame level" | 32 | 1 | 13195 frames | 24 | uncelar | 8 | unclear | F1-Score, IoU, Precision, Recall, Specificity | Over n frames in test set | no | Qualitative results | No | 28 ms | SD | Graph, Exact numbers | Over n frames in test set | none |
| Kolbinger, 2024 [71] | Cross-Validation | 4 | unclear | Internal Retrospective | "Yes, on patient and frame level" | 33 | 1 | 9037 frames | unclear | unclear | unclear | unclear | F1-Score, IoU, Precision, Recall, Specificity | Over n frames in test set | no | Qualitative results | No | n/a | SD | Exact numbers, Graph | Over n frames in test set | none |
| Kolbinger, 2024 [72] | Hold-out | n/a | unclear | Internal Retrospective | "Yes, on patient and frame level" | 32 | 1 | 13195 frames | 24 | uncelar | 8 | unclear | Accuracy, Precision, Recall, F1-Score, IoU, Specificity, Others | Over n frames in test set | no | no | No | n/a | None | None | none | none |
| Kondo, 2021 [73] | Cross-Validation | 4 | unclear | Internal Retrospective | "Yes, on patient and frame level" | 80 | 1 | 80 videos | 60 | unclear | 20 | unclear | Precision, Recall, F1-Score | Over n frames in test set | no | no | No | n/a | None | None | none | none |
| Konduri, 2024 [74] | Hold-out | n/a | unclear | Internal Retrospective | "Yes, on patient and frame level", "Yes, on frame level" | unclear | unclear | unclear | unclear | unclear | unclear | unclear | Accuracy, Precision, Recall, F1-Score | Unclear | no | no | No | n/a | None | None | none | none |
| Kong, 2021 [75] | Cross-Validation | 4 | unclear | Internal Retrospective | "Yes, on patient and frame level" | 2 | 1 | 2760 frames | unclear | unclear | unclear | unclear | IoU, DSC, Precision, Recall | "Over n independent samples (i.e., one patient/video) in independent test set" | no | Qualitative results | No | n/a | SD | Exact numbers | "Over n independent samples (i.e., one patient/video) in independent test set" | none |
| Kumazu, 2021 [76] | Hold-out | n/a | 60/10/30 | Internal Retrospective | "Yes, on patient and frame level" | 33 | 1 | 1880 frames | 23 | 1800 | 10 | 80 | Recall, F1-Score, DSC | Over n frames in test set | no | Qualitative results | No | 30 FPS | SD | Exact numbers | Over n frames in test set | Other |
| Kumazu, 2025 [77] | Hold-out | n/a | 60/10/30 | Prospective Observational | "Yes, on patient and frame level" | 93 | unclear | 31880 frames | unclear | unclear | unclear | unclear | DSC | Over n frames in test set | no | Qualitative results | No | unclear | SD | Exact numbers | Over n frames in test set | none |
| Labrunie, 2022 [78] | Cross-Validation | 10 | unclear | Internal Retrospective | "Yes, on patient and frame level" | 68 | unclear | 1415 frames | unclear | unclear | unclear | unclear | Others | Unclear | no | others | No | n/a | Other | Exact numbers | Unclear | none |
| Lam, 2022 [79] | Hold-out | n/a | unclear | Prospective Observational | "Yes, on frame level" | 5 | 1 | 2600 frames | 3 | 2600 | 2 | unclear | mAP, F1-Score | Over n frames in test set | no | Qualitative results, others | No | n/a | SD | Exact numbers | Over n frames in test set | none |
| Laplante, 2022 [80] | Cross-Validation | 10 | unclear | External Retrospective | "Yes, on patient and frame level" | 333 | 143 | 333 videos | 308 | unclear | 25 | 47 | IoU, F1-Score, DSC | Unclear | no | Qualitative results, Probability map | No | unclear | SD | Exact numbers | Unclear | none |
| Lavanchy, 2021 [81] | Cross-Validation | 10 | unclear | Internal Retrospective | "Yes, on frame level" | 242 | 1 | 13823 frames | unclear | 10935 | unclear | 2888 | AP, Recall | Unclear | no | Qualitative results | No | n/a | SD | Exact numbers | Unclear | "Frequentist parametric (e.g., t-test)" |
| Le, 2023 [82] | Hold-out | n/a | unclear | Internal Retrospective | "Yes, on patient and frame level" | 15 | 1 | 2811 frames | unclear | 1968 | unclear | 843 | Precision, Recall, mAP, IoU | Over n frames in test set | no | Qualitative results | No | n/a | None | None | none | none |
| Lee, 2024 [83] | Hold-out | n/a | unclear | Internal Retrospective | Unclear | 47 | 2 | 18120 frames | n/a | n/a | n/a | n/a | mIoU, AP | Unclear | no | Qualitative results | No | n/a | None | None | none | none |
| Leifman, 2024 [84] | Hold-out | n/a | 80/20 | Prospective Observational, Internal Retrospective | "Yes, on patient and frame level" | 740 | 6 | 740 videos | 560 | unclear | 180 | unclear | Confusion Matrix, Others, Sensitivity, Specificity | Unclear | no | Qualitative results | No | 49.2 ms | CI | Exact numbers | Unclear | none |
| Leifmann, 2022 [85] | Hold-out | n/a | unclear | Internal Retrospective | "Yes, on patient and frame level", "Yes, on frame level" | 30 | 1 | 12764 frames (incl. non-human) | unclear | unclear | unclear | unclear | IoU, DSC | Unclear | no | Qualitative results | No | n/a | None | None | none | none |
| Li, 2025 [86] | Hold-out | n/a | unclear | Internal Retrospective | Unclear | 2200 | unclear | 901000 frames | unclear | unclear | unclear | unclear | Precision, Recall, mAP | Unclear | no | no | No | 100.8 ms | None | None | none | none |
| Li, 2023 [87] | Hold-out | n/a | unclear | External Retrospective | "Yes, on frame level", "Yes, on patient and frame level" | 117 | 1 | 14080 frames | 17 | 8080 | 100 | 6000 | IoU, DSC | Unclear | no | Qualitative results | No | n/a | None | None | none | none |
| Liao, 2024 [88] | Hold-out | n/a | unclear | Internal Retrospective | "Yes, on patient and frame level", "Yes, on frame level" | 80 | 1 | 80 videos | 63 (incl. 5296 TOP clips) | unclear | 17 | 3000 | Accuracy, IoU, Others | Unclear | no | Qualitative results, Class activation map | No | n/a | None | None | none | none |
| Lin, 2024 [89] | Hold-out | | unclear | Internal Retrospective | "Yes, on frame level" | unclear | 1 | 14480 frames | 145 snips | unclear | 36 snips | unclear | mAP | Unclear | no | Qualitative results | No | 51 ms | None | None | none | "Frequentist non-parametric (e.g., Mann-Whitney)" |
| Lin, 2021 [90] | Cross-Validation | 3 | 75/12.5/12.5 | Internal Retrospective | "Yes, on patient and frame level" | 8 | 1 | 10040 frames | 7 | unclear | 1 | unclear | DSC, IoU | Unclear | no | Qualitative results | No | 6.4 ms | SD | Exact numbers | Unclear | none |

| | | | | | | | | | | | | | | | | | | | | | | |
|---|---|---|---|---|---|---|---|---|---|---|---|---|---|---|---|---|---|---|---|---|---|---|
| Liu, 2022 [91] | Hold-out | n/a | 32/4/5 | External Retrospective | "Yes, on patient and frame level" | 41 | 1 | 5533 frames | unclear | 4980 | unclear | 553 | Precision, Recall, IoU, AP | Unclear | no | no | No | n/a | None | None | none | Other |
| Liu, 2024 [92] | Hold-out | n/a | unclear | Internal Retrospective | "Yes, on frame level" | unclear | unclear | 3000 frames | unclear | 2250 | unclear | 750 | mIoU, Others | Unclear | no | Qualitative results | No | 29.14 FPS | None | None | none | none |
| Liu, 2022 [93] | Hold-out | n/a | 50/30/20 | Internal Retrospective | "Yes, on patient and frame level" | 80 | 1 | 80 videos | 40 | unclear | 40 | unclear | mAP | Other | no | Qualitative results, Heat map | No | 50 FPS, 20 ms | SD, Standard Error | Graph | Other | none |
| Lou, 2023 [94] | Hold-out, unclear | n/a | unclear | Internal Retrospective | "Yes, on frame level" | 29 | 1 | 816 frames | unclear | 662 | unclear | 154 | DSC, IoU, F1-Score | Unclear | no | Qualitative results | No | 40 FPS | Other | Exact numbers | Unclear | none |
| Loukas, 2020 [95] | Cross-Validation | 10 | unclear | Internal Retrospective | "Yes, on frame level" | unclear | 1 | 482 frames | unclear | unclear | unclear | unclear | Accuracy | Unclear | no | Qualitative results | No | n/a | SD, Other | Exact numbers | Unclear | "Frequentist parametric (e.g., t-test)" |
| Loza, 2024 [96] | Hold-out | n/a | 50/30/20 | Internal Retrospective | "Yes, on frame level" | 15 | 1 | 2532 frames | unclear | 2248 | unclear | 563 | mAP | Over n frames in test set | no | Qualitative results | No | 36 FPS | None | None | none | none |
| Maack, 2024 [97] | Hold-out | n/a | unclear | Internal Retrospective | "Yes, on patient and frame level" | 32 | 1 | 13195 frames | 24 | uncelar | 8 | unclear | mDSC | Unclear | no | Qualitative results | No | n/a | None | None | none | none |
| Madad Zadeh, 2020 [98] | Hold-out | n/a | unclear | Internal Retrospective | "Yes, on patient and frame level" | 8 | 1 | 461 frames | unclear | 361 | unclear | 100 | IoU, Accuracy | Unclear | no | Qualitative results | No | n/a | None | None | none | none |
| Madani, 2022 [99] | Cross-Validation | 10 | unclear | External Retrospective | "Yes, on frame level" | 290 | 136 | 2627 frames | unclear | unclear | unclear | unclear | F1-Score, IoU, Accuracy, Sensitivity, Specificity, NPV, PPV | Over frame-level averages of n cross-validation folds | no | Qualitative results, Heat map, Probability map | No | <10 ms | SD | Exact numbers | Over frame-level averages of n cross-validation folds | Other |
| Maqbool, 2020 [100] | Hold-out | n/a | unclear | Internal Retrospective | "Yes, on patient and frame level" | 2 | 1 | 307 frames | unclear | 245 | unclear | 62 | Precision, Recall, F1-Score, IoU | Unclear | no | Qualitative results | No | n/a | None | None | none | none |
| Marullo, 2023 [101] | Hold-out | n/a | unclear | Internal Retrospective | "Yes, on patient and frame level", "Yes, on frame level" | 26 | 1 | 318 frames | unclear | 232 | unclear | 86 | Accuracy, DSC | Over n frames in test set | no | Qualitative results | No | 15 FPS | None | None | none | none |
| Mascagni, 2022 [102] | Cross-Validation | 5 | 60/20/20 | Internal Retrospective | Unclear | 200 | 1 | 2854 frames | unclear | unclear | unclear | unclear | IoU, AP | Unclear | no | Qualitative results, Saliency Map | No | n/a | SD | Exact numbers | Unclear | none |
| Matsumoto, 2024 [103] | unclear | n/a | 66/34 | Internal Retrospective | "Yes, on frame level" | 18 | 1 | 18 Videos | 6 | 1080 | 12 | unclear | DSC, Accuracy | Unclear | no | Qualitative results | No | unclear | None | None | none | "Frequentist non-parametric (e.g., Mann-Whitney)" |
| Mehta, 2024 [104] | Hold-out | n/a | 80/10/10 | Internal Retrospective | "Yes, on patient and frame level" | 1107 | 25 | 65000 frames | 997 | unclear | 110 | unclear | F1-Score, Precision, Recall, DSC | Over sample-level averages of n cross-validation folds | no | Qualitative results, others | No | 12.5 FPS | SD | Exact numbers | Over sample-level averages of n cross-validation folds | none |
| Mishra, 2017 [105] | Hold-out | n/a | 67/33 | Internal Retrospective | "Yes, on patient and frame level" | 15 | 1 | 15 Videos | 10 | unclear | 5 | unclear | Accuracy | Unclear | no | Qualitative results | No | 2.42 ms | None | None | none | none |
| Murali, 2024 [106] | Hold-out | n/a | unclear | Internal Retrospective | Unclear | 50 | 1 | 493 frames | 40 | unclear | 10 | unclear | mIoU, mDSC | Unclear | no | no | No | n/a | None | None | none | none |
| Murali, 2023 [107] | Hold-out | n/a | unclear | Internal Retrospective | "Yes, on patient and frame level" | 201 | 1 | 11090 frames | unclear | unclear | unclear | unclear | mAP | Unclear | no | no | No | n/a | SD | Exact numbers | Unclear | none |
| Murali, 2022 [108] | Hold-out | n/a | 60/20/20 | Internal Retrospective | "Yes, on patient and frame level" | 201 | 1 | 11090 frames | 161 | unclear | 40 | unclear | mAP, DSC | Unclear | no | Qualitative results | No | | SD | Graph | Unclear | none |
| Myo, 2024 [109] | Hold-out | n/a | 80/20 | Prospective Observational | "Yes, on frame level" | 30 | 1 | 10040 frames | unclear | 5983 | unclear | 4057 | Precision, Recall, F1-Score, DSC, mAP | Over sample-level averages of n cross-validation folds | no | Qualitative results | No | 16.67 ms (60 FPS) | None | None | none | none |
| Nakanuma, 2023 [110] | Hold-out | n/a | unclear | Prospective Interventional | "Yes, on patient and frame level" | 10 | 1 | unclear | unclear | unclear | unclear | unclear | DSC, Others | Unclear | no | Qualitative results | No | 0.09s time lag | None | None | none | "Frequentist non-parametric (e.g., Mann-Whitney)" |
| Namazi, 2022 [111] | Hold-out | n/a | unclear | Internal Retrospective | "Yes, on frame level", "Yes, on patient and frame level" | 80 | 1 | 95 videos | 50 | unclear | 45 | unclear | Accuracy, F1-Score, Precision, Recall | Unclear | no | Class activation map | No | n/a | None | None | none | none |
| Nema , 2023 [112] | Hold-out | n/a | 67/33 | Internal Retrospective | "Yes, on frame level" | 6 | 1 | 9195 frames | 4 | 4560 | 2 | 4635 | IoU, Others, Specificity | Over n frames in test set | no | Qualitative results | No | n/a | None | None | none | none |
| Nwoye, 2024 [113] | Hold-out | n/a | 50/10/40 | Internal Retrospective | "Yes, on patient and frame level" | 20 | 1 | 35000 frames | unclear | unclear | unclear | unclear | Others | Unclear | „consistency accuracy metric | Qualitative results | No | 45828 | None | None | none | none |
| Nwoye, 2019 [114] | Hold-out | n/a | 50/12.5/37,5 | Internal Retrospective | "Yes, on patient and frame level" | 80 | 1 | 80 videos | 50 | unclear | 30 | unclear | Others | Unclear | no | Qualitative results, Heat map | No | 25 FPS | Other | Exact numbers | Unclear | none |
| Oh, 2024 [115] | Cross-Validation | 5 | 24/6/5 | Prospective Observational | "Yes, on patient and frame level" | 35 | 1 | 300 frames | 30 | 300 | 5 | 50 | Recall, Precision, DSC | Over frame-level averages of n cross-validation folds | no | Qualitative results, Heat map | Fluorescence Imaging | 15.3 FPS | SD | Exact numbers | Over frame-level averages of n cross-validation folds | none |
| Owen, 2022 [116] | Hold-out | n/a | 60/20/20 | Internal Retrospective | "Yes, on patient and frame level" | 232 | unclear | 6732 frames | 150 | 6122 | 15 | 610 | IoU, Precision, Recall, F1-Score | Over n frames in test set | no | Qualitative results | No | n/a | None | None | none | none |
| Ozbulak, 2025 [117] | Others | n/a | n/a | Internal Retrospective | No | 6 | 1 | 3600 frames | n/a | n/a | 6 | unclear | IoU | Other | no | no | No | 25 FPS | None | None | none | none |

| | | | | | | | | | | | | | | | | | | | | | | |
|---|---|---|---|---|---|---|---|---|---|---|---|---|---|---|---|---|---|---|---|---|---|---|
| Pan, 2023 [118] | Hold-out | n/a | unclear | Internal Retrospective | "Yes, on frame level" | 36 | 2 | 4468 frames | unclear | unclear | unclear | unclear | mAP, Precision, Recall | Unclear | no | Qualitative results | No | n/a | None | None | none | none |
| Penza, 2018 [119] | Others | n/a | n/a | Internal Retrospective | Unclear | unclear | 1 | unclear | unclear | unclear | unclear | unclear | Precision, Recall, F1-Score | Unclear | no | Qualitative results | No | 1.6 FPS | None | None | none | none |
| Pradeep, 2022 [120] | Hold-out | n/a | unclear | Internal Retrospective | Unclear | 80 | 1 | 80 videos | unclear | 2025 | unclear | unclear | mAP | Unclear | no | Qualitative results | No | n/a | None | None | none | none |
| Prokopetc, 2015 [121] | Cross-Validation | 7 | unclear | Internal Retrospective | "Yes, on patient and frame level" | 38 | unclear | 195 frames | unclear | unclear | unclear | unclear | Precision, Recall | Unclear | no | Qualitative results | No | n/a | SD | Exact numbers | Unclear | none |
| Protserov, 2024 [122] | Hold-out | n/a | 70/15/15 + 2. Dataset for testing again | Prospective Observational | "Yes, on patient and frame level" | 314 | 143 | 2674 frames | unclear | unclear | unclear | unclear | Precision, Recall, DSC | Over n frames in test set | no | Qualitative results, Heat map | No | 70 ms | SD | Exact numbers | Over n frames in test set | none |
| Rahbar, 2020 [123] | Hold-out | n/a | unclear | Internal Retrospective | "Yes, on patient and frame level" | 15 | unclear | unclear | 5 | unclear | 10 | unclear | Others | "Over n independent samples (i.e., one patient/video) in independent test set" | no | others | No | 0.1245 milliseconds per frame | SD, Other | Exact numbers | "Over n independent samples (i.e., one patient/video) in independent test set" | none |
| Rahbar, 2024 [124] | unclear | n/a | unclear | Internal Retrospective | Unclear | unclear | unclear | unclear | unclear | unclear | unclear | unclear | mIoU, mDSC | Over n frames in test set | no | Qualitative results | No | 30.2 ms | None | None | none | none |
| Raja, 2024 [125] | Hold-out | n/a | unclear | Internal Retrospective | Unclear | 10 | 1 | 2811 frames | unclear | 2248 | unclear | 563 | Precision, Recall, mAP | Over n frames in test set | no | Qualitative results | No | 2.5 ms | None | None | none | none |
| Ryu, 2024 [126] | Cross-Validation | 5 | unclear | Internal Retrospective | "Yes, on frame level" | 104 | 1 | 2624 frames | unclear | unclear | unclear | unclear | DSC, Precision, Recall | Unclear | no | Qualitative results | No | 80 ms | None | None | none | none |
| Sahu, 2017 [127] | Hold-out | n/a | unclear | Internal Retrospective | "Yes, on patient and frame level" | 15 | 1 | 15 videos | 10 | unclear | 5 | unclear | AP, mAP | Over n frames in test set | no | no | No | n/a | None | None | none | none |
| Samuel, 2021 [128] | unclear | n/a | unclear | Internal Retrospective | "Yes, on patient and frame level" | 80 | 1 | unclear | unclear | unclear | unclear | 10383 | Recall, Precision, F1-Score | Over n frames in test set | no | no | No | 25 ms | None | None | none | none |
| Sanchez-Matilla, 2022 [129] | Cross-Validation | 5 | unclear | Internal Retrospective | "Yes, on patient and frame level" | 17 | 1 | 8080 frames | 14 | unclear | 3 | unclear | Accuracy, F1-Score, mIoU, mDSC, Others | Over frame-level averages of n cross-validation folds | no | no | No | n/a | SD | Exact numbers | Over frame-level averages of n cross-validation folds | none |
| Satyanaik, 2024 [130] | Hold-out | n/a | 60/20/20 | Internal Retrospective | "Yes, on patient and frame level" | 271 | 2 | 271 videos | 216 | unclear | 55 | unclear | mAP | Unclear | no | no | No | n/a | SD | Exact numbers | Unclear | none |
| Satyanaik, 2024 [131] | Hold-out | n/a | unclear | Internal Retrospective | "Yes, on patient and frame level" | 271 | 2 | 18780 frames | 216 | unclear | 55 | unclear | mAP | Unclear | no | no | No | n/a | SD | Exact numbers | Unclear | none |
| Seenivasan, 2023 [132] | Hold-out | n/a | unclear | Internal Retrospective | "Yes, on patient and frame level" | unclear | 2 | unclear | unclear | unclear | unclear | unclear | Accuracy, Recall, F1-Score | Unclear | no | Qualitative results | No | n/a | None | None | none | none |
| Sengun, 2023 [133] | Cross-Validation | 5 | 70/15/15 | Internal Retrospective | "Yes, on patient and frame level" | 40 | 1 | 2000 frames | 34 | 1700 | 6 | 300 | IoU, mDSC, PPV, Sensitivity | Unclear | no | Qualitative results | No | unclear | SD | Exact numbers | Unclear | none |
| Sengun, 2024 [134] | Cross-Validation | 5 | 80/20/x | Internal Retrospective | "Yes, on patient and frame level" | 20 | 1 | 1000 frames | 16 | 800 | 4 | 200 | Accuracy, Sensitivity, Specificity, mIoU, mDSC | Over frame-level averages of n cross-validation folds | no | Qualitative results, Probability Map | No | n/a | None | None | none | none |
| Shen, 2023 [135] | Hold-out | n/a | unclear | Internal Retrospective | "Yes, on frame level" | unclear | 1 | 3000 frames | unclear | 2250 | unclear | 750 | mIoU, mDSC | Over n frames in test set | no | Qualitative results | No | 9.78 ms | None | None | none | none |
| Sheng, 2024 [136] | unclear | n/a | unclear | Internal Retrospective | Unclear | 17 | 1 | 8080 frames | unclear | unclear | unclear | unclear | mIoU | Unclear | no | others, Saliency Map | No | n/a | SD | Exact numbers | Unclear | none |
| Shi, 2020 [137] | Hold-out | n/a | unclear | Internal Retrospective | "Yes, on frame level" | 80 | 1 | 4011 frames, Frames from Cholec80 | unclear | 3289 | unclear | 722 | mAP | Over n frames in test set | no | Qualitative results | No | 55.5 FPS | None | None | none | none |
| Shimgekar, 2021 [138] | Hold-out | n/a | unclear | Internal Retrospective | Unclear | unclear | unclear | 242 frames | unclear | 220 | unclear | 22 | IoU | Unclear | no | Qualitative results | Other | n/a | None | None | none | none |
| Silva, 2022 [139] | Hold-out | n/a | unclear | Internal Retrospective | "Yes, on patient and frame level" | 17 | 1 | 8080 frames | 15 | unclear | 2 | unclear | Precision, Recall, IoU, DSC | Over n frames in test set | no | Qualitative results | No | n/a | None | None | none | none |
| Smithmaitrie, 2024 [140] | Hold-out | n/a | unclear | Prospective Interventional | "Yes, on patient and frame level", "Yes, on frame level" | 40 | 1 | 3200 frames | unclear | 2560 | unclear | 640 | AP, Precision, Recall | Over n frames in test set | no | Qualitative results | No | 32.90 ms | None | None | none | none |
| Sonsilphong, 2022 [141] | Others | n/a | unclear | Prospective Interventional | "Yes, on patient and frame level" | 5 | unclear | unclear | 5 | unclear | cadaver | unclear | Accuracy | Unclear | no | Qualitative results | No | 20 FPS | None | None | none | none |
| Streckert, 2023 [142] | Hold-out | n/a | unclear | Internal Retrospective | "Yes, on frame level" | 6 | 1 | 300 frames | unclear | 160 | unclear | 140 | IoU | Over n frames in test set | no | Qualitative results | No | n/a | None | None | none | none |
| Strong, 2024 [143] | Hold-out | n/a | unclear | Internal Retrospective | "Yes, on frame level" | 55 | 1 | 1383 frames | unclear | 1234 | unclear | 149 | IoU, Accuracy | Over n frames in test set | no | Qualitative results | No | n/a | SD | Exact numbers | Over n frames in test set | "Frequentist non-parametric (e.g., Mann-Whitney)" |
| Sun, 2022 [144] | Hold-out | n/a | unclear | Internal Retrospective | "Yes, on patient and frame level" | unclear | unclear | 8000 frames | 10 | 5000 | 10 | 3000 | Accuracy | Unclear | no | Qualitative results | No | 53 FPS | None | None | none | none |

| | | | | | | | | | | | | | | | | | | | | | | |
|---|---|---|---|---|---|---|---|---|---|---|---|---|---|---|---|---|---|---|---|---|---|---|
| Takeuchi, 2023 [145] | Hold-out | n/a | 81/19 | Prospective Observational | "Yes, on patient and frame level" | 160 | 1 | 2656 frames | 130 | 2356 | 30 | 300 | mAP, Accuracy, F1-Score, Sensitivity, Specificity, Precision | Over n frames in test set | no | Qualitative results | No | unclear | None | None | none | Other |
| Tao, 2023 [146] | Hold-out | n/a | unclear | Internal Retrospective | "Yes, on patient and frame level" | 121 | 1 | 121 videos | 67 | unclear | 54 | unclear | mAP | Other | no | no | No | 23.8 FPS | SD | Exact numbers | Other | none |
| Tokuyasu, 2021 [147] | Hold-out | n/a | unclear | Prospective Interventional | "Yes, on patient and frame level" | 100 | 1 | 2339 frames | 76 | 2145 | 23 | 194 | AP | Unclear | no | Qualitative results | No | 37.2 FPS | None | None | none | none |
| Urrea, 2024 [148] | Cross-Validation | unclear | unclear | Internal Retrospective | "Yes, on frame level" | 17 | 1 | 8080 frames | unclear | 6464 | unclear | 1616 | Accuracy, IoU, DSC, F1-Score, Recall, Precision | Unclear | no | Qualitative results | No | 56 ms | None | None | none | none |
| Valderrama, 2022 [149] | Cross-Validation | 2 | unclear | Internal Retrospective | "Yes, on patient and frame level" | 8 | 1 | 2238 frames (bounding box) | unclear | unclear | unclear | unclear | mAP | Unclear | no | Qualitative results | No | n/a | SD | Exact numbers | Unclear | none |
| Vardazaryan, 2018 [150] | Hold-out | n/a | 50/12.5/37,5 | Internal Retrospective | "Yes, on patient and frame level" | 80 | 1 | 80 videos | 50 | unclear | 30 | unclear | mAP | Unclear | no | Qualitative results, Localisation Map | No | unclear | None | None | none | none |
| Wagner, 2023 [151] | Hold-out | n/a | 73/27 | Internal Retrospective | "Yes, on patient and frame level" | 33 | 3 | 33 videos | 24 | unclear | 9 | unclear | F1-Score | "Over n independent samples (i.e., one patient/video) in independent test set" | no | no | No | n/a | None | None | none | Other |
| Wang, 2023 [152] | Hold-out | n/a | unclear | Internal Retrospective | "Yes, on frame level" | 20 | 1 | 2798 frames | unclear | 2518 | unclear | 280 | mAP, Precision, Recall | Over n frames in test set | no | Qualitative results | No | 37.049 FPS | None | None | none | none |
| Wang, 2017 [153] | Cross-Validation | 5 | unclear | Internal Retrospective | "Yes, on patient and frame level" | 15 | 1 | 15 Videos | unclear | unclear | unclear | unclear | mAP | Over n frames in test set | no | no | No | n/a | None | None | none | none |
| Wang, 2024 [154] | Hold-out | n/a | unclear | Internal Retrospective | "Yes, on patient and frame level", "Yes, on frame level" | 95 | 1 | 187309 Keyframes | 40 + unclear | 88551 | 40 + unclear | 98758 | mAP | Over n frames in test set | no | no | No | n/a | None | None | none | none |
| Wang, 2023 [155] | Hold-out | n/a | 80/20 | Internal Retrospective | "Yes, on patient and frame level" | 50 | 1 | 1372 frames | unclear | 872 | unclear | 500 | IoU, Accuracy | Over n frames in test set | no | Qualitative results | No | n/a | SD | Exact numbers, Graph | Over n frames in test set | none |
| Wang, 2025 [156] | Cross-Validation | 2 | 60/40 | Internal Retrospective | Unclear | 13 | 1 | unclear | 8 | unclear | 5 | unclear | IoU, AP, DSC, Others | Unclear | TSC (only for EV18) | Qualitative results | No | 219 ms | None | None | none | none |
| Wang, 2021 [157] | Hold-out | n/a | unclear | Internal Retrospective | "Yes, on patient and frame level" | 15 | 1 | 15 videos | 10 | 2532 | 5 | unclear | Recall, Precision, mAP | Over n frames in test set | no | Qualitative results | No | 40.2 FPS | None | None | none | none |
| Wang, 2020 [158] | Hold-out | n/a | unclear | External Retrospective | "Yes, on frame level" | 35 | 2 | 5696 frames | unclear | 4557 | unclear | 1139 | mAP | Unclear | no | Heat map | No | 32.3 FPS | None | None | none | none |
| Wang, 2024 [159] | Hold-out | n/a | unclear | Internal Retrospective | "Yes, on patient and frame level" | 81 | 2 | 81 videos | unclear | unclear | unclear | 3645 | IoU, AP | Over n frames in test set | no | Qualitative results, Class activation map | No | unclear | None | None | none | none |
| Wang, 2023 [160] | Hold-out | n/a | unclear | Internal Retrospective | "Yes, on frame level" | 15 | 1 | 2532 frames | unclear | 2026 | unclear | 506 | mAP, Precision, Recall, F1-Score | Over n frames in test set | no | Qualitative results | No | 50 FPS | None | None | none | none |
| Ward, 2022 [161] | Cross-Validation | 10 | 90/x/10 | Internal Retrospective | "Yes, on patient and frame level" | 200 | 1 | 200 Videos | 180 | unclear | 20 | unclear | Others, Confusion Matrix | Unclear | no | others | No | n/a | Other | Exact numbers, Graph | Unclear | Unclear |
| Wei, 2024 [162] | Cross-Validation | unclear | 80/20 | Internal Retrospective | "Yes, on patient and frame level" | 30 | 1 | 10040 frames | 24 | 7160 | 6 | 2880 | DSC, Others | Unclear | no | Qualitative results | No | n/a | None | None | none | none |
| Wei, 2023 [163] | Hold-out | n/a | unclear | Internal Retrospective | "Yes, on frame level" | 30 | 1 | 30 videos | unclear | 23600 | unclear | 4057 | DSC, Others | Unclear | no | Qualitative results | No | n/a | None | None | none | none |
| Wu, 2024 [164] | Hold-out | n/a | unclear | Internal Retrospective | "Yes, on frame level" | 21 | 1 | 1800 frames | unclear | 1440 | unclear | 360 | DSC, Accuracy, Specificity, Sensitivity | Over n frames in test set | no | Qualitative results | No | 26.32 ms | None | None | none | none |
| Xi, 2022 [165] | Hold-out | n/a | unclear | Internal Retrospective | "Yes, on patient and frame level" | 45 | 1 | 90489 frames | 35 | unclear | 10 | unclear | AP, mAP | "Over n independent samples (i.e., one patient/video) in independent test set" | no | Qualitative results | No | n/a | None | None | none | none |
| Xu, 2024 [166] | Hold-out | n/a | unclear | Internal Retrospective | "Yes, on patient and frame level" | 22 | 1 | 11480 (incl. non-human) | 17 | 4710 | 5 | 1138 | IoU, DSC, Others | Over n frames in test set | no | Qualitative results, others | No | 15.2 ms | SD | Exact numbers | Over n frames in test set | none |
| Xue, 2022 [167] | Hold-out | n/a | unclear | Internal Retrospective | "Yes, on frame level" | 15 | 1 | 29533 frames | unclear | unclear | unclear | unclear | mAP | Over n frames in test set | no | Qualitative results | No | 29.27 ms | None | None | none | none |

| | | | | | | | | | | | | | | | | | | | | | | |
|---|---|---|---|---|---|---|---|---|---|---|---|---|---|---|---|---|---|---|---|---|---|---|
| Yamazaki, 2020 [168] | Hold-out | n/a | unclear | Internal Retrospective | "Yes, on patient and frame level" | 62 | 1 | 13833 frames | 52 | 10716 | 10 | 3117 | IoU, Precision, Recall | Over n frames in test set | no | Qualitative results | No | n/a | None | None | none | none |
| Yamlahi, 2023 [169] | Cross-Validation | 5 | unclear | Internal Retrospective | "Yes, on patient and frame level" | 45 | 1 | 90489 frames | 40 | unclear | 5 | unclear | Accuracy, mAP | Unclear | no | others | No | n/a | None | None | none | none |
| Yang, 2024 [170] | Hold-out | n/a | unclear | Internal Retrospective | "Yes, on frame level" | 80 | 1 | 1510 frames | unclear | 1208 | unclear | 302 | Precision, Recall, mAP | Unclear | no | Qualitative results | No | unclear | None | None | none | none |
| Yang, 2023 [171] | Hold-out | n/a | unclear | Internal Retrospective | "Yes, on frame level" | 17 | 1 | 8080 frames | unclear | 6464 | unclear | 1616 | DSC | Over n frames in test set | no | Qualitative results, Uncertainty Map | No | 52 ms | SD, Other | Exact numbers, Graph | Over n frames in test set | none |
| Yang, 2021 [172] | Cross-Validation | 3 | unclear | Internal Retrospective | "Yes, on frame level" | 67 | 1 | 67 videos | unclear | unclear | unclear | unclear | mDSC, Accuracy | Unclear | no | Heat map | No | n/a | None | None | None | none |
| Yin, 2024 [173] | Hold-out | n/a | 80/20/x | Internal Retrospective | "Yes, on frame level" | unclear | 1 | unclear | unclear | unclear | unclear | unclear | Accuracy, AP | Over n frames in test set | no | no | No | n/a | None | None | none | none |
| Yuan, 2024 [174] | Hold-out | n/a | unclear | Internal Retrospective | "Yes, on patient and frame level" | 45 | 1 | unclear | 40 | unclear | 5 | unclear | Precision, Recall, DSC | Unclear | no | no | No | n/a | None | None | none | none |
| Zhang, 2024 [175] | Hold-out | n/a | 85/15/x | Internal Retrospective | "Yes, on patient and frame level" | 17 | 1 | 4298 frames | 13 | 3246 | 4 | 1052 | Accuracy, Others | Unclear | no | Qualitative results, Class activation map | No | n/a | SD | Exact numbers | Unclear | none |
| Zhang, 2023 [176] | Hold-out | n/a | unclear | Prospective Interventional | "Yes, on frame level" | unclear | 1 | 2000 frames | unclear | 1800 | unclear | 200 | Precision, Recall, mAP | Over n frames in test set | no | Qualitative results | No | unclear | None | None | none | none |
| Zhang, 2023 [177] | Hold-out | n/a | 50/50 | Internal Retrospective | "Yes, on patient and frame level" | 80 | 1 | 80 videos | 40 | unclear | 40 | unclear | mAP | "Over n independent samples (i.e., one patient/video) in independent test set" | no | others | No | n/a | SD | Exact numbers | "Over n independent samples (i.e., one patient/video) in independent test set" | "Frequentist non-parametric (e.g., Mann-Whitney)" |
| Zhang, 2020 [178] | Hold-out | n/a | unclear | External Retrospective | "Yes, on frame level" | 35 | 2 | 5975 frames | unclear | 4780 | unclear | 1195 | mAP | Unclear | no | Qualitative results, Heat map | No | unclear | None | None | none | none |
| Zhang, 2023 [179] | unclear | unclear | 67/17/16 | Internal Retrospective | "Yes, on patient and frame level" | 12 | 1 | unclear | 10 | unclear | 2 | unclear | mAP | Unclear | no | Qualitative results | No | 10-35 ms | Other | Exact numbers | Unclear | none |
| Zhang, 2020 [180] | Hold-out | n/a | unclear | Internal Retrospective | "Yes, on frame level" | 15 | 1 | 1000 Frames | unclear | unclear | unclear | unclear | Sensitivity, DSC, Specificity, Others | Over n frames in test set | no | Qualitative results | No | 15 ms | None | None | none | none |
| Zhao, 2021 [181] | Hold-out | n/a | unclear | External Retrospective | "Yes, on patient and frame level" | unclear | 1 | unclear | unclear | unclear | unclear | unclear | IoU, DSC | "Over n independent samples (i.e., one patient/video) in independent test set" | no | Qualitative results | No | 300 ms | None | None | none | none |
| Zhao, 2023 [182] | Hold-out | n/a | unclear | Internal Retrospective | "Yes, on frame level" | 15 | 1 | 5199 frames | unclear | 4679 | unclear | 520 | AP | Unclear | no | Qualitative results | No | 68.5 FPS | None | None | none | none |
| Zhao, 2021 [183] | Hold-out | n/a | unclear | External Retrospective | "Yes, on patient and frame level" | 8 | 1 | unclear | unclear | unclear | unclear | unclear | IoU, DSC | "Over n independent samples (i.e., one patient/video) in independent test set" | no | Qualitative results | No | 4 FPS | None | None | none | none |
| Zhao, 2019 [184] | Hold-out | n/a | unclear | External Retrospective, Internal Retrospective | Unclear | unclear | unclear | 9376 frames | unclear | unclear | unclear | unclear | Accuracy | Unclear | no | Qualitative results | No | 33.19 | SD | Exact numbers | Unclear | none |
| Zheng, 2023 [185] | Hold-out | n/a | unclear | External Retrospective | "Yes, on patient and frame level" | 132 | 2 | 1888 frames | unclear | 1500 | unclear | 388 | Precision, F1-Score, IoU | Unclear | no | Qualitative results | No | < 10 ms | None | None | none | Other |
| Zhou, 2024 [186] | Cross-Validation | 5 | unclear | Internal Retrospective | Unclear | 55 | 1 | 531 frames | unclear | unclear | unclear | unclear | IoU, DSC | Unclear | no | Qualitative results, others | No | n/a | None | None | none | none |
| Zhou, 2022 [187] | Hold-out | n/a | 67/33 | Internal Retrospective | "Yes, on patient and frame level" | 15 | 1 | 2532 frames | 10 | unclear | 5 | unclear | Precision, Recall, mAP | Unclear | no | Qualitative results | No | 102.7 FPS | None | None | none | none |
| Zygomalas, 2024 [188] | Hold-out | n/a | unclear | Internal Retrospective | "Yes, on frame level" | 25 | unclear | 1095 frames | unclear | 800 | unclear | 295 | F1-Score, mAP | Unclear | no | Qualitative results | No | n/a | None | None | none | none |

Abbreviations: *n/a: not applicable, SD: standard deviation, CI: confidence interval, IQR: Interquartile Range*

**Footnotes:**

a. Data Split into Training/Validation/Testing: Only if specified based on the number of patients or videos per patient.
b. Indicates whether the model was evaluated on data from the same dataset as used for training (Internal Retrospective), on data from different institutions or datasets (External Retrospective), on prospectively collected data (Prospective Observational), or in clinical testing (Prospective Interventional).

c. Reported numbers of the data used throughout the entire study.
d. If the number of images was specified, it was reported as such; otherwise, the number of videos was provided.
e. Possible options for the calculation of performance metrics as well as model uncertainty and prediction variability: Over n independent samples (i.e., one patient/video) in independent test set (i.e., final models were tested on independent test set. Metrics were aggregated within one sample, if applicable (i.e., all frames of one video). Metrics/metric aggregates were aggregated over all samples in the test set)
   - Over n independent samples (i.e., one patient/video) in independent test set (i.e., final models were tested on independent test set. Metrics were aggregated within one sample, if applicable (i.e., all frames of one video). Metrics/metric aggregates were aggregated over all samples in the test set)
   - Over n independent samples across all cross-validation folds (i.e., models were cross-validated on CV folds split along patient/video lines, i.e., they are independent. Metrics were aggregated within one sample of the validation fold, if applicable (i.e., all frames of one video). This process was repeated for all CV folds and per-sample metrics/metric aggregates were aggregated over all samples across all CV folds)
   - Over sample-level averages of n cross-validation folds (i.e., models were cross-validated on CV folds split along patient/video lines, i.e., they are independent. Metrics were aggregated within one sample of the validation fold, if applicable (i.e., all frames of one video). Then, per-sample metrics/metric aggregates were aggregated for each CV fold. The per-fold aggregates of n CV folds were aggregated for the final reported metric.)
   - Over n frames in test set (i.e., final models were tested on test set. Metrics were calculated per frame and there were multiple frames per sample/patient/video in the test set, which were not aggregated. Per-frame metrics were aggregated over all frames in the test set, without consideration of "patient lines")
   - Over n frames across all cross-validation folds (i.e., models were cross-validated on CV folds. Metrics were calculated per frame and there were multiple frames per sample/patient/video in the test set, which were not aggregated. This process was repeated for all CV folds and per-frame metrics were aggregated over all individual frames across all CV folds)
   - Over frame-level averages of n cross-validation folds (i.e., models were cross-validated on CV folds. Metrics were calculated per frame and there were multiple frames per sample/patient/video in the test set, which were not aggregated. Per-frame metrics were aggregated for each CV fold. The per-fold aggregates of n CV folds were aggregated for the final reported metric.)
   - Other
   - Unclear
f. Other statistical tests such as comparisons of model performance or with human expert annotations using frequentist parametric (e.g., t-test) or non-parametric methods.

#### **Supplementary Table 5**: Studies and study characteristics included in the systematic review and meta-analysis (3)

The following table presents an overview of all included studies, detailing key characteristics across four main categories: **Performance Metrics**. If the values were reported as average metrics for the most commonly used metrics in our review (not per every segment).

| First author, year [reference] | IoU | DSC | Precision | Recall | mAP | Accuracy |
|---|---|---|---|---|---|---|
| Acharya, 2022 [1] | n/a | n/a | n/a | n/a | n/a | 70/30 Hold-out: 96.09, 10-fold CV: 97.80 |
| Al Hajj, 2018 [2] | n/a | n/a | n/a | n/a | 97.89 | n/a |
| Alapatt, 2021 [3] | 61.58 | n/a | n/a | n/a | n/a | 95.22 |
| Ali, 2022 [4] | n/a | n/a | n/a | n/a | 42.64 (10% labelled data) | n/a |
| Alkhamaiseh, 2023 [5] | 74.65 | n/a | 93.9 | n/a | n/a | 92 |
| Alshirbaji, 2021 [6] | n/a | n/a | n/a | n/a | 94.95 | n/a |
| Alshirbaji, 2021 [7] | n/a | n/a | n/a | n/a | n/a | n/a |
| Alshirbaji, 2018 [8] | n/a | n/a | 73.93 | 63.24 | n/a | 93.65 |
| Ángeles-Cerón, 2021 [9] | n/a | 31.3 | n/a | n/a | n/a | n/a |
| Ángeles-Cerón, 2022 [10] | n/a | 48.9 | n/a | n/a | n/a | n/a |
| Aoyama, 2024 [11] | n/a | n/a | n/a | n/a | n/a | n/a |
| Arabian, 2022 [12] | n/a | n/a | n/a | n/a | 88.4 | n/a |
| Arabian, 2023 [13] | n/a | n/a | n/a | n/a | 93.14 | n/a |
| Aspart, 2022 [14] | n/a | n/a | n/a | n/a | n/a | n/a |
| Attia, 2017 [15] | 82.7 | n/a | n/a | n/a | n/a | 93.3 |
| Ayobi, 2024 [16] | 87.05 | n/a | n/a | n/a | n/a | n/a |
| Bai, 2023 [17] | n/a | n/a | n/a | n/a | n/a | n/a |
| Bakker, 2024 [18] | n/a | DSAD: 39.7 (All classes), Private: 73.5 (Catheter) and 75.5 (Urethra) | n/a | n/a | n/a | n/a |
| Bamba, 2021 [19] | n/a | n/a | 93.4 | 91.3 | n/a | n/a |
| Bamba, 2021 [20] | n/a | n/a | grasping forceps: 98, ultrasonic scalpel: 93.9, clip forceps: 92.7, angled forceps: 100, spatula forceps: 94.5 | grasping forceps: 98.1, ultrasonic scalpel: 99.4, clip forceps: 96.2, angled forceps: 94.9, spatula forceps: 98.1 | n/a | n/a |
| Ban, 2023 [21] | n/a | n/a | n/a | n/a | n/a | CVS100: 67.1, PGS200: 67.5, CholecT45: 30.6 (all triplets) |
| Batić, 2024 [22] | Semantic: 65.32 | n/a | n/a | n/a | Action Triplet: 30.17 | n/a |
| Batić, 2023 [23] | n/a | n/a | n/a | n/a | Action Triplet: 30.17 | n/a |
| Boonkong, 2022 [24] | n/a | n/a | 96.18 | 90.9 | n/a | n/a |
| Brandenburg, 2023 [25] | n/a | n/a | n/a | n/a | n/a | n/a |
| Casella, 2021 [26] | n/a | 71.76 | 89.76 | 50.66 | n/a | n/a |
| Chen, 2024 [27] | n/a | n/a | n/a | n/a | n/a | n/a |
| Chen, 2013 [28] | n/a | n/a | n/a | n/a | n/a | n/a |
| Chen, 2017 [29] | n/a | n/a | n/a | n/a | n/a | 93.2 |
| Choi, 2017 [30] | n/a | n/a | n/a | n/a | 72.26 | n/a |
| Ciaparrone, 2020 [31] | n/a | n/a | 87 | n/a | n/a | n/a |
| Colleoni, 2024 [32] | LC: 84.5, Robotic Partial Nephrectomy: 66.35, Robotic Prostatectomy: 47.88 | n/a | n/a | n/a | n/a | n/a |
| Colleoni, 2022 [33] | Tool Head: 38.99, Tool Shaft: 56.95 | n/a | n/a | n/a | n/a | n/a |
| Daneshgar Rahbar, 2023 [34] | n/a | n/a | n/a | n/a | n/a | n/a |
| Davila, 2023 [35] | n/a | n/a | n/a | n/a | n/a | n/a |
| De Backer, 2023 [36] | 94.4 | n/a | n/a | n/a | n/a | n/a |
| den Boer, 2023 [37] | n/a | Azygos vein: 79, Aorta: 74, Lung: 89 | n/a | n/a | n/a | n/a |
| Derathé, 2020 [38] | n/a | n/a | n/a | n/a | n/a | 68 |
| Du, 2019 [39] | n/a | n/a | n/a | n/a | n/a | n/a |
| Endo, 2023 [40] | n/a | n/a | n/a | n/a | n/a | n/a |
| Fernandez-Rodríguez, 2024 [41] | n/a | 63.14 | 66.5 | 71.43 | n/a | n/a |
| Fuentes-Hurtado, 2019 [42] | EndoVis15: 75.43, Sleeve Gastrectomy: 59 | n/a | n/a | n/a | n/a | n/a |
| Fujinaga, 2023 [43] | n/a | Common Bile Duct: 45, Cystic Duct: 20, S4 Segment: 27, Rouvier Sulcus: 30 | n/a | n/a | n/a | n/a |
| Ghamsarian, 2024 [44] | Endometriosis: 54.04, Colorectal: 60.01 | Endometriosis: 66.37, Colorectal: 73.09 | n/a | n/a | n/a | n/a |
| Gitau, 2024 [45] | n/a | n/a | n/a | n/a | n/a | n/a |
| Grammatikopoulou, 2023 [46] | LC: 69.38, PN: 62.91 | n/a | n/a | n/a | n/a | n/a |
| Guédon, 2021 [47] | n/a | n/a | LC: 89, LH: 88 | LC: 81, LH: 79 | n/a | n/a |
| Hasan, 2021 [48] | n/a | 89.56 | n/a | n/a | n/a | n/a |
| Huang, 2022 [49] | 74.92 | 85.63 | n/a | n/a | n/a | n/a |
| Jalal, 2022 [50] | n/a | n/a | n/a | n/a | n/a | n/a |
| Jalal, 2023 [51] | n/a | n/a | n/a | n/a | 95.6 | n/a |

| | | | | | | |
|---|---|---|---|---|---|---|
| Jamal, 2024 [52] | RARP: 85.8, LH: 78 | n/a | n/a | n/a | n/a | n/a |
| Jang, 2023 [53] | n/a | n/a | n/a | n/a | n/a | n/a |
| Jaspers, 2024 [54] | n/a | n/a | n/a | n/a | n/a | n/a |
| Jearanai, 2023 [55] | n/a | n/a | n/a | n/a | n/a | n/a |
| Jha, 2021 [56] | 81.83 | 87.39 | 93.01 | 88.99 | n/a | 98.98 |
| Jin, 2020 [57] | n/a | n/a | n/a | n/a | 89.2 | n/a |
| Jin, 2018 [58] | n/a | n/a | n/a | n/a | 63.1 | n/a |
| Kamrul Hasan, 2021 [59] | n/a | n/a | n/a | n/a | n/a | n/a |
| Kanakatte, 2020 [60] | n/a | n/a | n/a | n/a | 82 | n/a |
| Kawamura, 2023 [61] | n/a | n/a | 97.1 | 73.7 | n/a | 83.4 |
| Khalid, 2023 [62] | n/a | n/a | n/a | n/a | n/a | n/a |
| Khalid, 2023 [63] | n/a | n/a | n/a | n/a | n/a | n/a |
| Kim, 2024 [64] | n/a | n/a | n/a | n/a | n/a | n/a |
| Kinoshita, 2024 [65] | 29.2 | 44.2 | n/a | n/a | n/a | n/a |
| Kitaguchi, 2023 [66] | n/a | Ureter: 72.2, Hypogastric Nerves: 57.9, Aortic Plexus: 62.8 | Ureter: 70.7, Hypogastric Nerves: 59.2, Aortic Plexus: 65.6 | Ureter: 73.9, Hypogastric Nerves: 56.6, Aortic Plexus: 60.3 | n/a | n/a |
| Kitaguchi, 2022 [67] | n/a | n/a | n/a | n/a | 91.9 | n/a |
| Kitaguchi, 2022 [68] | n/a | n/a | n/a | n/a | n/a | n/a |
| Kletz, 2019 [69] | n/a | n/a | n/a | n/a | n/a | n/a |
| Kolbinger, 2023 [70] | n/a | n/a | n/a | n/a | n/a | n/a |
| Kolbinger, 2024 [71] | n/a | n/a | n/a | n/a | n/a | n/a |
| Kolbinger, 2024 [72] | n/a | n/a | n/a | n/a | n/a | n/a |
| Kondo, 2021 [73] | n/a | n/a | 80.2 | 68 | n/a | n/a |
| Konduri, 2024 [74] | n/a | n/a | n/a | n/a | n/a | LapGyn4: 99.74, Cholec80: 99.45, Heichole: 99.34 |
| Kong, 2021 [75] | 40.46 | 48.42 | 71.34 | 49.03 | n/a | n/a |
| Kumazu, 2021 [76] | n/a | 54.9 | n/a | 60.6 | n/a | n/a |
| Kumazu, 2025 [77] | n/a | 46 | n/a | 53 | n/a | n/a |
| Labrunie, 2022 [78] | n/a | n/a | n/a | n/a | n/a | n/a |
| Lam, 2022 [79] | n/a | n/a | n/a | n/a | 83.9 | n/a |
| Laplante, 2022 [80] | n/a | n/a | n/a | n/a | n/a | Go: 92, No-Go: 92 |
| Lavanchy, 2021 [81] | n/a | n/a | n/a | n/a | n/a | 87 |
| Le, 2023 [82] | n/a | n/a | 96.9 | 93.7 | n/a | n/a |
| Lee, 2024 [83] | n/a | n/a | n/a | n/a | n/a | n/a |
| Leifman, 2024 [84] | n/a | n/a | n/a | n/a | n/a | n/a |
| Leifmann, 2022 [85] | n/a | n/a | n/a | n/a | n/a | n/a |
| Li, 2025 [86] | n/a | n/a | n/a | n/a | n/a | n/a |
| Li, 2023 [87] | n/a | n/a | n/a | n/a | n/a | CVS: 92 |
| Liao, 2024 [88] | n/a | n/a | n/a | n/a | n/a | n/a |
| Lin, 2024 [89] | n/a | n/a | n/a | n/a | n/a | n/a |
| Lin, 2021 [90] | 73.5 | 81 | n/a | n/a | n/a | n/a |
| Liu, 2022 [91] | n/a | n/a | n/a | n/a | 71 | n/a |
| Liu, 2024 [92] | 48.2 | n/a | n/a | n/a | n/a | n/a |
| Liu, 2022 [93] | n/a | n/a | n/a | n/a | n/a | n/a |
| Lou, 2023 [94] | n/a | n/a | n/a | n/a | n/a | n/a |
| Loukas, 2020 [95] | n/a | n/a | n/a | n/a | n/a | n/a |
| Loza, 2024 [96] | n/a | n/a | n/a | n/a | n/a | n/a |
| Maack, 2024 [97] | n/a | n/a | n/a | n/a | n/a | n/a |
| Madad Zadeh, 2020 [98] | Uterus: 84.5, Ovaries: 29.6, Tools: 54.5 | n/a | n/a | n/a | n/a | n/a |
| Madani, 2022 [99] | Go: 53, No-Go: 71, Liver: 86, Gallbladder: 72, Triangle: 65 | n/a | n/a | n/a | n/a | n/a |
| Maqbool, 2020 [100] | 46 | n/a | 66 | 55 | n/a | n/a |
| Marullo, 2023 [101] | n/a | 81.89 | n/a | n/a | n/a | 90.63 |
| Mascagni, 2022 [102] | n/a | n/a | n/a | n/a | n/a | n/a |
| Matsumoto, 2024 [103] | n/a | 83.7 | n/a | n/a | n/a | n/a |
| Mehta, 2024 [104] | n/a | 71.5 | n/a | n/a | n/a | n/a |
| Mishra, 2017 [105] | n/a | n/a | n/a | n/a | n/a | 88.75 |
| Murali, 2024 [106] | 45.02 | 57.6 | n/a | n/a | n/a | n/a |
| Murali, 2023 [107] | n/a | n/a | n/a | n/a | 69.7 | n/a |
| Murali, 2022 [108] | n/a | n/a | n/a | n/a | n/a | n/a |
| Myo, 2024 [109] | n/a | n/a | n/a | n/a | n/a | n/a |
| Nakanuma, 2023 [110] | n/a | n/a | n/a | n/a | n/a | n/a |
| Namazi, 2022 [111] | n/a | n/a | n/a | n/a | n/a | m2cai: 81.84, Cholec80: 91.92 |
| Nema , 2023 [112] | 80.89 | n/a | n/a | n/a | n/a | n/a |
| Nwoye, 2024 [113] | n/a | n/a | n/a | n/a | n/a | n/a |
| Nwoye, 2019 [114] | n/a | n/a | n/a | n/a | n/a | n/a |
| Oh, 2024 [115] | n/a | BD: 72.8, AW: 42.9 | n/a | n/a | n/a | n/a |
| Owen, 2022 [116] | 68.1 | n/a | 86.7 | 76.1 | n/a | n/a |
| Ozbulak, 2025 [117] | n/a | n/a | n/a | n/a | n/a | n/a |
| Pan, 2023 [118] | n/a | n/a | n/a | n/a | m2cai: 96.8, LGIL: 95.6 | n/a |
| Penza, 2018 [119] | n/a | n/a | n/a | n/a | n/a | n/a |
| Pradeep, 2022 [120] | n/a | n/a | n/a | n/a | 97.51 | n/a |
| Prokopetc, 2015 [121] | n/a | n/a | n/a | Left FU: 73, Right FU: 64 | n/a | n/a |

| | | | | | | |
|---|---|---|---|---|---|---|
| Protserov, 2024 [122] | n/a | n/a | n/a | n/a | n/a | n/a |
| Rahbar, 2020 [123] | n/a | n/a | n/a | n/a | n/a | n/a |
| Rahbar, 2024 [124] | n/a | n/a | n/a | n/a | n/a | 85.5 |
| Raja, 2024 [125] | n/a | n/a | n/a | n/a | n/a | n/a |
| Ryu, 2024 [126] | n/a | n/a | n/a | n/a | n/a | n/a |
| Sahu, 2017 [127] | n/a | n/a | n/a | n/a | 65 | n/a |
| Samuel, 2021 [128] | n/a | n/a | 91.5 | 78.4 | n/a | n/a |
| Sanchez-Matilla, 2022 [129] | 38.4 | 49.33 | n/a | n/a | 72.67 | n/a |
| Satyanaik, 2024 [130] | n/a | n/a | n/a | n/a | n/a | n/a |
| Satyanaik, 2024 [131] | n/a | n/a | n/a | n/a | n/a | n/a |
| Seenivasan, 2023 [132] | n/a | n/a | n/a | n/a | n/a | n/a |
| Sengun, 2023 [133] | 66 | 77 | n/a | n/a | n/a | n/a |
| Sengun, 2024 [134] | 63.7 | 77.1 | n/a | n/a | n/a | n/a |
| Shen, 2023 [135] | 63.37 | 75.48 | n/a | n/a | n/a | n/a |
| Sheng, 2024 [136] | 46.31 | n/a | n/a | n/a | n/a | n/a |
| Shi, 2020 [137] | n/a | n/a | n/a | n/a | 91.65 | n/a |
| Shimgekar, 2021 [138] | n/a | n/a | n/a | n/a | n/a | n/a |
| Silva, 2022 [139] | 55 | n/a | 73 | 72 | n/a | n/a |
| Smithmaitrie, 2024 [140] | n/a | n/a | 89.8 | 91.7 | 95.8 | n/a |
| Sonsilphong, 2022 [141] | n/a | n/a | n/a | n/a | n/a | n/a |
| Streckert, 2023 [142] | 76.15 | n/a | n/a | n/a | n/a | n/a |
| Strong, 2024 [143] | 82 | n/a | n/a | n/a | n/a | 87.2 |
| Sun, 2022 [144] | n/a | n/a | n/a | n/a | n/a | n/a |
| Takeuchi, 2023 [145] | n/a | n/a | n/a | n/a | n/a | 77.1 |
| Tao, 2023 [146] | n/a | n/a | n/a | n/a | Cholec80: 95.15, m2cai: 93.4 | n/a |
| Tokuyasu, 2021 [147] | n/a | n/a | Common bile duct: 32, Cystic duct: 74, Lower edge of the left medial liver segment: 31.4, Rouviere's sulcus: 10.1 | n/a | n/a | n/a |
| Urrea, 2024 [148] | 97.7 | n/a | n/a | n/a | n/a | 97.6 |
| Valderrama, 2022 [149] | n/a | n/a | n/a | n/a | n/a | n/a |
| Vardazaryan, 2018 [150] | n/a | n/a | n/a | n/a | 87.4 | n/a |
| Wagner, 2023 [151] | n/a | n/a | n/a | n/a | n/a | n/a |
| Wang, 2023 [152] | n/a | n/a | 97.4 | 98.1 | n/a | n/a |
| Wang, 2017 [153] | n/a | n/a | n/a | n/a | n/a | n/a |
| Wang, 2024 [154] | n/a | n/a | n/a | n/a | n/a | n/a |
| Wang, 2023 [155] | n/a | n/a | n/a | n/a | n/a | n/a |
| Wang, 2025 [156] | n/a | GRASP: 88.72 | n/a | n/a | n/a | n/a |
| Wang, 2021 [157] | n/a | n/a | n/a | n/a | 87.6 | n/a |
| Wang, 2020 [158] | n/a | n/a | n/a | n/a | m2cai: 70.1, AJU: 77.3 | n/a |
| Wang, 2024 [159] | n/a | n/a | n/a | n/a | n/a | n/a |
| Wang, 2023 [160] | n/a | n/a | 97.8 | n/a | n/a | n/a |
| Ward, 2022 [161] | n/a | n/a | n/a | n/a | n/a | n/a |
| Wei, 2024 [162] | Robust-Mis: 86.6, CholecSeg8k: 70.2 | Robust-Mis: 92.9, CholecSeg8k: 86.5 | n/a | n/a | n/a | n/a |
| Wei, 2023 [163] | n/a | n/a | n/a | n/a | n/a | n/a |
| Wu, 2024 [164] | n/a | 92.33 | n/a | n/a | n/a | n/a |
| Xi, 2022 [165] | n/a | n/a | 67.01 | n/a | n/a | n/a |
| Xu, 2024 [166] | 80.26 | 87.37 | n/a | n/a | n/a | n/a |
| Xue, 2022 [167] | n/a | n/a | n/a | n/a | 85.43 | n/a |
| Yamazaki, 2020 [168] | n/a | n/a | 87 | 83 | n/a | n/a |
| Yamlahi, 2023 [169] | n/a | n/a | n/a | n/a | 37.4 | 74 |
| Yang, 2024 [170] | n/a | n/a | n/a | n/a | 69.34 | n/a |
| Yang, 2023 [171] | n/a | n/a | n/a | n/a | n/a | n/a |
| Yang, 2021 [172] | n/a | n/a | n/a | n/a | n/a | n/a |
| Yin, 2024 [173] | n/a | n/a | n/a | n/a | n/a | n/a |
| Yuan, 2024 [174] | n/a | n/a | n/a | n/a | n/a | n/a |
| Zhang, 2024 [175] | n/a | n/a | n/a | 10.59 | n/a | 77.1 |
| Zhang, 2023 [176] | n/a | n/a | n/a | 95 | 95 | n/a |
| Zhang, 2023 [177] | n/a | n/a | n/a | n/a | 96.23 | n/a |
| Zhang, 2020 [178] | n/a | n/a | n/a | n/a | m2cai: 69.6, AJU: 76.5 | n/a |
| Zhang, 2023 [179] | n/a | n/a | n/a | n/a | n/a | n/a |
| Zhang, 2020 [180] | n/a | n/a | n/a | n/a | n/a | n/a |
| Zhao, 2021 [181] | 80.1 | 89.5 | n/a | n/a | n/a | n/a |
| Zhao, 2023 [182] | n/a | n/a | 98.9 | n/a | n/a | n/a |
| Zhao, 2021 [183] | 77.7 | 86.2 | n/a | n/a | n/a | n/a |
| Zhao, 2019 [184] | n/a | n/a | n/a | n/a | n/a | n/a |
| Zheng, 2023 [185] | 82.8 | n/a | 91.35 | n/a | n/a | n/a |
| Zhou, 2024 [186] | 72.6 | 84.1 | n/a | n/a | n/a | n/a |
| Zhou, 2022 [187] | n/a | n/a | 96.3 | 92.3 | n/a | n/a |
| Zygomalas, 2024 [188] | n/a | n/a | n/a | n/a | n/a | n/a |

*n/a: not applicable*

#### **Supplementary Figure 1**: Overview of Findings by Category

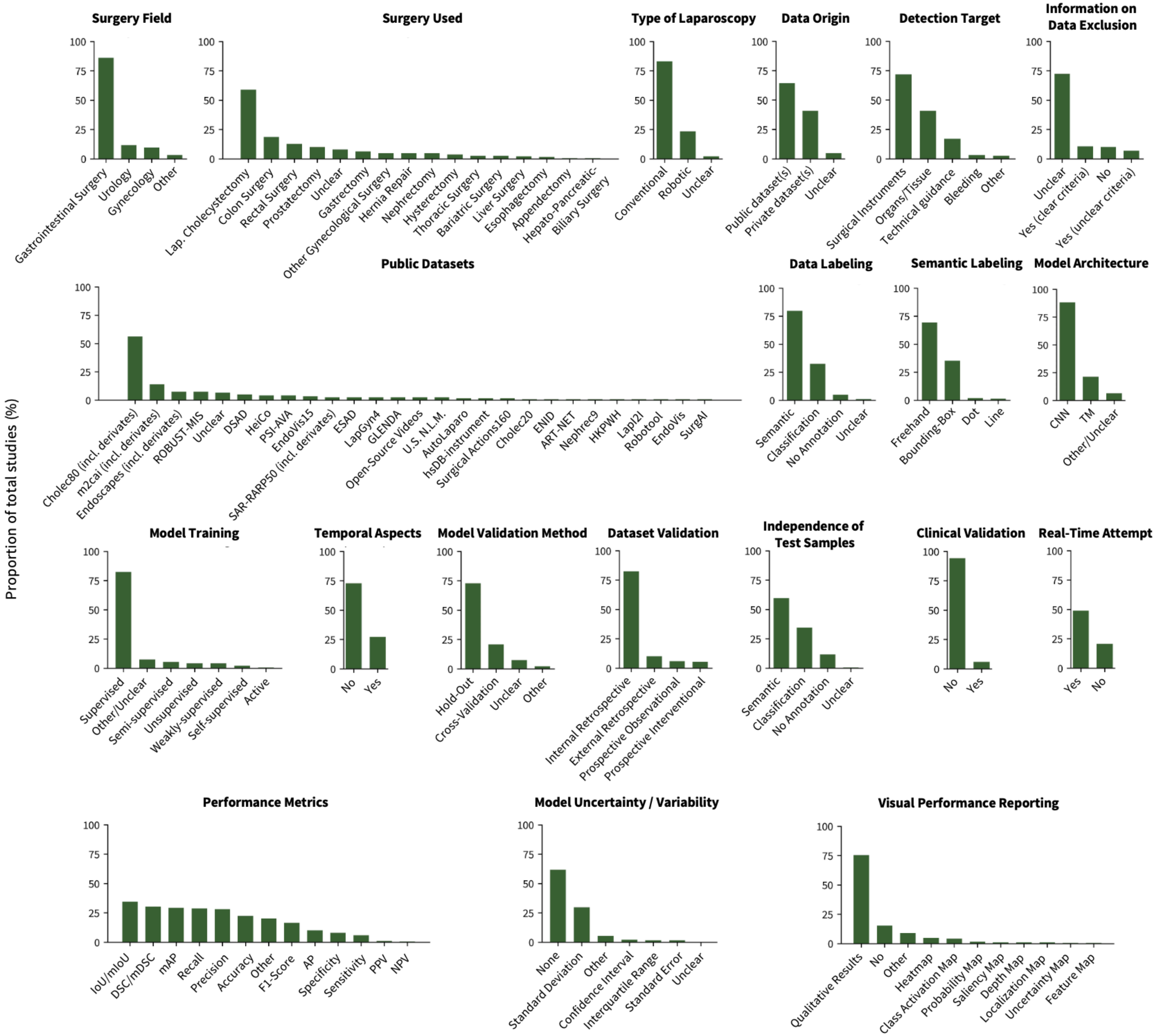

Bar charts display the proportion of individual data points within specific categories, based on all 188 included publications.

#### **Supplementary Table 6**: Clinical Translation

All 11 studies that tested the developed AI application in real-life scenarios on patients.

| First author, year [reference] | Surgery | Number of patients | Type of AI implementation in the OR |
|---|---|---|---|
| Nakanuma, 2023 [110] | Laparoscopic Cholecystectomy | 10 | AI-based landmark detection and its performance evaluation clinically using a novel expert-based assessment method to verify its feasibility and accuracy in real surgical settings. |
| Fujinaga, 2023 [43] | Laparoscopic Cholecystectomy | 10 | A cross-AI system combining landmark detection and surgical phase recognition was developed to support surgeons in identifying key anatomical structures, potentially helping to prevent bile duct injury. |
| Smithmaitrie, 2024 [140] | Laparoscopic Cholecystectomy | unclear | AI application for detecting key anatomical landmarks and generating a real-time guided dissection line, supporting safer and more precise surgical navigation to help prevent bile duct injury. |
| Tokuyasu, 2021 [147] | Laparoscopic Cholecystectomy | 1 | AI–based system was developed to detect four key anatomical landmarks during laparoscopic cholecystectomy, aiming to raise intraoperative awareness and reduce the risk of bile duct injury. |
| Leifman, 2024 [84] | Laparoscopic Cholecystectomy | 40 | AI-based assessment of the achievement of the Critical View of Safety, demonstrating high sensitivity and specificity, for improving surgical safety and outcomes. |
| Brandenburg, 2023 [25] | Robotic Esophagectomy | unclear | AI-based recognition of key surgical features related to bleeding from intraoperative videos, aiming to personalize surgical outcome predictions. |
| Aoyama, 2024 [11] | Laparoscopic Gastrectomy | 10 | AI-based real-time navigation system that identifies specific anatomical landmarks linked to postoperative pancreatic fistula, helping surgeons reduce complication risks by highlighting critical structures during surgery. |
| Kitaguchi, 2023 [66] | Laparoscopic Left-sided colorectal resection | 20 | AI-based real-time recognition of critical anatomical structures, specifically the ureter and autonomic nerves, achieving faster and accurate identification compared to surgeons as a decision-support tool. |
| Zheng, 2023 [185] | Laparoscopic Partial nephrectomy | 12 | AI-based real-time system that evaluates surgeons' laparoscope control during surgery, helping non-experts reaching higher expert-level performance in visual field management. |
| De Backer, 2023 [36] | Different robotic kidney surgeries | 10 | AI-based detecting surgical instruments in real time during AR-guided robotic kidney surgeries, aiming to enhance safety by preventing hazardous overlay issues and enabling more reliable augmented reality integration in clinical settings. |
| Jearanai, 2023 [55] | Different laparoscopic surgeries | Unclear | AI model with a real-time alarm system to accurately detect abdominal wall layers during trocar insertion, aiming to enhance surgical safety by reducing complications through timely anatomical recognition and feedback. |

### **Supplementary Table 7**: Reporting Quality

All reporting quality ratings by category and publication. The following coding is used:

- Reported = 3
- Partially Reported = 2
- Reference to another publication = 1
- Not Reported = 0
- Not applicable = NA

Each cell shows three values (e.g., 3/2/2), representing the ratings from Annotator 1, Annotator 2, and the final consensus rating by Annotator 3, which was used for statistical analysis. Row 1 shows the interrater reliability between Annotator 1 and Annotator 2, assessed using Cohen's Kappa for each of the 18 reporting quality categories.

| | Clinical Rationale | | | | | Data | | | | Model Training and Validation | | | | | | Critical appraisal | | Ethics and Reproducibility | |
|---|---|---|---|---|---|---|---|---|---|---|---|---|---|---|---|---|---|---|---|
| **First author, year [reference]** | **Study Design** | **Prediction Problem** | **Clinical Problem** | **Clinical Goal** | **Existing AI** | **Data Sources** | **Data Selection** | **Data Preprocessing** | **Labeling** | **Model Type** | **Model Development** | **Model Validation** | **Model Performance** | **Statistical Methods** | **Performance Errors** | **Clinical Implications** | **Limitations** | **Ethical Statement** | **Code Publication** |
| **Cohen's Kappa Value** | **0.854** | **0.946** | **0.907** | **0.843** | **0.725** | **0.94** | **0.95** | **0.884** | **0.947** | **1** | **0.854** | **0.814** | **0.883** | **0.821** | **0.932** | **0.939** | **0.938** | **0.975** | **0.954** |
| Acharya, 2022 [1] | 3/3/3 | 2/2/2 | 0/0/0 | 0/0/0 | 2/2/2 | 1/1/1 | 0/NA/0 | 1/1/1 | 1/1/1 | 3/3/3 | 2/2/2 | 2/2/2 | 2/2/2 | 2/2/2 | 2/3/2 | 0/0/0 | 0/0/0 | NA/NA/NA | 0/0/0 |
| Al Hajj, 2018 [2] | 3/3/3 | 3/3/3 | 0/0/0 | 0/0/0 | 3/3/3 | 1/1/1 | 0/0/0 | 2/2/2 | 1/1/1 | 3/3/3 | 3/3/3 | 2/2/2 | 3/3/3 | 3/3/3 | 3/3/3 | 0/0/0 | 2/2/2 | NA/NA/NA | 2/2/2 |
| Alapatt, 2021 [3] | 3/3/3 | 2/2/2 | 0/0/0 | 0/0/0 | 2/2/2 | 2/2/2 | 0/0/0 | 2/2/2 | 0/0/0 | 3/3/3 | 2/2/2 | 2/2/2 | 2/2/2 | 2/2/2 | 0/0/0 | 0/0/0 | 0/0/0 | 3/3/3 | 0/0/0 |
| Ali, 2022 [4] | 3/3/3 | 3/3/3 | 2/2/2 | 2/2/2 | 3/3/3 | 1/1/1 | 1/1/1 | 2/2/2 | 1/1/1 | 3/3/3 | 3/3/3 | 3/3/3 | 3/3/3 | 3/3/3 | 2/2/2 | 0/0/0 | 0/0/0 | NA/NA/NA | 3/3/3 |
| Alkhamaiseh, 2023 [5] | 3/3/3 | 3/3/3 | 3/3/3 | 3/3/3 | 2/2/2 | 2/2/2 | 2/2/2 | 2/2/2 | 3/3/3 | 3/3/3 | 2/2/2 | 3/3/3 | 2/2/2 | 2/2/2 | 0/0/0 | 3/3/3 | 3/3/3 | 0/0/0 | 0/0/0 |
| Alshirbaji, 2021 [6] | 3/3/3 | 3/3/3 | 0/0/0 | 0/0/0 | 3/3/3 | 2/2/2 | 1/1/1 | 2/2/2 | 1/1/1 | 3/3/3 | 3/3/3 | 3/3/3 | 2/2/2 | 2/2/2 | 2/2/2 | 0/0/0 | 2/2/2 | NA/NA/NA | 0/0/0 |
| Alshirbaji, 2021 [7] | 3/3/3 | 2/2/2 | 0/0/0 | 0/0/0 | 2/3/2 | 1/1/1 | 0/0/0 | 0/0/0 | 1/1/1 | 3/3/3 | 0/0/0 | 2/2/2 | 2/2/2 | 0/0/0 | 0/0/0 | 0/0/0 | 2/2/2 | 3/3/3 | 0/0/0 |
| Alshirbaji, 2018 [8] | 3/3/3 | 2/2/2 | 0/2/0 | 0/2/0 | 2/3/2 | 1/1/1 | 0/0/0 | 0/0/0 | 1/1/1 | 3/3/3 | 2/2/2 | 2/3/2 | 2/2/2 | 0/0/0 | 0/0/0 | 0/0/0 | 0/2/0 | NA/NA/NA | 0/0/0 |
| Ángeles-Cerón, 2021 [9] | 3/3/3 | 2/2/2 | 0/0/0 | 0/0/0 | 2/2/2 | 1/1/1 | 2/2/2 | 2/3/3 | 1/1/1 | 3/3/3 | 2/2/2 | 2/2/2 | 2/2/2 | 2/2/2 | 0/0/0 | 0/0/0 | 2/2/2 | NA/NA/NA | 0/0/0 |
| Ángeles-Cerón, 2022 [10] | 3/3/3 | 3/3/3 | 2/2/2 | 2/2/2 | 2/2/2 | 1/1/1 | 2/2/2 | 3/3/3 | 1/1/1 | 3/3/3 | 3/3/3 | 3/3/3 | 3/3/3 | 2/2/2 | 0/0/0 | 2/2/2 | 0/0/0 | NA/NA/NA | 0/0/0 |
| Aoyama, 2024 [11] | 3/3/3 | 3/3/3 | 3/3/3 | 3/3/3 | 2/3/2 | 3/3/3 | 2/2/2 | 2/2/2 | 3/3/3 | 3/3/3 | 0/0/0 | 2/2/2 | 2/2/2 | 2/2/2 | 2/2/2 | 3/3/3 | 3/3/3 | 3/3/3 | 0/0/0 |
| Arabian, 2022 [12] | 3/3/3 | 3/3/3 | 0/0/0 | 0/0/0 | 2/2/2 | 1/1/1 | 0/0/0 | 0/0/0 | 1/1/1 | 3/3/3 | 2/3/2 | 2/2/2 | 2/2/2 | 0/0/0 | 0/0/0 | 0/0/0 | 0/0/0 | NA/NA/NA | 0/0/0 |
| Arabian, 2023 [13] | 3/3/3 | 3/3/3 | 2/2/2 | 2/2/2 | 3/3/3 | 1/1/1 | 0/0/0 | 2/2/2 | 1/1/1 | 3/3/3 | 2/2/2 | 3/3/3 | 2/2/2 | 2/2/2 | 0/0/0 | 0/0/0 | 0/0/0 | NA/NA/NA | 0/0/0 |
| Aspart, 2022 [14] | 3/3/3 | 3/3/3 | 3/3/3 | 3/3/3 | 2/2/2 | 2/2/2 | 2/2/2 | 2/2/2 | 3/3/3 | 3/3/3 | 3/3/3 | 3/3/3 | 3/3/3 | 2/3/2 | 2/2/2 | 3/3/3 | 2/2/2 | 3/3/3 | 0/0/0 |
| Attia, 2017 [15] | 3/3/3 | 2/2/2 | 0/0/0 | 0/2/0 | 2/2/2 | 1/1/1 | 0/0/0 | 2/2/2 | 1/1/1 | 3/3/3 | 2/3/2 | 1/1/1 | 2/2/2 | 0/3/2 | 0/2/2 | 0/2/0 | 0/3/0 | NA/NA/NA | 0/0/0 |
| Ayobi, 2024 [16] | 3/3/3 | 2/2/2 | 0/0/0 | 0/0/0 | 3/3/3 | 2/2/2 | 0/0/0 | 2/2/2 | 3/3/3 | 3/3/3 | 3/3/3 | 2/2/2 | 3/3/3 | 2/2/2 | 2/2/2 | 0/0/0 | 0/0/0 | 2/2/2 | 3/3/3 |
| Bai, 2023 [17] | 3/3/3 | 3/3/3 | 0/0/0 | 0/0/0 | 2/3/2 | 1/1/1 | 0/0/0 | 0/0/0 | 1/1/1 | 3/3/3 | 2/2/2 | 2/2/2 | 3/3/3 | 2/2/2 | 0/0/0 | 0/0/0 | 0/0/0 | NA/NA/NA | 3/3/3 |
| Bakker, 2024 [18] | 3/3/3 | 3/3/3 | 3/3/3 | 3/3/3 | 2/2/2 | 2/2/2 | 2/2/2 | 2/2/2 | 3/3/3 | 3/3/3 | 2/2/2 | 2/2/2 | 2/2/2 | 2/2/2 | 2/2/2 | 2/2/2 | 3/3/3 | 3/3/3 | 0/0/0 |
| Bamba, 2021 [19] | 3/3/3 | 2/3/3 | 0/0/0 | 0/0/0 | 2/2/2 | 2/2/2 | 0/0/0 | 0/0/0 | 0/0/0 | 3/3/3 | 0/0/0 | 2/2/2 | 2/2/2 | 0/0/0 | 0/0/0 | 2/2/2 | 3/3/3 | 3/3/3 | 0/0/0 |
| Bamba, 2021 [20] | 3/3/3 | 3/3/3 | 2/2/2 | 2/2/2 | 3/3/3 | 2/3/3 | 2/2/2 | 2/3/2 | 2/2/2 | 3/3/3 | 2/2/2 | 2/2/2 | 3/3/3 | 2/2/2 | 2/2/2 | 3/3/3 | 3/3/3 | 3/3/3 | 2/2/2 |
| Ban, 2023 [21] | 3/3/3 | 3/3/3 | 0/0/0 | 2/2/2 | 2/2/2 | 1/1/1 | 0/0/0 | 2/2/2 | 1/1/1 | 3/3/3 | 3/3/3 | 2/3/2 | 3/3/3 | 2/2/2 | 0/0/0 | 2/2/2 | 0/0/0 | 0/0/0 | 0/0/0 |
| Batić, 2024 [22] | 3/3/3 | 2/2/2 | 0/0/0 | 0/0/0 | 2/2/2 | 1/1/1 | 0/0/0 | 1/1/1 | 1/1/1 | 3/3/3 | 2/2/2 | 2/2/2 | 2/2/2 | 2/2/2 | 0/0/0 | 0/0/0 | 0/0/0 | NA/NA/NA | 3/3/3 |
| Batić, 2023 [23] | 3/3/3 | 2/2/2 | 0/0/0 | 0/0/0 | 2/2/2 | 1/1/1 | 0/0/0 | 2/2/2 | 0/1/0 | 3/3/3 | 0/0/0 | 2/2/2 | 2/2/2 | 0/0/0 | 2/2/2 | 0/0/0 | 2/2/2 | NA/NA/NA | 2/2/2 |
| Boonkong, 2022 [24] | 3/3/3 | 2/2/2 | 0/0/0 | 0/0/0 | 2/2/2 | 0/0/0 | 0/0/0 | 0/0/0 | 0/0/0 | 3/3/3 | 0/0/0 | 2/2/2 | 2/2/2 | 0/0/0 | 0/0/0 | 0/0/0 | 0/0/0 | 0/0/0 | 0/0/0 |
| Brandenburg, 2023 [25] | 3/3/3 | 3/3/3 | 2/2/2 | 2/2/2 | 2/2/2 | 3/3/3 | 3/3/3 | 2/2/2 | 3/3/3 | 3/3/3 | 2/2/2 | 2/2/2 | 3/3/3 | 3/3/3 | 3/3/3 | 3/3/3 | 3/3/3 | 3/3/3 | 3/3/3 |
| Casella, 2021 [26] | 3/3/3 | 3/3/3 | 2/2/2 | 2/3/3 | 2/2/2 | 1/1/1 | 2/2/2 | 3/3/3 | 1/1/1 | 3/3/3 | 2/2/2 | 2/2/2 | 2/2/2 | 2/2/2 | 2/2/2 | 2/2/2 | 3/3/3 | NA/NA/NA | 0/0/0 |
| Chen, 2024 [27] | 3/3/3 | 2/2/2 | 2/2/2 | 2/2/2 | 0/0/0 | 2/2/2 | 0/0/0 | 0/0/0 | NA/NA/NA | 3/3/3 | 2/2/2 | 2/2/2 | 2/2/2 | 2/2/2 | 0/0/0 | 2/2/2 | 0/0/0 | 0/0/0 | 0/0/0 |
| Chen, 2013 [28] | 3/3/3 | 2/2/2 | 0/0/0 | 0/0/0 | 2/2/2 | 0/0/0 | 0/0/0 | 0/0/0 | 0/0/0 | 3/3/3 | 2/2/2 | 0/0/0 | 2/2/2 | 0/0/0 | 0/0/0 | 0/0/0 | 0/0/0 | 0/0/0 | 0/0/0 |
| Chen, 2017 [29] | 3/3/3 | 3/3/3 | 2/2/2 | 2/2/2 | 2/2/2 | 2/2/2 | 0/0/0 | 3/3/3 | 2/2/2 | 3/3/3 | 3/3/3 | 2/2/2 | 2/2/2 | 2/2/2 | 3/3/3 | 0/0/0 | 0/0/0 | 3/3/3 | 3/3/3 |
| Choi, 2017 [30] | 3/3/3 | 2/2/2 | 0/0/0 | 0/0/0 | 2/2/2 | 1/1/1 | 0/0/0 | 2/2/2 | 0/0/0 | 3/3/3 | 2/3/2 | 2/2/2 | 2/2/2 | 0/0/0 | 3/3/3 | 0/2/0 | 2/2/2 | NA/NA/NA | 0/0/0 |
| Ciaparrone, 2020 [31] | 3/3/3 | 2/2/2 | 2/2/2 | 2/2/2 | 2/2/2 | 2/2/2 | 2/2/2 | 3/3/3 | 2/2/2 | 3/3/3 | 2/2/2 | 2/2/2 | 2/2/2 | 0/0/0 | 0/0/0 | 2/2/2 | 0/2/0 | 0/0/0 | 0/0/0 |
| Colleoni, 2024 [32] | 3/3/3 | 2/2/2 | 2/2/2 | 2/2/2 | 2/2/2 | 2/2/2 | 0/0/0 | 2/2/2 | 0/0/0 | 3/3/3 | 2/2/2 | 2/2/2 | 3/3/3 | 3/3/3 | 0/0/0 | 2/2/2 | 0/0/0 | 0/0/0 | 0/0/0 |
| Colleoni, 2022 [33] | 3/3/3 | 2/2/2 | 0/0/0 | 0/0/0 | 2/2/2 | 2/2/2 | 0/0/0 | 2/2/2 | 0/0/0 | 3/3/3 | 3/3/3 | 2/2/2 | 3/3/3 | 2/2/2 | 0/0/0 | 0/0/0 | 0/0/0 | 0/0/0 | 3/3/3 |
| Daneshgar Rahbar, 2023 [34] | 3/3/3 | 2/2/2 | 2/2/2 | 2/2/2 | 2/2/2 | 2/2/2 | 0/0/0 | 2/2/2 | 0/0/0 | 3/3/3 | 2/2/2 | 2/2/2 | 2/2/2 | 2/2/2 | 2/2/2 | 0/0/0 | 0/0/0 | NA/NA/NA | 3/3/3 |
| Davila, 2023 [35] | 3/3/3 | 2/2/2 | 0/0/0 | 0/0/0 | 2/2/2 | 1/1/1 | 0/0/0 | 2/2/2 | 1/0/1 | 3/3/3 | 2/2/2 | 2/2/2 | 2/2/2 | 0/0/0 | 0/0/0 | 0/0/0 | 0/0/0 | NA/NA/NA | 0/0/0 |
| De Backer, 2023 [36] | 3/3/3 | 3/3/3 | 3/3/3 | 3/3/3 | 2/2/2 | 2/2/2 | 0/0/0 | 2/2/2 | 2/2/2 | 2/2/2 | 0/2/2 | 2/0/2 | 2/2/2 | 2/2/2 | 0/0/0 | 3/3/3 | 2/2/2 | 0/0/0 | 0/0/0 |
| den Boer, 2023 [37] | 3/3/3 | 3/3/3 | 3/3/3 | 2/3/3 | 2/2/2 | 3/3/3 | 3/3/3 | 2/2/2 | 3/3/3 | 3/3/3 | 2/3/3 | 2/2/2 | 3/3/3 | 3/3/3 | 0/0/0 | 3/3/3 | 3/3/3 | 3/3/3 | 0/0/0 |
| Derathé, 2020 [38] | 3/3/3 | 3/3/3 | 2/2/2 | 2/2/2 | 2/2/2 | 2/2/2 | 0/0/0 | 2/2/2 | 3/3/3 | 3/3/3 | 2/2/2 | 2/3/3 | 2/2/2 | 0/0/0 | 0/0/0 | 2/2/2 | 0/0/0 | 3/3/3 | 0/0/0 |
| Du, 2019 [39] | 3/3/3 | 0/0/0 | 0/0/0 | 0/0/0 | 3/3/3 | 2/2/2 | 2/2/2 | 2/2/2 | 1/1/1 | 3/3/3 | 3/3/3 | 2/2/2 | 2/2/2 | 2/2/2 | 2/2/2 | 0/0/0 | 2/2/2 | NA/NA/NA | 3/3/3 |
| Endo, 2023 [40] | 3/3/3 | 3/3/3 | 2/2/2 | 2/2/2 | 2/2/2 | 2/2/2 | 2/2/2 | 2/2/2 | 2/2/2 | 3/3/3 | 2/2/2 | 2/2/2 | 0/0/0 | 0/0/0 | 0/0/0 | 3/3/3 | 3/3/3 | 3/3/3 | 0/0/0 |
| Fernandez-Rodríguez, 2024 [41] | 3/3/3 | 2/2/2 | 0/0/0 | 0/0/0 | 2/2/2 | 1/1/1 | 0/0/0 | 2/2/2 | 1/1/1 | 3/3/3 | 2/2/2 | 2/2/2 | 3/3/3 | 2/3/2 | 0/0/0 | 0/0/0 | 2/2/2 | NA/NA/NA | 0/0/0 |
| Fuentes-Hurtado, 2019 [42] | 3/3/3 | 3/3/3 | 2/2/2 | 0/0/0 | 2/2/2 | 2/2/2 | 0/0/0 | 2/2/2 | 2/2/2 | 3/3/3 | 3/3/3 | 2/2/2 | 2/2/2 | 2/2/2 | 0/0/0 | 0/0/0 | 2/2/2 | NA/NA/NA | 0/0/0 |
| Fujinaga, 2023 [43] | 3/3/3 | 3/3/3 | 3/3/3 | 3/3/3 | 3/3/3 | 3/3/3 | 2/2/2 | 2/2/2 | 2/2/2 | 3/3/3 | 2/2/2 | 2/2/2 | 2/2/2 | 2/2/2 | 0/0/0 | 3/3/3 | 3/3/3 | 3/3/3 | 0/0/0 |

| | | | | | | | | | | | | | | | | | | | |
|---|---|---|---|---|---|---|---|---|---|---|---|---|---|---|---|---|---|---|---|
| Ghamsarian, 2024 [44] | 3/3/3 | 2/2/2 | 0/0/0 | 0/0/0 | 0/0/0 | 2/2/2 | 0/0/0 | 2/2/2 | 0/0/0 | 3/3/3 | 2/2/2 | 2/2/2 | 2/2/2 | 2/2/2 | 0/0/0 | 0/0/0 | 0/0/0 | NA/NA/NA | 3/3/3 |
| Gitau, 2024 [45] | 3/3/3 | 2/3/3 | 2/2/2 | 0/0/0 | 2/2/2 | 2/2/2 | 0/0/0 | 2/2/2 | 1/1/1 | 3/3/3 | 2/2/2 | 2/2/2 | 2/2/2 | 2/2/2 | 0/0/0 | 0/0/0 | 0/0/0 | 0/0/0 | 0/0/0 |
| Grammatikopoulou, 2023 [46] | 3/3/3 | 3/3/3 | 0/0/0 | 0/0/0 | 2/2/2 | 2/3/3 | 0/0/0 | 0/0/0 | 0/0/0 | 3/3/3 | 2/2/2 | 2/2/2 | 2/2/2 | 2/2/2 | 0/0/0 | 0/0/0 | 0/0/0 | 2/2/2 | 0/0/0 |
| Guédon, 2021 [47] | 3/3/3 | 3/3/3 | 2/2/2 | 2/2/2 | 2/2/2 | 3/3/3 | 2/2/2 | 2/2/2 | 2/2/2 | 3/3/3 | 2/2/2 | 2/2/2 | 2/2/2 | 0/0/0 | 0/0/0 | 3/3/3 | 3/3/3 | 2/2/2 | 0/0/0 |
| Hasan, 2021 [48] | 3/3/3 | 0/2/2 | 0/2/2 | 0/2/2 | 2/3/2 | 1/1/1 | 0/0/0 | 2/2/2 | 1/1/1 | 3/3/3 | 2/2/2 | 2/2/2 | 2/3/2 | 2/2/2 | 0/0/0 | 0/2/2 | 0/0/0 | NA/NA/NA | 0/0/0 |
| Huang, 2022 [49] | 3/3/3 | 2/2/2 | 0/0/0 | 0/0/0 | 2/2/2 | 1/1/1 | 0/0/0 | 2/2/2 | 0/0/0 | 3/3/3 | 3/3/3 | 3/3/3 | 2/2/2 | 0/0/0 | 0/0/0 | 0/0/0 | 0/0/0 | NA/NA/NA | 0/0/0 |
| Jalal, 2022 [50] | 3/3/3 | 2/2/2 | 0/0/0 | 0/0/0 | 2/2/2 | 1/1/1 | 1/1/1 | 2/2/2 | 0/0/0 | 3/3/3 | 2/2/2 | 2/2/2 | 2/2/2 | 0/0/0 | 0/2/0 | 0/0/0 | 0/0/0 | NA/NA/NA | 0/0/0 |
| Jalal, 2023 [51] | 3/3/3 | 3/3/3 | 2/2/2 | 2/2/2 | 2/2/2 | 1/1/1 | 0/0/0 | 2/2/2 | 2/2/2 | 3/3/3 | 2/2/2 | 2/2/2 | 2/2/2 | 2/2/2 | 0/0/0 | 2/2/2 | 2/2/2 | NA/NA/NA | 2/2/2 |
| Jamal, 2024 [52] | 3/3/3 | 2/2/2 | 2/2/2 | 0/0/0 | 2/2/2 | 1/1/1 | 0/0/0 | 0/0/0 | 1/1/1 | 3/3/3 | 3/3/3 | 2/2/2 | 2/2/2 | 2/3/2 | 0/0/0 | 0/0/0 | 0/0/0 | NA/NA/NA | 0/0/0 |
| Jang, 2023 [53] | 2/2/2 | 0/0/0 | 0/0/0 | 0/0/0 | 2/2/2 | 0/0/0 | 0/0/0 | 0/0/0 | 0/0/0 | 3/3/3 | 0/0/0 | 2/2/2 | 2/2/2 | 0/0/0 | 2/2/2 | 0/0/0 | 0/0/0 | 0/0/0 | 0/0/0 |
| Jaspers, 2024 [54] | 3/3/3 | 2/2/2 | 0/0/0 | 0/0/0 | 2/2/2 | 1/1/1 | 0/0/0 | 2/2/2 | 1/1/1 | 3/3/3 | 2/2/2 | 2/2/2 | 2/2/2 | 2/2/2 | 0/0/0 | 0/0/0 | 0/0/0 | NA/NA/NA | 3/3/3 |
| Jearanai, 2023 [55] | 3/3/3 | 3/3/3 | 3/3/3 | 3/3/3 | 2/2/2 | 3/3/3 | 0/0/0 | 2/3/2 | 0/0/0 | 3/3/3 | 3/3/3 | 3/2/2 | 2/2/2 | 2/2/2 | 0/0/0 | 3/3/3 | 3/3/3 | 3/3/3 | 0/0/0 |
| Jha, 2021 [56] | 3/3/3 | 3/3/3 | 0/0/0 | 0/0/0 | 2/2/2 | 1/1/1 | 0/0/0 | 2/2/2 | 1/1/1 | 3/3/3 | 2/2/2 | 2/2/2 | 2/2/2 | 2/2/2 | 0/0/0 | 0/0/0 | 0/0/0 | NA/NA/NA | 0/0/0 |
| Jin, 2020 [57] | 3/3/3 | 3/3/3 | 2/2/2 | 2/2/2 | 3/3/3 | 1/1/1 | 0/0/0 | 2/2/2 | 1/1/1 | 3/3/3 | 3/3/3 | 2/2/2 | 3/3/3 | 2/2/2 | 2/2/2 | 0/0/0 | 0/0/0 | 0/0/0 | 3/3/3 |
| Jin, 2018 [58] | 3/3/3 | 3/3/3 | 2/2/2 | 2/2/2 | 2/2/2 | 1/1/1 | 0/0/0 | 2/2/2 | 2/2/2 | 3/3/3 | 2/2/2 | 2/2/2 | 2/2/2 | 0/0/0 | 0/0/0 | 2/2/2 | 0/0/0 | NA/NA/NA | 0/0/0 |
| Kamrul Hasan, 2021 [59] | 3/3/3 | 3/3/3 | 0/0/0 | 0/0/0 | 2/2/2 | 2/2/2 | 0/0/0 | 2/2/2 | 0/0/0 | 3/3/3 | 3/3/3 | 2/2/2 | 2/2/2 | 3/3/3 | 2/2/2 | 0/0/0 | 0/0/0 | 0/0/0 | 2/2/2 |
| Kanakatte, 2020 [60] | 3/2/2 | 2/3/3 | 0/2/2 | 0/2/2 | 2/3/2 | 1/1/1 | 0/0/0 | 2/3/2 | 2/2/2 | 3/3/3 | 2/3/2 | 2/2/2 | 2/2/2 | 0/2/2 | 0/0/0 | 0/0/0 | 0/0/0 | NA/NA/NA | 0/0/0 |
| Kawamura, 2023 [61] | 3/3/3 | 3/3/3 | 3/3/3 | 3/3/3 | 2/2/2 | 2/2/2 | 2/2/2 | 2/2/2 | 0/0/0 | 3/3/3 | 2/2/2 | 2/2/2 | 2/2/2 | 2/2/2 | 0/0/0 | 3/3/3 | 3/3/3 | 3/3/3 | 3/3/3 |
| Khalid, 2023 [62] | 3/3/3 | 2/2/2 | 2/2/2 | 0/0/0 | 2/2/2 | 1/1/1 | 0/0/0 | 2/2/2 | 2/2/2 | 3/3/3 | 3/3/3 | 2/2/2 | 2/2/2 | 0/0/0 | 0/0/0 | 0/0/0 | 2/2/2 | 0/0/0 | 3/3/3 |
| Khalid, 2023 [63] | 3/3/3 | 3/3/3 | 3/3/3 | 3/3/3 | 2/2/2 | 2/2/2 | 2/2/2 | 2/2/2 | 2/2/2 | 3/3/3 | 1/1/1 | 2/2/2 | 1/1/1 | 0/0/0 | 2/2/2 | 3/3/3 | 3/3/3 | 3/3/3 | 0/0/0 |
| Kim, 2024 [64] | 3/3/3 | 2/2/2 | 0/2/2 | 0/2/2 | 2/2/2 | 1/1/1 | 0/2/0 | 2/2/2 | 1/1/1 | 3/3/3 | 2/2/2 | 2/2/2 | 2/2/2 | 0/0/0 | 0/0/0 | 0/0/0 | 0/0/0 | NA/NA/NA | 0/0/0 |
| Kinoshita, 2024 [65] | 3/3/3 | 3/3/3 | 3/3/3 | 3/3/3 | 2/2/2 | 3/3/3 | 2/2/2 | 0/0/0 | 3/2/3 | 2/2/2 | 0/0/0 | 2/2/2 | 2/2/2 | 2/3/3 | 0/0/0 | 2/2/2 | 3/3/3 | 3/3/3 | 0/0/0 |
| Kitaguchi, 2023 [66] | 3/3/3 | 3/3/3 | 2/3/3 | 2/2/2 | 2/3/2 | 2/2/2 | 0/0/0 | 2/2/2 | 3/3/3 | 3/3/3 | 2/2/2 | 2/2/2 | 2/2/2 | 0/0/0 | 0/0/0 | 3/3/3 | 3/3/3 | 3/3/3 | 0/0/0 |
| Kitaguchi, 2022 [67] | 3/3/3 | 3/3/3 | 2/2/2 | 2/2/2 | 2/2/2 | 2/2/2 | 2/2/2 | 2/2/2 | 3/3/3 | 3/3/3 | 2/2/2 | 2/2/2 | 2/2/2 | 2/2/2 | 0/0/0 | 2/2/2 | 3/3/3 | 3/3/3 | 2/2/2 |
| Kitaguchi, 2022 [68] | 3/3/3 | 3/3/3 | 2/3/3 | 3/3/3 | 2/2/2 | 3/3/3 | 2/2/2 | 2/3/2 | 2/3/3 | 3/3/3 | 3/3/3 | 2/2/2 | 3/3/3 | 3/3/3 | 2/2/2 | 3/3/3 | 3/3/3 | 3/3/3 | 2/2/2 |
| Kletz, 2019 [69] | 3/3/3 | 2/2/2 | 2/2/2 | 2/2/2 | 2/2/2 | 2/2/2 | 0/0/0 | 2/2/2 | 0/0/0 | 3/3/3 | 2/2/2 | 2/2/2 | 2/2/2 | 2/2/2 | 0/0/0 | 2/2/2 | 0/0/0 | 0/0/0 | 0/0/0 |
| Kolbinger, 2023 [70] | 3/3/3 | 3/3/3 | 3/3/3 | 3/3/3 | 2/2/2 | 2/2/2 | 1/1/1 | 2/2/2 | 1/1/1 | 3/3/3 | 2/2/2 | 3/3/3 | 3/3/3 | 2/2/2 | 0/0/0 | 3/3/3 | 3/3/3 | 3/3/3 | 3/3/3 |
| Kolbinger, 2024 [71] | 3/3/3 | 3/3/3 | 3/3/3 | 3/3/3 | 2/2/2 | 3/3/3 | 3/3/3 | 3/3/3 | 3/3/3 | 3/3/3 | 2/2/2 | 2/2/2 | 2/2/2 | 2/2/2 | 0/0/0 | 2/2/2 | 3/3/3 | 3/0/3 | 3/0/3 |
| Kolbinger, 2024 [72] | 3/3/3 | 3/3/3 | 2/2/2 | 0/0/0 | 2/2/2 | 1/1/1 | 0/0/0 | 2/2/2 | 1/0/1 | 3/3/3 | 1/1/1 | 2/2/2 | 3/3/3 | 2/2/2 | 0/0/0 | 0/0/0 | 3/3/3 | NA/NA/NA | 0/0/0 |
| Kondo, 2021 [73] | 3/3/3 | 3/3/3 | 2/2/2 | 2/2/2 | 2/2/2 | 1/1/1 | 0/0/0 | 2/2/2 | 1/1/1 | 3/3/3 | 3/3/3 | 3/3/3 | 2/2/2 | 2/2/2 | 0/0/0 | 0/0/0 | 0/0/0 | NA/NA/NA | 0/0/0 |
| Konduri, 2024 [74] | 3/3/3 | 2/2/2 | 0/0/0 | 0/0/0 | 2/2/2 | 1/1/1 | 0/0/0 | 2/2/2 | 1/1/1 | 3/3/3 | 2/2/2 | 2/2/2 | 2/2/2 | 2/2/2 | 0/0/0 | 0/0/0 | 2/2/2 | NA/NA/NA | 0/0/0 |
| Kong, 2021 [75] | 3/3/3 | 3/3/3 | 2/2/2 | 2/2/2 | 2/2/2 | 2/2/2 | 0/0/0 | 3/3/3 | 3/3/3 | 3/3/3 | 3/3/3 | 3/3/3 | 3/3/3 | 2/2/2 | 0/2/0 | 0/0/0 | 0/0/0 | NA/NA/NA | 3/3/3 |
| Kumazu, 2021 [76] | 3/3/3 | 3/3/3 | 3/3/3 | 3/3/3 | 2/2/2 | 2/2/2 | 3/3/3 | 2/2/2 | 2/2/2 | 3/3/3 | 2/2/2 | 1/1/1 | 2/2/2 | 2/2/2 | 3/3/3 | 3/3/3 | 3/3/3 | 3/3/3 | 0/0/0 |
| Kumazu, 2025 [77] | 3/3/3 | 3/3/3 | 3/3/3 | 3/3/3 | 2/2/2 | 2/2/2 | 2/2/2 | 2/2/2 | 2/2/2 | 3/3/3 | 2/2/2 | 2/2/2 | 2/2/2 | 0/0/0 | 2/2/2 | 3/3/3 | 3/3/3 | 3/3/3 | 0/0/0 |
| Labrunie, 2022 [78] | 3/3/3 | 2/2/2 | 2/2/2 | 2/2/2 | 2/2/2 | 2/2/2 | 0/0/0 | 2/2/2 | 0/0/0 | 2/2/2 | 2/2/2 | 2/2/2 | 2/2/2 | 2/2/2 | 2/2/2 | 2/2/2 | 2/2/2 | 3/3/3 | 0/0/0 |
| Lam, 2022 [79] | 3/3/3 | 3/3/3 | 2/2/2 | 2/3/2 | 2/2/2 | 2/2/2 | 0/0/0 | 2/2/2 | 3/3/3 | 3/3/3 | 2/2/2 | 2/2/2 | 2/2/2 | 0/0/0 | 0/0/0 | 0/0/0 | 2/2/2 | 0/0/0 | 0/0/0 |
| Laplante, 2022 [80] | 3/3/3 | 3/3/3 | 3/3/3 | 2/2/2 | 2/2/2 | 2/2/2 | 3/3/3 | 0/0/0 | 3/3/3 | 1/1/1 | 0/0/0 | 1/1/1 | 2/2/2 | 0/0/0 | 0/0/0 | 3/3/3 | 2/2/2 | 0/0/0 | 1/1/1 |
| Lavanchy, 2021 [81] | 3/3/3 | 3/3/3 | 2/2/2 | 2/3/3 | 2/2/2 | 3/3/3 | 0/0/0 | 2/2/2 | 2/2/2 | 3/3/3 | 2/2/2 | 3/3/3 | 3/3/3 | 2/3/2 | 2/2/2 | 3/3/3 | 0/0/0 | 3/3/3 | 0/0/0 |
| Le, 2023 [82] | 3/3/3 | 2/2/2 | 0/0/0 | 0/0/0 | 2/2/2 | 1/1/1 | 0/0/0 | 2/2/2 | 1/1/1 | 3/3/3 | 2/2/2 | 2/2/2 | 2/2/2 | 2/2/2 | 0/0/0 | 0/0/0 | 0/0/0 | NA/NA/NA | 0/0/0 |
| Lee, 2024 [83] | 3/3/3 | 2/2/2 | 2/2/2 | 0/0/0 | 2/2/2 | 1/1/1 | 0/0/0 | 2/2/2 | 1/1/1 | 3/3/3 | 3/3/3 | 2/3/2 | 3/3/3 | 3/3/3 | 0/0/0 | 0/0/0 | 0/0/0 | NA/NA/NA | 0/0/0 |
| Leifman, 2024 [84] | 3/3/3 | 3/3/3 | 2/2/2 | 0/0/0 | 3/3/3 | 1/1/1 | 0/0/0 | 0/0/0 | 1/1/1 | 3/3/3 | 2/2/2 | 1/1/1 | 2/2/2 | 2/2/2 | 0/0/0 | 0/0/0 | 0/0/0 | NA/NA/NA | 0/0/0 |
| Leifmann, 2022 [85] | 3/3/3 | 3/3/3 | 3/3/3 | 3/3/3 | 2/2/2 | 3/3/3 | 2/2/2 | 2/2/2 | 3/3/3 | 3/3/3 | 2/2/2 | 2/2/2 | 3/3/3 | 2/2/2 | 2/3/2 | 3/3/3 | 3/3/3 | 3/3/3 | 0/0/0 |
| Li, 2025 [86] | 3/3/3 | 3/3/3 | 3/3/3 | 3/3/3 | 2/2/2 | 1/1/1 | 0/0/0 | 2/2/2 | 1/1/1 | 3/3/3 | 2/2/2 | 2/2/2 | 2/3/2 | 2/3/2 | 0/0/0 | 2/3/3 | 2/2/2 | 0/0/0 | 0/0/0 |
| Li, 2023 [87] | 3/3/3 | 3/3/3 | 2/2/2 | 0/0/0 | 2/2/2 | 1/1/1 | 0/0/0 | 0/0/0 | 1/1/1 | 3/3/3 | 2/2/2 | 2/3/3 | 2/3/3 | 0/0/0 | 0/0/0 | 2/2/2 | 0/0/0 | NA/NA/NA | 3/3/3 |
| Liao, 2024 [88] | 3/3/3 | 0/0/0 | 0/0/0 | 0/2/2 | 2/2/2 | 1/1/1 | 0/0/0 | 2/2/2 | 1/1/1 | 3/3/3 | 3/2/3 | 2/2/2 | 2/2/2 | 3/3/3 | 0/0/0 | 0/2/0 | 0/2/0 | NA/NA/NA | 0/0/0 |
| Lin, 2024 [89] | 3/3/3 | 2/2/2 | 0/0/0 | 0/0/0 | 2/2/2 | 1/1/1 | 0/0/0 | 2/2/2 | 1/1/1 | 3/3/3 | 3/3/3 | 2/2/2 | 2/2/2 | 2/2/2 | 2/2/2 | 0/0/0 | 0/0/0 | NA/NA/NA | 2/2/2 |
| Lin, 2021 [90] | 3/3/3 | 3/3/3 | 0/0/0 | 0/0/0 | 2/2/2 | 1/1/1 | 2/2/2 | 2/2/2 | 1/1/1 | 3/3/3 | 3/3/3 | 2/2/2 | 3/3/3 | 3/3/3 | 0/0/0 | 0/0/0 | 0/0/0 | NA/NA/NA | 0/0/0 |
| Liu, 2022 [91] | 3/3/3 | 3/3/3 | 3/3/3 | 3/3/3 | 3/3/3 | 3/3/3 | 3/3/3 | 2/2/2 | 3/3/3 | 3/3/3 | 2/2/2 | 3/3/3 | 3/3/3 | 3/3/3 | 2/2/2 | 3/3/3 | 3/3/3 | 3/3/3 | 0/0/0 |
| Liu, 2024 [92] | 3/3/3 | 3/3/3 | 2/2/2 | 2/2/2 | 2/2/2 | 2/2/2 | 2/2/2 | 2/2/2 | 0/0/0 | 3/3/3 | 3/3/3 | 2/2/2 | 3/3/3 | 2/3/3 | 0/0/0 | 0/0/0 | 0/0/0 | 0/0/0 | 3/3/3 |
| Liu, 2022 [93] | 3/3/3 | 2/2/2 | 2/2/2 | 2/2/2 | 2/2/2 | 1/1/1 | 0/0/0 | 2/2/2 | 2/2/2 | 3/3/3 | 3/3/3 | 2/2/2 | 2/2/2 | 2/2/2 | 2/2/2 | 0/0/0 | 2/2/2 | NA/NA/NA | 0/0/0 |
| Lou, 2023 [94] | 3/3/3 | 3/3/3 | 0/0/0 | 0/0/0 | 3/3/3 | 1/1/1 | 0/0/0 | 2/2/2 | 1/1/1 | 3/3/3 | 3/3/3 | 2/2/2 | 2/2/2 | 2/2/2 | 3/3/3 | 0/0/0 | 0/0/0 | NA/NA/NA | 0/0/0 |
| Loukas, 2020 [95] | 3/3/3 | 3/3/3 | 2/2/2 | 0/0/0 | 2/2/2 | 1/1/1 | 0/0/0 | 2/2/2 | 1/1/1 | 3/3/3 | 2/2/2 | 3/3/3 | 2/2/2 | 2/2/2 | 2/2/2 | 2/2/2 | 2/2/2 | NA/NA/NA | 0/0/0 |
| Loza, 2024 [96] | 3/3/3 | 3/3/3 | 2/2/2 | 2/2/2 | 2/2/2 | 1/1/1 | 0/0/0 | 2/2/2 | 1/1/1 | 3/3/3 | 3/2/2 | 2/2/2 | 2/2/2 | 2/2/2 | 0/0/0 | 2/2/2 | 0/0/0 | NA/NA/NA | 0/0/0 |
| Maack, 2024 [97] | 3/3/3 | 3/3/3 | 2/2/2 | 2/2/2 | 2/2/2 | 1/1/1 | 0/0/0 | 2/2/2 | 1/1/1 | 3/3/3 | 2/0/2 | 2/2/2 | 2/3/2 | 2/2/2 | 0/0/0 | 0/0/0 | 0/0/0 | NA/NA/NA | 3/3/3 |
| Madad Zadeh, 2020 [98] | 3/3/3 | 2/2/2 | 0/0/0 | 0/0/0 | 2/2/2 | 2/2/2 | 0/0/0 | 0/0/0 | 2/2/2 | 3/3/3 | 0/0/0 | 2/2/2 | 2/2/2 | 0/0/0 | 0/0/0 | 2/2/2 | 2/2/2 | 3/3/3 | 2/2/2 |
| Madani, 2022 [99] | 3/3/3 | 3/3/3 | 3/3/3 | 3/3/3 | 2/3/2 | 2/2/2 | 2/2/2 | 2/2/2 | 3/3/3 | 3/3/3 | 3/3/3 | 2/2/2 | 2/2/2 | 0/0/0 | 2/2/2 | 3/3/3 | 3/3/3 | 0/0/0 | 0/0/0 |
| Maqbool, 2020 [100] | 3/3/3 | 3/3/3 | 2/2/2 | 2/2/2 | 3/2/3 | 1/1/1 | 0/0/0 | 2/2/2 | 1/1/1 | 3/3/3 | 2/2/2 | 2/2/2 | 2/2/2 | 2/2/2 | 0/0/0 | 0/0/0 | 0/0/0 | NA/NA/NA | 3/3/3 |
| Marullo, 2023 [101] | 3/3/3 | 3/3/3 | 2/2/2 | 2/2/2 | 0/0/0 | 2/3/3 | 0/0/0 | 2/2/2 | 2/2/2 | 3/3/3 | 2/3/2 | 2/2/2 | 2/2/2 | 0/0/0 | 0/0/0 | 2/2/2 | 2/2/2 | 3/3/3 | 0/0/0 |
| Mascagni, 2022 [102] | 3/3/3 | 3/3/3 | 3/3/3 | 3/3/3 | 2/2/2 | 3/3/3 | 0/0/0 | 2/2/2 | 3/3/3 | 3/3/3 | 2/3/3 | 2/2/2 | 3/3/3 | 2/2/2 | 0/0/0 | 3/3/3 | 3/3/3 | 3/3/3 | 0/0/0 |
| Matsumoto, 2024 [103] | 3/3/3 | 3/3/3 | 3/3/3 | 2/2/2 | 2/2/2 | 2/2/2 | 0/0/0 | 2/2/2 | 0/0/0 | 2/2/2 | 0/0/0 | 2/2/2 | 0/0/0 | 0/0/0 | 0/0/0 | 3/3/3 | 3/3/3 | 3/3/3 | 3/3/3 |
| Mehta, 2024 [104] | 3/3/3 | 2/2/2 | 2/2/2 | 2/2/2 | 2/2/2 | 2/2/2 | 0/0/0 | 2/2/2 | 0/0/0 | 3/3/3 | 3/3/3 | 3/3/3 | 2/3/2 | 2/2/2 | 2/2/2 | 2/2/2 | 2/2/2 | 3/3/3 | 0/0/0 |
| Mishra, 2017 [105] | 3/3/3 | 2/2/2 | 2/2/2 | 0/0/0 | 0/0/0 | 1/1/1 | 0/0/0 | 2/2/2 | 1/1/1 | 3/3/3 | 3/3/3 | 2/2/2 | 2/2/2 | 0/0/0 | 2/2/2 | 0/0/0 | 0/0/0 | NA/NA/NA | 3/3/3 |
| Murali, 2024 [106] | 3/3/3 | 2/2/2 | 0/0/0 | 0/0/0 | 2/2/2 | 1/1/1 | 0/0/0 | 0/0/0 | 1/1/1 | 3/3/3 | 2/3/3 | 0/3/2 | 2/2/2 | 2/3/2 | 0/0/0 | 0/0/0 | 0/0/0 | NA/NA/NA | 0/0/0 |
| Murali, 2023 [107] | 3/3/3 | 2/2/2 | 0/0/0 | 0/0/0 | 2/2/2 | 1/1/1 | 0/0/0 | 0/0/0 | 1/1/1 | 3/3/3 | 3/2/2 | 2/2/2 | 2/2/2 | 2/2/2 | 0/0/0 | 0/0/0 | 0/0/0 | NA/NA/NA | 0/0/0 |
| Murali, 2022 [108] | 3/3/3 | 3/3/3 | 2/2/2 | 2/2/2 | 2/2/2 | 2/2/2 | 0/0/0 | 2/2/2 | 2/2/2 | 3/3/3 | 3/3/3 | 2/2/2 | 3/3/3 | 2/2/2 | 2/2/2 | 0/0/0 | 0/0/0 | 0/0/0 | 0/0/0 |
| Myo, 2024 [109] | 3/3/3 | 3/3/3 | 2/2/2 | 2/2/2 | 2/2/2 | 1/1/1 | 0/0/0 | 3/3/3 | 3/3/3 | 3/3/3 | 3/3/3 | 3/3/3 | 2/2/2 | 2/2/2 | 0/0/0 | 3/3/3 | 3/3/3 | 3/3/3 | 0/0/0 |
| Nakanuma, 2023 [110] | 3/3/3 | 3/3/3 | 3/3/3 | 3/3/3 | 2/3/3 | 3/3/3 | 0/0/0 | 2/2/2 | 3/3/3 | 3/3/3 | 2/2/2 | 3/3/3 | 2/2/2 | 2/2/2 | 3/3/3 | 3/3/3 | 3/3/3 | 3/3/3 | 0/0/0 |
| Namazi, 2022 [111] | 3/3/3 | 3/3/3 | 2/2/2 | 2/2/2 | 2/3/2 | 2/2/2 | 1/1/1 | 2/2/2 | 1/1/1 | 3/3/3 | 3/3/3 | 2/3/2 | 2/2/2 | 3/3/3 | 0/0/0 | 2/2/2 | 0/2/0 | NA/NA/NA | 1/1/1 |
| Nema , 2023 [112] | 3/3/3 | 2/2/2 | 2/2/2 | 0/0/0 | 3/3/3 | 1/1/1 | 2/2/2 | 0/0/0 | 1/1/1 | 3/3/3 | 3/3/3 | 2/2/2 | 2/2/2 | 0/0/0 | 0/0/0 | 0/0/0 | 0/0/0 | NA/NA/NA | 0/0/0 |
| Nwoye, 2024 [113] | 3/3/3 | 3/3/3 | 2/2/2 | 2/2/2 | 2/3/2 | 1/1/1 | 0/0/0 | 3/3/3 | 1/1/1 | 3/3/3 | 3/3/3 | 2/2/2 | 3/3/3 | 2/2/2 | 3/3/3 | 2/2/2 | 3/3/3 | NA/NA/NA | 2/2/2 |
| Nwoye, 2019 [114] | 3/3/3 | 3/3/3 | 0/0/0 | 0/0/0 | 2/2/2 | 1/1/1 | 0/0/0 | 2/2/2 | 0/0/0 | 3/3/3 | 2/2/2 | 2/2/2 | 2/2/2 | 0/0/0 | 2/2/2 | 0/0/0 | 0/0/0 | NA/NA/NA | 0/0/0 |
| Oh, 2024 [115] | 3/3/3 | 3/3/3 | 3/3/3 | 3/3/3 | 3/3/3 | 3/3/3 | 3/3/3 | 2/2/2 | 3/3/3 | 3/3/3 | 3/3/3 | 3/3/3 | 2/2/2 | 2/2/2 | 0/0/0 | 3/3/3 | 3/3/3 | 3/3/3 | 0/0/0 |
| Owen, 2022 [116] | 3/3/3 | 3/3/3 | 2/3/3 | 2/2/2 | 2/2/2 | 0/0/0 | 0/0/0 | 2/2/2 | 2/2/2 | 3/3/3 | 2/2/2 | 3/3/3 | 2/2/2 | 2/2/2 | 2/2/2 | 2/2/2 | 0/0/0 | NA/NA/NA | 0/0/0 |
| Ozbulak, 2025 [117] | 3/3/3 | 2/3/3 | 2/2/2 | 2/2/2 | 2/2/2 | 1/1/1 | 0/0/0 | 0/0/0 | 1/1/1 | 3/3/3 | 2/3/3 | 2/2/2 | 2/2/2 | 0/0/0 | 2/2/2 | 2/2/2 | 0/0/0 | NA/NA/NA | 0/0/0 |

| | | | | | | | | | | | | | | | | | | |
|---|---|---|---|---|---|---|---|---|---|---|---|---|---|---|---|---|---|---|
| Pan, 2023 [118] | 3/3/3 | 3/3/3 | 2/2/2 | 2/3/3 | 2/2/2 | 2/2/2 | 0/0/0 | 2/2/2 | 0/0/0 | 3/3/3 | 3/3/3 | 2/2/2 | 2/2/2 | 2/2/2 | 0/0/0 | 0/0/0 | 0/0/0 | NA/NA/NA | 0/0/0 |
| Penza, 2018 [119] | 2/2/2 | 2/2/2 | 0/0/0 | 0/0/0 | 0/0/0 | 2/2/2 | 0/0/0 | 2/3/2 | 0/0/0 | 3/3/3 | 2/2/2 | 0/0/0 | 3/2/2 | 0/0/0 | 2/2/2 | 0/0/0 | 0/0/0 | 0/0/0 | 0/0/0 |
| Pradeep, 2022 [120] | 3/3/3 | 2/2/2 | 0/0/0 | 0/0/0 | 2/2/2 | 1/1/1 | 0/0/0 | 2/2/2 | 1/1/1 | 3/3/3 | 2/3/2 | 2/2/2 | 2/2/2 | 2/2/2 | 0/0/0 | 0/0/0 | 0/0/0 | NA/NA/NA | 0/0/0 |
| Prokopetc, 2015 [121] | 3/3/3 | 3/3/3 | 2/2/2 | 2/2/2 | 0/2/2 | 0/0/0 | 0/0/0 | 0/0/0 | 2/2/2 | 2/2/2 | 0/0/0 | 2/2/2 | 2/2/2 | 0/2/2 | 0/3/2 | 0/2/0 | 0/2/0 | 0/0/0 | 0/0/0 |
| Protserov, 2024 [122] | 3/3/3 | 3/3/3 | 3/3/3 | 3/3/3 | 2/2/2 | 3/3/3 | 0/0/0 | 2/2/2 | 2/2/2 | 3/3/3 | 3/3/3 | 2/2/2 | 3/3/3 | 2/3/3 | 3/3/3 | 3/3/3 | 3/3/3 | 3/3/3 | 3/3/3 |
| Rahbar, 2020 [123] | 3/3/3 | 3/3/3 | 3/3/3 | 3/3/3 | 2/2/2 | 2/2/2 | 0/0/0 | 2/2/2 | 2/2/2 | 2/2/2 | 0/0/0 | 2/2/2 | 2/2/2 | 2/0/0 | 3/3/3 | 3/3/3 | 3/3/3 | 0/0/0 | 0/0/0 |
| Rahbar, 2024 [124] | 3/3/3 | 3/3/3 | 2/2/2 | 2/2/2 | 2/2/2 | 1/1/1 | 0/0/0 | 2/2/2 | 1/1/1 | 3/3/3 | 2/2/2 | 0/0/0 | 2/2/2 | 0/0/0 | 0/0/0 | 2/2/2 | 2/2/2 | NA/NA/NA | 2/2/2 |
| Raja, 2024 [125] | 3/3/3 | 2/2/2 | 0/2/0 | 0/3/0 | 0/2/0 | 1/1/1 | 0/0/0 | 2/2/2 | 1/1/1 | 3/3/3 | 2/3/2 | 2/2/2 | 2/2/2 | 0/2/2 | 0/0/0 | 0/0/0 | 0/0/0 | NA/0/NA | 0/3/0 |
| Ryu, 2024 [126] | 3/3/3 | 3/3/3 | 2/2/2 | 2/2/2 | 2/3/2 | 3/3/3 | 0/0/0 | 2/2/2 | 3/3/3 | 3/3/3 | 3/3/3 | 2/2/2 | 2/2/2 | 2/2/2 | 0/0/0 | 3/3/3 | 3/3/3 | 3/3/3 | 0/0/0 |
| Sahu, 2017 [127] | 3/3/3 | 2/2/2 | 2/2/2 | 2/2/2 | 2/2/2 | 1/1/1 | 1/1/1 | 2/2/2 | 1/1/1 | 3/3/3 | 2/2/2 | 2/2/2 | 3/3/3 | 3/3/3 | 3/3/3 | 0/0/0 | 2/2/2 | NA/NA/NA | 0/0/0 |
| Samuel, 2021 [128] | 3/3/3 | 2/2/2 | 0/0/0 | 0/0/0 | 2/2/2 | 1/1/1 | 0/0/0 | 2/2/2 | 0/0/0 | 3/3/3 | 2/2/2 | 2/2/2 | 2/2/2 | 0/0/0 | 2/2/2 | 0/0/0 | 0/0/0 | NA/NA/NA | 0/0/0 |
| Sanchez-Matilla, 2022 [129] | 3/3/3 | 2/2/2 | 0/0/0 | 0/0/0 | 2/2/2 | 1/1/1 | 0/0/0 | 2/2/2 | 1/1/1 | 3/3/3 | 3/3/3 | 2/2/2 | 2/3/2 | 2/2/2 | 0/0/0 | 0/0/0 | 0/0/0 | NA/NA/NA | 0/0/0 |
| Satyanaik, 2024 [130] | 3/3/3 | 3/3/3 | 2/2/2 | 0/0/0 | 2/2/2 | 1/1/1 | 0/0/0 | 2/2/2 | 1/1/1 | 3/3/3 | 2/2/2 | 2/2/2 | 2/2/2 | 2/2/2 | 0/0/0 | 0/0/0 | 0/0/0 | NA/NA/NA | 3/3/3 |
| Satyanaik, 2024 [131] | 2/2/2 | 2/2/2 | 2/2/2 | 2/2/2 | 2/2/2 | 3/3/3 | 0/0/0 | 2/2/2 | 2/2/2 | 3/3/3 | 2/2/2 | 2/2/2 | 2/2/2 | 2/2/2 | 0/0/0 | 2/2/2 | 0/0/0 | 3/3/3 | 3/3/3 |
| Seenivasan, 2023 [132] | 3/3/3 | 3/3/3 | 3/3/3 | 3/3/3 | 2/2/2 | 1/1/1 | 0/1/0 | 2/2/2 | 0/1/1 | 3/3/3 | 3/3/3 | 2/2/2 | 2/2/2 | 0/0/0 | 0/0/0 | 2/2/2 | 0/0/0 | NA/NA/NA | 3/3/3 |
| Sengun, 2023 [133] | 3/3/3 | 3/3/3 | 3/3/3 | 2/2/2 | 2/2/2 | 3/3/3 | 0/0/0 | 0/0/0 | 3/3/3 | 3/3/3 | 2/2/2 | 2/2/2 | 2/2/2 | 2/2/2 | 0/0/0 | 3/3/3 | 3/2/2 | 3/3/3 | 0/0/0 |
| Sengun, 2024 [134] | 3/3/3 | 3/3/3 | 2/2/2 | 3/3/3 | 2/2/2 | 3/3/3 | 2/2/2 | 0/0/0 | 3/3/3 | 3/3/3 | 0/0/0 | 3/3/3 | 2/2/2 | 0/0/0 | 0/0/0 | 3/3/3 | 2/2/2 | 3/3/3 | 0/0/0 |
| Shen, 2023 [135] | 3/3/3 | 2/2/2 | 2/2/2 | 2/2/2 | 2/2/2 | 1/1/1 | 0/0/0 | 2/2/2 | 1/1/1 | 3/3/3 | 3/3/3 | 2/2/2 | 2/2/2 | 2/2/2 | 0/0/0 | 0/0/0 | 0/0/0 | 0/0/0 | 3/3/3 |
| Sheng, 2024 [136] | 3/3/3 | 3/3/3 | 2/2/2 | 2/2/2 | 2/2/2 | 1/1/1 | 0/0/0 | 0/0/0 | 1/1/1 | 3/3/3 | 2/2/2 | 0/0/0 | 2/2/2 | 0/0/0 | 0/0/0 | 0/0/0 | 2/2/2 | NA/NA/NA | 3/3/3 |
| Shi, 2020 [137] | 3/3/3 | 3/3/3 | 2/2/2 | 2/2/2 | 2/2/2 | 1/1/1 | 0/0/0 | 2/2/2 | 0/0/0 | 3/3/3 | 3/3/3 | 2/2/2 | 2/2/2 | 2/3/2 | 0/0/0 | 2/3/2 | 2/2/2 | 0/0/0 | 0/0/0 |
| Shimgekar, 2021 [138] | 3/3/3 | 2/2/2 | 0/0/0 | 0/0/0 | 2/2/2 | 2/2/2 | 0/0/0 | 0/0/0 | 2/2/2 | 2/2/2 | 0/0/0 | 2/2/2 | 0/0/0 | 0/0/0 | 0/0/0 | 2/2/2 | 0/0/0 | 0/0/0 | 0/0/0 |
| Silva, 2022 [139] | 3/3/3 | 3/3/3 | 2/2/2 | 2/2/2 | 2/2/2 | 2/2/2 | 0/0/0 | 2/2/2 | 1/1/1 | 3/3/3 | 2/2/2 | 2/2/2 | 2/2/2 | 2/3/2 | 0/0/0 | 0/0/0 | 0/0/0 | NA/NA/NA | 0/0/0 |
| Smithmaitrie, 2024 [140] | 3/3/3 | 3/3/3 | 3/3/3 | 3/3/3 | 2/2/2 | 2/2/2 | 2/2/2 | 2/2/2 | 3/3/3 | 3/3/3 | 2/2/2 | 2/2/2 | 2/2/2 | 2/3/2 | 0/0/0 | 3/3/3 | 2/2/2 | 3/3/3 | 2/2/2 |
| Sonsilphong, 2022 [141] | 3/3/3 | 0/0/0 | 0/0/0 | 0/0/0 | 0/0/0 | 2/2/2 | 0/0/0 | 2/2/2 | 2/2/2 | 3/3/3 | 2/2/2 | 2/2/2 | 2/2/2 | 2/2/2 | 0/0/0 | 0/0/0 | 0/0/0 | 0/0/0 | 0/0/0 |
| Streckert, 2023 [142] | 3/3/3 | 3/3/3 | 0/0/0 | 0/0/0 | 2/2/2 | 1/1/1 | 0/0/0 | 2/2/2 | 1/1/1 | 3/3/3 | 2/2/2 | 2/2/2 | 2/2/2 | 0/0/0 | 0/0/0 | 0/0/0 | 0/0/0 | NA/NA/NA | 0/0/0 |
| Strong, 2024 [143] | 3/3/3 | 3/3/3 | 3/3/3 | 3/3/3 | 3/3/3 | 3/3/3 | 2/2/2 | 2/2/2 | 3/3/3 | 2/2/2 | 2/2/2 | 2/2/2 | 2/2/2 | 2/2/2 | 0/0/0 | 3/3/3 | 3/3/3 | 3/3/3 | 0/0/0 |
| Sun, 2022 [144] | 3/3/3 | 3/3/3 | 2/2/2 | 2/2/2 | 2/2/2 | 0/0/0 | 0/0/0 | 2/2/2 | 0/0/0 | 3/3/3 | 3/3/3 | 2/2/2 | 2/2/2 | 2/2/2 | 3/3/3 | 2/2/2 | 0/0/0 | NA/NA/NA | 2/2/2 |
| Takeuchi, 2023 [145] | 3/3/3 | 3/3/3 | 3/3/3 | 3/3/3 | 2/2/2 | 3/3/3 | 2/2/2 | 2/2/2 | 2/2/2 | 3/3/3 | 2/2/2 | 2/2/2 | 2/2/2 | 0/0/0 | 0/0/0 | 3/3/3 | 3/3/3 | 3/3/3 | 2/2/2 |
| Tao, 2023 [146] | 3/3/3 | 3/3/3 | 2/2/2 | 2/2/2 | 2/2/2 | 1/1/1 | 0/0/0 | 2/2/2 | 1/1/1 | 3/3/3 | 3/3/3 | 2/2/2 | 3/3/3 | 3/3/3 | 0/0/0 | 2/2/2 | 0/0/0 | NA/NA/NA | 0/0/0 |
| Tokuyasu, 2021 [147] | 3/3/3 | 3/3/3 | 3/3/3 | 3/3/3 | 2/2/2 | 2/2/2 | 3/3/3 | 2/3/2 | 3/3/3 | 3/3/3 | 2/2/2 | 2/2/2 | 2/2/2 | 2/3/2 | 0/0/0 | 3/3/3 | 2/2/2 | 2/2/2 | 0/0/0 |
| Urrea, 2024 [148] | 3/3/3 | 3/3/3 | 2/2/2 | 2/2/2 | 2/2/2 | 1/1/1 | 0/0/0 | 3/3/3 | 1/1/1 | 3/3/3 | 3/3/3 | 2/2/2 | 2/2/2 | 2/2/2 | 2/2/2 | 2/2/2 | 2/2/2 | NA/NA/NA | 2/2/2 |
| Valderrama, 2022 [149] | 3/3/3 | 3/3/3 | 2/2/2 | 2/2/2 | 2/2/2 | 2/2/2 | 2/2/2 | 2/2/2 | 2/2/2 | 3/3/3 | 2/2/2 | 2/2/2 | 2/2/2 | 0/0/0 | 0/0/0 | 0/0/0 | 0/0/0 | 3/3/3 | 3/3/3 |
| Vardazaryan, 2018 [150] | 3/3/3 | 2/2/2 | 0/0/0 | 0/0/0 | 0/0/0 | 1/1/1 | 0/0/0 | 2/2/2 | 0/0/0 | 3/3/3 | 2/2/2 | 2/2/2 | 2/2/2 | 0/0/0 | 2/2/2 | 0/0/0 | 0/0/0 | 0/0/0 | 0/0/0 |
| Wagner, 2023 [151] | 3/3/3 | 3/3/3 | 3/3/3 | 3/3/3 | 2/3/2 | 3/3/3 | 2/2/2 | 2/2/2 | 3/3/3 | 3/3/3 | 1/1/1 | 1/3/3 | 3/3/3 | 3/3/3 | 3/3/3 | 3/3/3 | 3/3/3 | NA/NA/NA | 3/3/3 |
| Wang, 2023 [152] | 3/3/3 | 3/3/3 | 2/2/2 | 2/2/2 | 3/3/3 | 1/1/1 | 1/1/1 | 2/2/2 | 1/1/1 | 3/3/3 | 2/2/2 | 2/2/2 | 2/2/2 | 2/2/2 | 0/0/0 | 0/0/0 | 0/0/0 | NA/NA/NA | 0/0/0 |
| Wang, 2017 [153] | 3/3/3 | 2/2/2 | 2/2/2 | 0/3/2 | 2/2/2 | 1/1/1 | 0/0/0 | 2/2/2 | 1/1/1 | 3/3/3 | 2/2/2 | 2/2/2 | 2/2/2 | 0/0/0 | 0/0/0 | 0/0/0 | 0/0/0 | NA/NA/NA | 0/0/0 |
| Wang, 2024 [154] | 3/3/3 | 2/2/2 | 0/0/0 | 0/0/0 | 2/2/2 | 1/1/1 | 0/0/0 | 2/2/2 | 1/1/1 | 3/3/3 | 3/3/3 | 2/2/2 | 3/3/3 | 3/3/3 | 0/0/0 | 0/0/0 | 0/0/0 | NA/NA/NA | 0/0/0 |
| Wang, 2023 [155] | 3/3/3 | 3/3/3 | 3/3/3 | 3/3/3 | 3/3/3 | 2/2/2 | 0/0/0 | 2/3/2 | 3/3/3 | 3/3/3 | 3/3/3 | 2/3/2 | 2/2/2 | 2/2/2 | 2/2/2 | 2/2/2 | 3/3/3 | 3/3/3 | 2/2/2 |
| Wang, 2025 [156] | 3/3/3 | 3/3/3 | 2/2/2 | 2/2/2 | 3/3/3 | 1/1/1 | 0/0/0 | 2/2/2 | 1/1/1 | 3/3/3 | 2/2/2 | 1/1/1 | 2/2/2 | 3/3/3 | 0/0/0 | 0/0/0 | 0/0/0 | NA/NA/NA | 0/0/0 |
| Wang, 2021 [157] | 3/3/3 | 2/2/2 | 0/0/0 | 0/0/0 | 3/2/2 | 1/1/1 | 0/0/0 | 2/2/2 | 1/1/1 | 3/3/3 | 2/2/2 | 2/2/2 | 2/2/2 | 2/2/2 | 3/3/3 | 0/0/0 | 0/0/0 | NA/NA/NA | 0/0/0 |
| Wang, 2020 [158] | 3/3/3 | 2/2/2 | 0/2/2 | 0/0/0 | 2/2/2 | 2/3/3 | 0/0/0 | 2/2/2 | 2/2/2 | 3/3/3 | 2/2/2 | 2/2/2 | 2/2/2 | 2/2/2 | 0/0/0 | 0/0/0 | 0/0/0 | 0/0/0 | 0/0/0 |
| Wang, 2024 [159] | 3/3/3 | 2/2/2 | 2/2/2 | 0/0/0 | 2/2/2 | 1/1/1 | 0/0/0 | 2/2/2 | 1/1/1 | 3/3/3 | 2/2/2 | 2/2/2 | 2/2/2 | 2/2/2 | 2/2/2 | 0/0/0 | 0/0/0 | NA/NA/NA | 0/0/0 |
| Wang, 2023 [160] | 3/3/3 | 2/2/2 | 2/2/2 | 0/0/0 | 2/2/2 | 1/1/1 | 0/0/0 | 2/2/2 | 0/0/0 | 3/3/3 | 3/3/3 | 2/2/2 | 2/2/2 | 2/2/2 | 0/0/0 | 0/0/0 | 0/0/0 | NA/NA/NA | 0/0/0 |
| Ward, 2022 [161] | 3/3/3 | 3/3/3 | 3/3/3 | 2/3/2 | 2/2/2 | 2/2/2 | 0/0/0 | 2/3/3 | 3/3/3 | 3/3/3 | 0/0/0 | 2/2/2 | 2/2/2 | 2/3/2 | 0/0/0 | 3/3/3 | 3/3/3 | 3/3/3 | 0/0/0 |
| Wei, 2024 [162] | 3/3/3 | 2/2/2 | 0/0/0 | 0/0/0 | 2/2/2 | 1/1/1 | 0/0/0 | 2/2/2 | 1/1/1 | 3/3/3 | 2/2/2 | 2/2/2 | 2/2/2 | 2/2/2 | 0/0/0 | 2/2/2 | 0/0/0 | NA/NA/NA | 3/3/3 |
| Wei, 2023 [163] | 3/3/3 | 3/3/3 | 2/2/2 | 0/0/0 | 2/2/2 | 1/1/1 | 0/0/0 | 2/2/2 | 1/1/1 | 3/3/3 | 3/3/3 | 3/3/3 | 3/3/3 | 2/2/2 | 0/0/0 | 0/0/0 | 0/0/0 | NA/NA/NA | 0/0/0 |
| Wu, 2024 [164] | 3/3/3 | 3/3/3 | 2/2/2 | 2/2/2 | 2/2/2 | 1/1/1 | 0/0/0 | 2/2/2 | 1/1/1 | 3/3/3 | 3/3/3 | 2/2/2 | 2/2/2 | 2/2/2 | 0/0/0 | 2/2/2 | 0/0/0 | 0/0/0 | 3/3/3 |
| Xi, 2022 [165] | 3/3/3 | 2/2/2 | 0/0/0 | 0/0/0 | 2/2/2 | 1/1/1 | 0/0/0 | 2/2/2 | 1/1/1 | 3/3/3 | 0/0/0 | 2/2/2 | 2/3/2 | 2/2/2 | 0/0/0 | 0/0/0 | 2/2/2 | NA/NA/NA | 0/0/0 |
| Xu, 2024 [166] | 3/3/3 | 3/3/3 | 0/0/0 | 0/0/0 | 3/3/3 | 1/1/1 | 0/0/0 | 3/3/3 | 1/1/1 | 3/3/3 | 3/3/3 | 2/2/2 | 2/2/2 | 2/2/2 | 0/0/0 | 0/0/0 | 0/0/0 | NA/NA/NA | 3/3/3 |
| Xue, 2022 [167] | 3/3/3 | 2/2/2 | 2/2/2 | 0/0/0 | 2/2/2 | 1/1/1 | 0/0/0 | 3/3/3 | 1/1/1 | 3/3/3 | 3/3/3 | 3/3/3 | 2/2/2 | 2/2/2 | 0/0/0 | 0/0/0 | 0/0/0 | NA/NA/NA | 0/0/0 |
| Yamazaki, 2020 [168] | 3/3/3 | 2/2/2 | 0/0/0 | 0/0/0 | 2/2/2 | 3/3/3 | 0/0/0 | 2/2/2 | 2/2/2 | 3/3/3 | 2/2/2 | 2/3/2 | 2/2/2 | 2/2/2 | 0/0/0 | 0/0/0 | 3/3/3 | 3/3/3 | 0/0/0 |
| Yamlahi, 2023 [169] | 3/3/3 | 3/3/3 | 0/0/0 | 0/0/0 | 2/2/2 | 1/1/1 | 0/0/0 | 2/2/2 | 1/1/1 | 3/3/3 | 3/3/3 | 1/1/1 | 2/2/2 | 0/0/0 | 0/0/0 | 0/0/0 | 0/0/0 | NA/NA/NA | 0/0/0 |
| Yang, 2024 [170] | 3/3/3 | 3/3/3 | 0/0/0 | 0/0/0 | 2/2/2 | 1/1/1 | 1/1/1 | 0/0/0 | 1/1/1 | 2/2/2 | 0/0/0 | 2/2/2 | 2/2/2 | 0/0/0 | 0/0/0 | 2/2/2 | 2/2/2 | NA/NA/NA | 0/0/0 |
| Yang, 2023 [171] | 3/3/3 | 3/3/3 | 2/2/2 | 2/2/2 | 2/2/2 | 2/2/2 | 0/2/0 | 2/2/2 | 2/2/2 | 3/3/3 | 3/3/3 | 2/2/2 | 2/2/2 | 2/2/2 | 0/0/0 | 0/0/0 | 0/0/0 | 2/2/2 | 0/0/0 |
| Yang, 2021 [172] | 3/3/3 | 3/3/3 | 2/2/2 | 0/0/0 | 2/2/2 | 1/1/1 | 0/0/0 | 2/2/2 | 1/1/1 | 3/3/3 | 3/3/3 | 2/2/2 | 2/2/2 | 0/0/0 | 3/3/3 | 0/0/0 | 2/2/2 | NA/NA/NA | 3/3/3 |
| Yin, 2024 [173] | 3/3/3 | 3/3/3 | 2/2/2 | 2/2/2 | 2/2/2 | 1/1/1 | 0/0/0 | 2/2/2 | 1/1/1 | 3/3/3 | 2/2/2 | 2/2/2 | 2/2/2 | 2/2/2 | 0/0/0 | 2/2/2 | 2/2/2 | NA/NA/NA | 0/0/0 |
| Yuan, 2024 [174] | 3/3/3 | 3/3/3 | 2/2/2 | 2/2/2 | 2/2/2 | 1/1/1 | 2/2/2 | 1/1/1 | 1/1/1 | 3/3/3 | 2/2/2 | 2/2/2 | 2/2/2 | 2/2/2 | 0/0/0 | 0/0/0 | 0/0/0 | NA/NA/NA | 3/3/3 |
| Zhang, 2024 [175] | 3/3/3 | 3/3/3 | 2/2/2 | 2/2/2 | 2/2/2 | 2/2/2 | 0/0/0 | 2/2/2 | 2/2/2 | 3/3/3 | 3/3/3 | 2/2/2 | 2/2/2 | 2/2/2 | 0/0/0 | 2/2/2 | 2/2/2 | 3/3/3 | 0/0/0 |
| Zhang, 2023 [176] | 3/3/3 | 3/3/3 | 2/2/2 | 2/2/2 | 2/2/2 | 2/2/2 | 0/0/0 | 2/2/2 | 2/2/2 | 3/3/3 | 2/2/2 | 2/2/2 | 3/3/3 | 2/2/2 | 2/2/2 | 3/3/3 | 3/3/3 | 3/3/3 | 3/3/3 |
| Zhang, 2023 [177] | 3/3/3 | 2/2/2 | 0/0/0 | 2/3/3 | 2/2/2 | 1/1/1 | 0/0/0 | 2/2/2 | 1/1/1 | 3/3/3 | 2/2/2 | 2/2/2 | 2/2/2 | 2/2/2 | 2/2/2 | 2/2/2 | 2/2/2 | NA/NA/NA | 0/0/0 |
| Zhang, 2020 [178] | 3/3/3 | 2/2/2 | 2/2/2 | 2/3/2 | 2/2/2 | 1/3/1 | 0/0/0 | 2/2/2 | 2/2/2 | 3/3/3 | 2/2/2 | 2/2/2 | 2/2/2 | 0/0/0 | 0/0/0 | 2/2/2 | 0/0/0 | NA/NA/NA | 0/0/0 |
| Zhang, 2023 [179] | 3/3/3 | 2/2/2 | 0/0/0 | 0/0/0 | 2/2/2 | 2/3/3 | 0/0/0 | 2/2/2 | 2/2/2 | 3/3/3 | 2/2/2 | 2/2/2 | 2/2/2 | 2/2/2 | 0/0/0 | 0/0/0 | 0/0/0 | 0/0/0 | 0/0/0 |
| Zhang, 2020 [180] | 3/3/3 | 3/3/3 | 3/3/3 | 3/3/3 | 2/2/2 | 1/1/1 | 0/0/0 | 2/2/2 | 1/0/1 | 3/3/3 | 2/2/2 | 2/2/2 | 2/2/2 | 2/2/2 | 0/0/0 | 3/3/3 | 0/0/0 | NA/NA/NA | 0/0/0 |
| Zhao, 2021 [181] | 3/3/3 | 3/3/3 | 2/2/2 | 2/2/2 | 3/3/3 | 2/2/2 | 0/0/0 | 2/2/2 | 1/1/1 | 3/3/3 | 2/2/2 | 2/2/2 | 2/2/2 | 0/0/0 | 0/0/0 | 2/2/2 | 2/2/2 | 0/2/0 | 0/2/0 |
| Zhao, 2023 [182] | 3/3/3 | 3/3/3 | 0/3/0 | 0/2/0 | 2/2/2 | 1/1/1 | 0/0/0 | 2/2/2 | 2/2/2 | 3/3/3 | 3/3/3 | 2/2/2 | 2/2/2 | 2/2/2 | 0/0/0 | 0/0/0 | 0/0/0 | NA/NA/NA | 0/0/0 |
| Zhao, 2021 [183] | 3/3/3 | 2/2/2 | 0/0/0 | 0/0/0 | 2/2/2 | 1/1/1 | 0/0/0 | 2/2/2 | 1/1/1 | 3/3/3 | 3/3/3 | 2/2/2 | 2/2/2 | 2/2/2 | 0/0/0 | 0/0/0 | 0/0/0 | NA/NA/NA | 0/0/0 |
| Zhao, 2019 [184] | 3/3/3 | 3/3/3 | 2/2/2 | 2/2/2 | 2/2/2 | 2/1/2 | 0/0/0 | 2/2/2 | 0/0/0 | 3/3/3 | 3/3/3 | 2/2/2 | 3/3/3 | 2/2/2 | 2/2/2 | 0/0/0 | 0/0/0 | 0/0/0 | 0/0/0 |
| Zheng, 2023 [185] | 3/3/3 | 3/3/3 | 2/2/2 | 2/3/3 | 2/2/2 | 2/2/2 | 2/2/2 | 3/3/3 | 3/3/3 | 3/3/3 | 2/2/2 | 2/2/2 | 2/2/2 | 2/2/2 | 0/0/0 | 3/3/3 | 3/3/3 | 3/3/3 | 2/3/2 |
| Zhou, 2024 [186] | 3/3/3 | 2/2/2 | 0/2/2 | 0/2/2 | 2/2/2 | 1/1/1 | 0/0/0 | 0/0/0 | 2/2/2 | 3/3/3 | 2/2/2 | 2/2/2 | 2/2/2 | 0/0/0 | 0/0/0 | 0/0/0 | 0/0/0 | NA/NA/NA | 0/0/0 |
| Zhou, 2022 [187] | 3/3/3 | 3/3/3 | 2/2/2 | 2/2/2 | 2/2/2 | 1/1/1 | 0/0/0 | 2/2/2 | 2/2/2 | 3/3/3 | 2/2/2 | 2/2/2 | 2/2/2 | 2/2/2 | 0/0/0 | 0/0/0 | 0/0/0 | NA/NA/NA | 0/0/0 |
| Zygomalas, 2024 [188] | 3/3/3 | 2/2/2 | 2/2/2 | 2/2/2 | 0/0/0 | 2/2/2 | 0/0/0 | 0/0/0 | 2/2/2 | 3/3/3 | 0/0/0 | 2/2/2 | 2/2/2 | 0/0/0 | 2/2/2 | 2/2/2 | 2/2/2 | 2/2/2 | 0/0/0 |

#### **Supplementary Table 8**: Reporting Quality Across JCR-Based Journal Categories

All journals were categorized into three groups based on Clarivate Journal Citation Reports (JCR) categories: medical journals (purely medical content), interdisciplinary journals (medical and technical), and technical-only journals (primarily engineering, biomedical engineering, robotics, or computer science). Each reporting quality category was compared between these groups using the Kruskal-Wallis test, followed by Dunn's post hoc tests with Bonferroni correction for pairwise comparisons. Table 8 shows the medians for each reporting quality criterion across the JCR-based journal groups, using the following coding: Reported = 3, Partially Reported = 2, Reference to another publication = 1, Not Reported = 0, and displays the p-values from the Kruskal–Wallis tests and Dunn's post hoc comparisons.

| Reporting Quality Criterium | JCR Group | Median | P-value (Kruskal-Wallis test) | Group 1 | Group 2 | P-value (after Bonferroni correction) (Dunn test) |
|---|---|---|---|---|---|---|
| Study Design | Medical Journals | 3 | 0.499 | Medical Journals | Multidisciplinary Journals | 1 |
| | Multidisciplinary Journals | 3 | | Medical Journals | Technical Journals | 0.765325324994855 |
| | Technical Journals | 3 | | Multidisciplinary Journals | Technical Journals | 1 |
| Prediction Problem | Medical Journals | 3 | 0.00015 | Medical Journals | Multidisciplinary Journals | 0.073132530180647 |
| | Multidisciplinary Journals | 3 | | Medical Journals | Technical Journals | 9.9850717723128E-05 |
| | Technical Journals | 2 | | Multidisciplinary Journals | Technical Journals | 0.0537047330531948 |
| Clinical Problem | Medical Journals | 3 | 7.92E-09 | Medical Journals | Multidisciplinary Journals | 2.73016394291022E-07 |
| | Multidisciplinary Journals | 2 | | Medical Journals | Technical Journals | 2.88425703895975E-08 |
| | Technical Journals | 2 | | Multidisciplinary Journals | Technical Journals | 1 |
| Clinical Goal | Medical Journals | 2 | 2.55E-07 | Medical Journals | Multidisciplinary Journals | 2.9449504203937E-05 |
| | Multidisciplinary Journals | 2 | | Medical Journals | Technical Journals | 2.21253639239217E-07 |
| | Technical Journals | 0 | | Multidisciplinary Journals | Technical Journals | 0.60143721403075 |
| Existing AI | Medical Journals | 2 | 0.108 | Medical Journals | Multidisciplinary Journals | 1 |
| | Multidisciplinary Journals | 2 | | Medical Journals | Technical Journals | 0.824635275494482 |
| | Technical Journals | 2 | | Multidisciplinary Journals | Technical Journals | 0.107407641222633 |
| Data Source | Medical Journals | 2 | 1.31E-11 | Medical Journals | Multidisciplinary Journals | 9.96126730348103E-08 |
| | Multidisciplinary Journals | 1 | | Medical Journals | Technical Journals | 9.86787604187975E-12 |
| | Technical Journals | 1 | | Multidisciplinary Journals | Technical Journals | 0.175164604457521 |
| Data Selection | Medical Journals | 2 | 1.67E-08 | Medical Journals | Multidisciplinary Journals | 6.09188781125351E-05 |
| | Multidisciplinary Journals | 0 | | Medical Journals | Technical Journals | 7.2810454428434E-09 |
| | Technical Journals | 0 | | Multidisciplinary Journals | Technical Journals | 0.0873072048233087 |
| Data Preprocessing | Medical Journals | 2 | 0.611 | Medical Journals | Multidisciplinary Journals | 1 |
| | Multidisciplinary Journals | 2 | | Medical Journals | Technical Journals | 1 |
| | Technical Journals | 2 | | Multidisciplinary Journals | Technical Journals | 1 |
| Labeling | Medical Journals | 3 | 1.44E-08 | Medical Journals | Multidisciplinary Journals | 6.67666677096481E-08 |
| | Multidisciplinary Journals | 1 | | Medical Journals | Technical Journals | 2.9947559703384E-07 |
| | Technical Journals | 1 | | Multidisciplinary Journals | Technical Journals | 1 |
| Model Type | Medical Journals | 3 | 0.0553 | Medical Journals | Multidisciplinary Journals | 0.0556118184830427 |
| | Multidisciplinary Journals | 3 | | Medical Journals | Technical Journals | 0.165249650645181 |
| | Technical Journals | 3 | | Multidisciplinary Journals | Technical Journals | 1 |
| Model Development | Medical Journals | 2 | 0.00225 | Medical Journals | Multidisciplinary Journals | 0.00514704999470532 |
| | Multidisciplinary Journals | 2 | | Medical Journals | Technical Journals | 1 |
| | Technical Journals | 2 | | Multidisciplinary Journals | Technical Journals | 0.0242649184928813 |
| Model Validation | Medical Journals | 2 | 0.4 | Medical Journals | Multidisciplinary Journals | 1 |
| | Multidisciplinary Journals | 2 | | Medical Journals | Technical Journals | 0.528331531720964 |
| | Technical Journals | 2 | | Multidisciplinary Journals | Technical Journals | 1 |
| Model Performance | Medical Journals | 2 | 0.00305 | Medical Journals | Multidisciplinary Journals | 0.0652087331103511 |
| | Multidisciplinary Journals | 2 | | Medical Journals | Technical Journals | 1 |
| | Technical Journals | 2 | | Multidisciplinary Journals | Technical Journals | 0.00391209438840239 |
| Statistical Methods | Medical Journals | 2 | 0.0867 | Medical Journals | Multidisciplinary Journals | 1 |
| | Multidisciplinary Journals | 2 | | Medical Journals | Technical Journals | 0.916285120635758 |
| | Technical Journals | 2 | | Multidisciplinary Journals | Technical Journals | 0.0812204904855827 |
| Performance Errors | Medical Journals | 0 | 0.0546 | Medical Journals | Multidisciplinary Journals | 0.218077119662363 |
| | Multidisciplinary Journals | 0 | | Medical Journals | Technical Journals | 1 |
| | Technical Journals | 0 | | Multidisciplinary Journals | Technical Journals | 0.0860241498328219 |
| Clinical Implications | Medical Journals | 3 | 3.71E-11 | Medical Journals | Multidisciplinary Journals | 1.61284089238289E-08 |
| | Multidisciplinary Journals | 0 | | Medical Journals | Technical Journals | 8.81760085061061E-11 |
| | Technical Journals | 0 | | Multidisciplinary Journals | Technical Journals | 0.7656533258143 |
| Limitations | Medical Journals | 3 | 7.93E-13 | Medical Journals | Multidisciplinary Journals | 6.74958540629431E-11 |
| | Multidisciplinary Journals | 0 | | Medical Journals | Technical Journals | 1.69258447462909E-11 |
| | Technical Journals | 0 | | Multidisciplinary Journals | Technical Journals | 1 |
| Ethical Statement | Medical Journals | 3 | 1.72E-05 | Medical Journals | Multidisciplinary Journals | 0.0648486908940682 |
| | Multidisciplinary Journals | 3 | | Medical Journals | Technical Journals | 1.33805329647371E-05 |
| | Technical Journals | 0 | | Multidisciplinary Journals | Technical Journals | 0.0132301601652264 |
| Code Publication | Medical Journals | 0 | 0.0138 | Medical Journals | Multidisciplinary Journals | 0.983601033186889 |
| | Multidisciplinary Journals | 0 | | Medical Journals | Technical Journals | 0.449472967005708 |
| | Technical Journals | 0 | | Multidisciplinary Journals | Technical Journals | 0.0105135703624522 |

**Supplementary Figure 2**: Reporting Quality Across JCR-Based Journal Categories

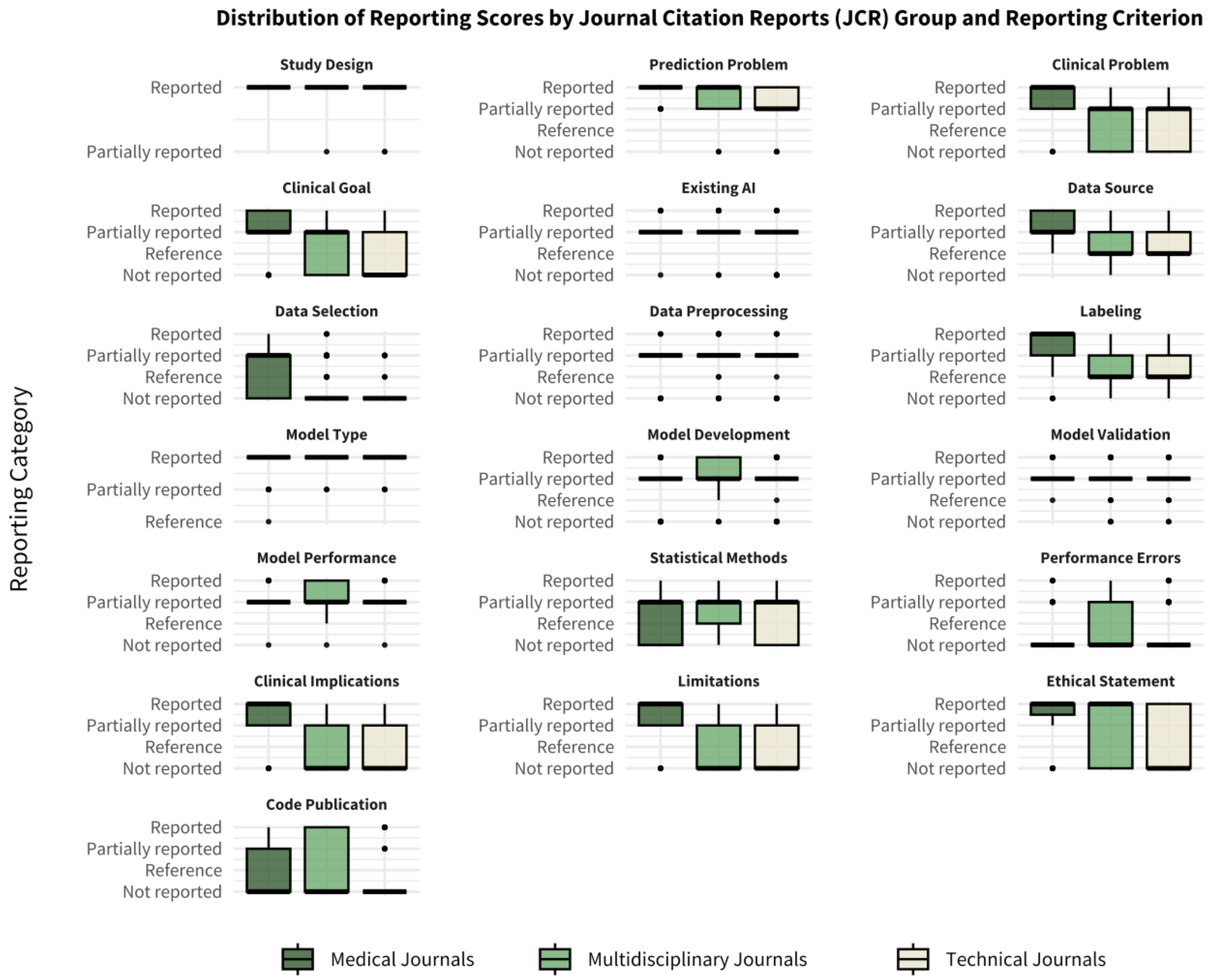

Boxplots show the distribution of reporting quality scores by category and Clarivate Journal Citation Reports (JCR) category group, based on the journal type: medical journals (purely medical content), interdisciplinary journals (medical and technical), and technical-only journals (primarily engineering, biomedical engineering, robotics, or computer science). The central line represents the median, the box shows the interquartile range (IQR: 25th–75th percentile), and the whiskers extend to the most extreme data points within 1.5×IQR from the box. Individual points represent outliers beyond this range.
